# Mathematical Modeling of Japanese Encephalitis: Multi-Host Transmission Dynamics and Intervention Strategies

**DOI:** 10.64898/2026.08.25.26361297

**Authors:** Mangala C. Devihosoor, K. P. Suresh, P V Sudarshan, T. R. Devika, Jagadish Hiremath, P. P. Sengupta

## Abstract

Japanese encephalitis virus (JEV) transmission involves complex interactions among Culex mosquitoes, amplifying pig hosts, reservoir wading birds, humans, and environmental conditions, complicating quantitative assessment of transmission dynamics and intervention effectiveness. We developed a deterministic, fourteen-compartment One Health mathematical framework that integrates these interconnected host–vector populations and their epidemiological states. The model incorporates temperature-dependent mosquito biting, seasonal transmission, human vaccination, pig biosecurity, environmental barriers, and mosquito-control interventions. Mathematical properties were established through analyses of non-negativity, boundedness, biologically feasible equilibria, local and global stability, and optimal control. District-specific simulations were conducted for Bellary, Udupi, Kolkata, and Purba Bardhaman during the August transmission period. Intervention scenarios were evaluated, and global sensitivity analysis was performed using 500 Latin hypercube samples with partial rank correlation coefficients. Model outputs were also compared with district-level surveillance observations. Vaccination-adjusted basic reproduction numbers were 0.905 in Bellary, 0.965 in Udupi, 1.817 in Kolkata, and 0.885 in Purba Bardhaman, with only Kolkata exceeding the epidemic threshold. Under maximum intervention, total infections decreased by 80.6%, 96.8%, 80.5%, and 72.2%, respectively, while infected mosquito populations declined to zero across all four settings. In Kolkata, vaccinating 3.6 million individuals with dose series II reduced the reproduction number from 1.817 to 0.9846, whereas population-wide dose series I vaccination alone was insufficient to reduce it below unity. Sensitivity analysis identified mosquito recruitment, temperature-dependent biting, carrying capacity, mosquito mortality, density-dependent regulation, and mosquito-to-human transmission as major determinants of peak human infection. Overall, the framework demonstrates heterogeneity in JEV transmission and intervention effectiveness and provides a mathematically grounded One Health approach for comparative evaluation of integrated control strategies.

## 1. Introduction

Japanese encephalitis (JE) is a mosquito-borne zoonotic disease caused by the Japanese encephalitis virus (JEV), a flavivirus transmitted primarily by *Culex* mosquitoes. Endemic transmission occurs in 24 countries across the World Health Organization (WHO) South-East Asia and Western Pacific regions, placing more than three billion people at risk (World Health Organization, 2024). Although most infections are asymptomatic, severe cases may progress to encephalitis, with case-fatality rates of up to 30%, while 30–50% of survivors experience long-term neurological, cognitive, or behavioural sequelae. Recent estimates indicate that approximately 100,000 clinical cases and 25,000 deaths occur globally each year (Quan et al., 2020; World Health Organization, 2024).

JEV is maintained through an enzootic transmission cycle involving *Culex* mosquitoes, domestic pigs, and wading birds. Pigs are the principal amplifying hosts because they develop sufficient viraemia to infect mosquitoes, whereas wading birds act as reservoir hosts that contribute to viral maintenance and dispersal. Humans are incidental dead-end hosts because they generally do not develop viraemia sufficient for onward transmission. Consequently, human infection depends on the interactions among vectors, animal hosts, environmental conditions, and human populations. Transmission is favoured in rice-growing and irrigated agricultural landscapes where mosquito breeding habitats overlap with livestock and human settlements. Environmental factors such as temperature, rainfall, land use, and agricultural practices strongly influence mosquito abundance, biting behaviour, and transmission dynamics (Le Flohic et al., 2013; Walsh et al., 2022).

In India, JE remains an important public health concern, particularly in rural and agricultural regions. The first confirmed case was reported in Vellore in 1955, and the disease is now endemic in several states. The principal vector, *Culex tritaeniorhynchus*, is widely distributed across irrigated, coastal, and sub-Himalayan regions, contributing to seasonal outbreaks (Das et al., 2023). Although safe and effective vaccines are available, incomplete vaccination coverage, limited diagnostic capacity, inadequate surveillance, and environmental changes continue to hinder effective disease prevention and control.

Mathematical modelling provides a systematic framework for understanding JEV transmission dynamics, estimating epidemiological thresholds, and evaluating disease-control strategies. Previous studies have investigated human–pig–mosquito transmission (Baniya and Keval, 2020), livestock composition and host interactions (Ladreyt et al., 2022), environmental factors influencing mosquito development (Ndaïrou et al., 2020), climatic determinants of vector abundance (Franklinos et al., 2022), and JEV–dengue co-infection dynamics (Dwivedi et al., 2024). However, most existing models focus primarily on humans, pigs, and mosquitoes while neglecting the explicit role of wading birds in viral maintenance. Climatic influences are often simplified, and interventions such as vaccination, biosecurity, environmental barriers, and vector control are rarely evaluated together within a single modelling framework. In addition, many models rely on assumed or literature-derived parameter values, limiting their applicability to local epidemiological settings (Laidlow et al., 2025).

To address these limitations, the present study develops a comprehensive multi-host mathematical model for JEV transmission involving humans, pigs, mosquitoes, and wading birds. The model incorporates host-specific transmission pathways, temperature-dependent mosquito biting behaviour, and multiple intervention strategies, including vaccination, farm-level biosecurity, environmental barriers, and mosquito-control measures. In addition, a structured questionnaire-based protocol is proposed to estimate location-specific epidemiological and livestock-management parameters, thereby improving model parameterization and enhancing its practical applicability for regional disease surveillance and control.

The present study aims to develop a multi-host mathematical model of Japanese encephalitis virus transmission involving humans, pigs, mosquitoes, and wading birds. The specific objectives are to: (i) formulate the transmission dynamics among these four populations using a compartmental modelling framework; (ii) incorporate temperature-dependent mosquito biting, seasonal variation, vaccination, biosecurity, environmental barriers, and vector-control measures; (iii) estimate the basic and time-varying reproduction numbers and examine the disease-free and endemic equilibrium states; (iv) compare disease transmission with and without selected control measures, including different human vaccination strategies; and (v) identify the main epidemiological and ecological parameters influencing JE transmission through local and global sensitivity analyses. The inclusion of wading birds together with pigs, mosquitoes, and humans provides a broader representation of the JE transmission cycle within a single modelling framework.

## 2. Materials and Methods

### 2.1 Model framework and assumptions

The model was formulated as a deterministic system of nonlinear ordinary differential equations describing interactions among humans, pigs, wading birds, and mosquitoes. Homogeneous mixing was assumed within each district. Mosquitoes transmit Japanese encephalitis virus to humans, pigs, and wading birds, whereas pigs and wading birds contribute to infection of susceptible mosquitoes. Humans were treated as dead-end hosts in the biological baseline; the human-to-mosquito coefficient was retained only as a hypothetical sensitivity parameter. Demographic recruitment and mortality were included, and logistic density regulation was applied to the mosquito and wading-bird subsystems. Control measures included vaccination, treatment, pig biosecurity, environmental barriers, and mosquito-control interventions.

### 2.2 Population compartments and transmission pathways

The proposed mathematical model in Figure 1 captures the transmission dynamics of JEV by integrating four interconnected populations: wading birds, pigs, mosquitoes, and humans. Each population is subdivided into compartments representing key epidemiological states, and the interactions among these compartments reflect ecological transmission cycles and control strategies.

**Fig. 1.**
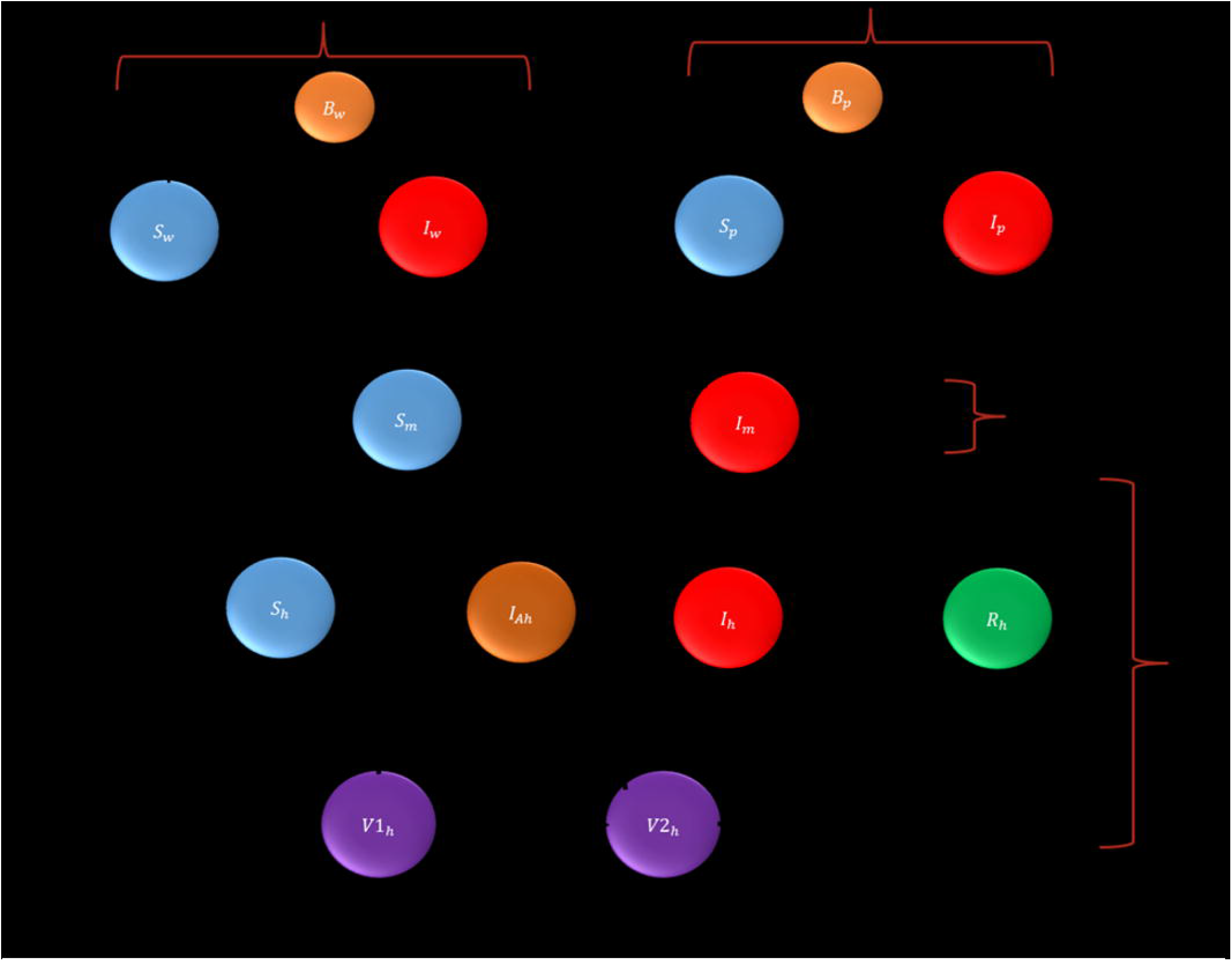
JE Transmission Pathways in a Multi-Host with SVIIR in humans, SI in

In the wading bird population, individuals are categorized as susceptible (*S*_*w*_), infectious (*I*_*p*_) and barrier (*B*_*w*_) compartments. *S*_*w*_ may become infected through bites from infectious mosquitoes (*I*_*m*_), with the rate of infection influenced by the temperature-dependent biting rate *b*(*T*). A portion of *S*_*w*_ transitions into the barrier class (*B*_*w*_) at rate *ε*_*w*_, representing birds shielded by physical or environmental interventions. All bird compartments experience natural mortality at a rate *μ*_*w*_.

In the pig population, individuals are grouped into susceptible (*Sp*), infectious (*Ip*) and bio-secured (*B*_*p*_) compartments. *S*_*p*_ may become infected by *I*_*m*_, while a subset of *S*_*p*_is moved into a *B*_*p*_ at rate *ε*_*p*_, representing enhanced containment through farm-level interventions. *I*_*p*_ may be returned to the *S*_*p*_ through treatment at rate *α*_2_, while natural death occurs in all pig compartments at a rate *μ*_*p*_

The mosquito population includes susceptible (*S*_*m*_) and infectious (*I*_*m*_) compartments. *S*_*m*_ become *I*_*m*_ by biting *I*_*p*_ and *I*_*w*_. The force of infection depends on the average biting rates on pigs (C) and birds (B), as well as the temperature-modulated biting rate *b*(*T*). The mosquito population is regulated by intrinsic growth (*r*_*m*_) and carrying capacity (*K*_*m*_), capturing environmental and seasonal constraints, while mortality occurs at rate *μm*.

The human population is divided into susceptible (*S*_*h*_), infectious symptomatic and asymptomatic (*I*_*h*_ & *I*_*ah*_), recovered (*R*_*h*_) and vaccinated groups. Susceptible individuals are vaccinated at a rate *u*_1*α*1_, into dose series I*V*1_*h*_, and subsequently at a rate (1-*u*_1_) *α*_1_ into dose series II *V*2_*h*_. *S*_*h*_ acquire infection through bites from *Im*, with additional risk from travel exposure (*θ*). *I*_*h*_may recover naturally (*τ*_1_) or with medical treatment (*τ*_2_) and move into the *R*_*h*_, while disease-induced death occurs at a rate *σ*. Immunity wanes at rates *ρ*_1_and *ρ*_2_ for dose series I and II, respectively, leading individuals back to the susceptible state.

### 2.3 System of differential equations

Wading Bird: Rice production, Flooding irrigation, water level – spread, Physical or mechanical barrier –control

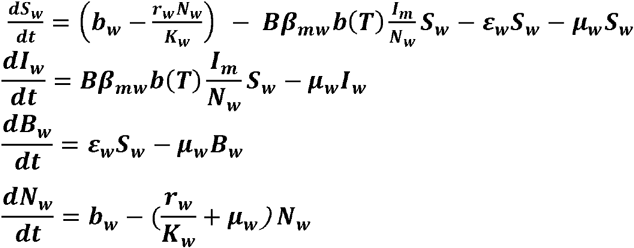

Pigs: Biosecurity measures or preventive measures – control, Vaccination – control

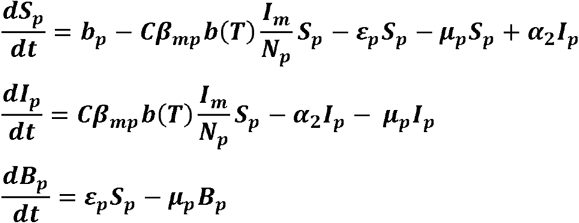

Mosquitoes: Rice production, Flooding irrigation, water level – spread, Environmental factor i.e., temperature – spread

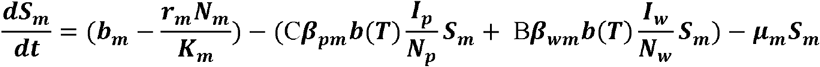

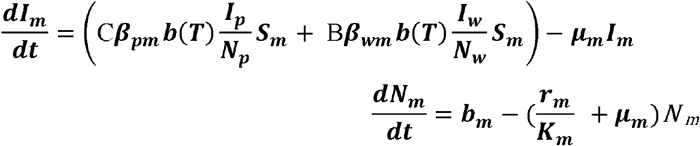

Humans: RRTE – Risk Region Travel Exposure – spread, Vaccination – Control

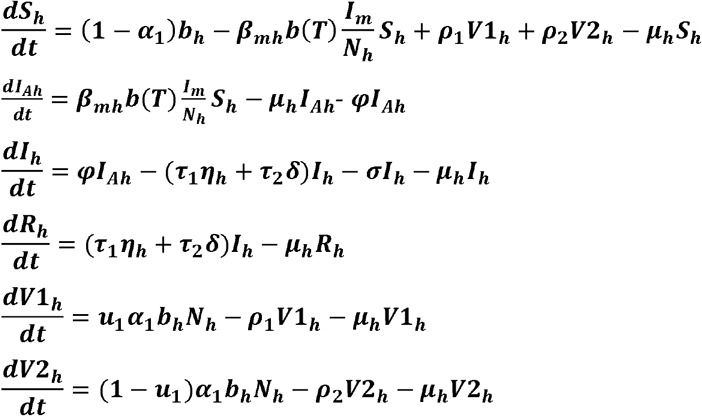

Where:

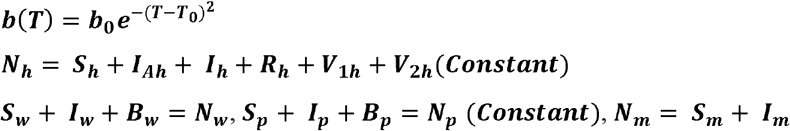

### 2.4 Study districts and data sources

Data used in the district-level simulations were compiled from official surveillance and population records assembled within the Health Management Information System (HMIS) data framework and implemented in the ICAR-NIVEDI Japanese encephalitis simulation dashboard. The complete procedure for operating the model and interactive dashboard is provided in the Technical Support section.

The analysis included Bellary and Udupi districts in Karnataka and Kolkata and Purba Bardhaman districts in West Bengal. Human case, testing, mortality and vaccination inputs were retained at district level, while pig populations were district-specific. Wading-bird and mosquito starting populations and several biological rates were specified as common model inputs.

The initial susceptible compartments were calculated from the corresponding total population after subtracting infectious, recovered, vaccinated, bio-secured or barrier-protected compartments, as applicable. Thus, each population identity was preserved at the start of simulation.

For reproducibility, the exact point values used in the final HMIS district simulations are reported below. District-specific initial conditions are presented in Table 1a, whereas parameters held constant across districts are presented in Table 1b. Broad literature ranges and uniform distributions used during preliminary model development were not reported as final baseline inputs unless they were directly used in the simulation.

**Table 1a.**
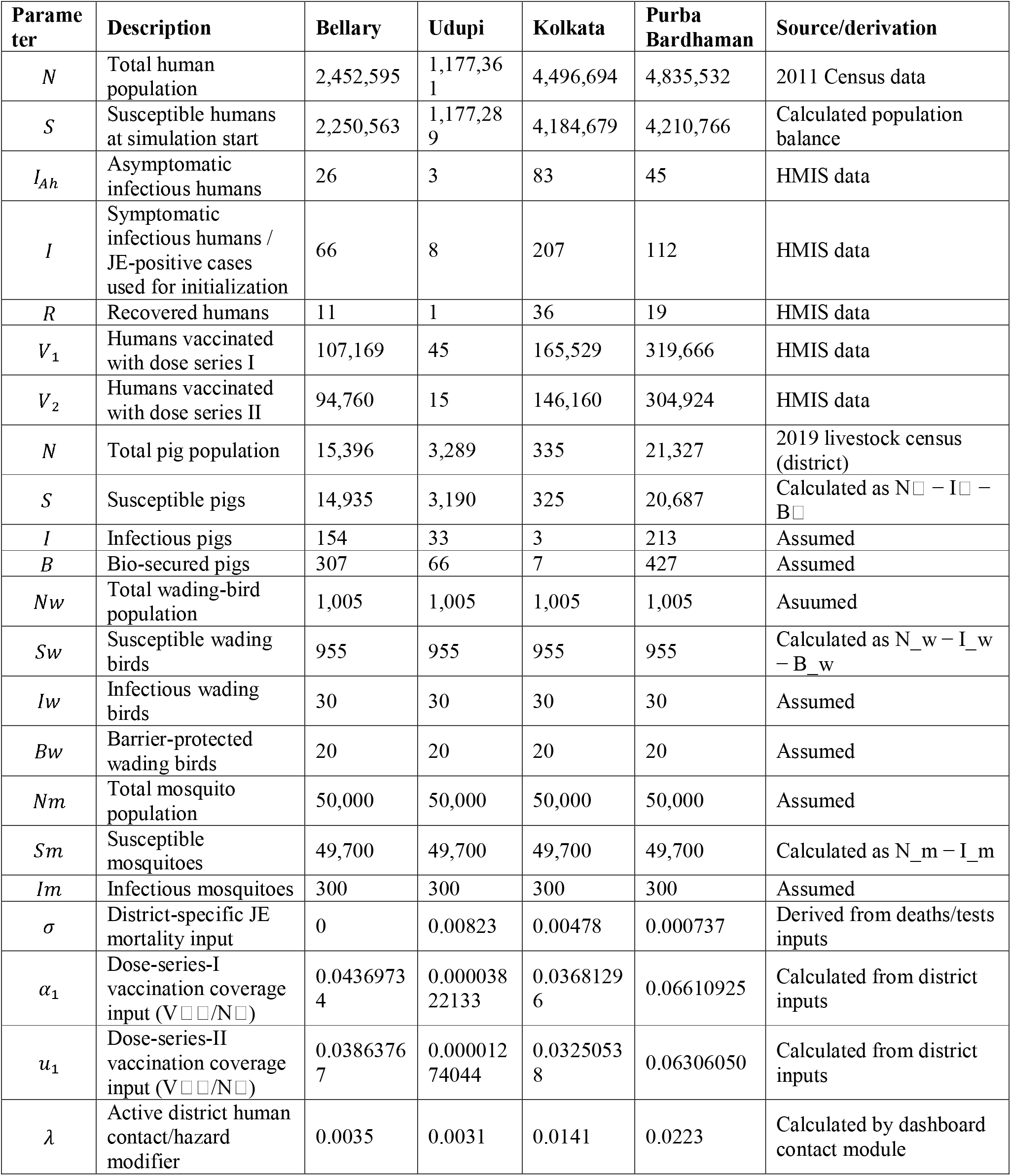
District-specific initial conditions and data-derived inputs used in the Japanese encephalitis simulations.

**Table 1b.**
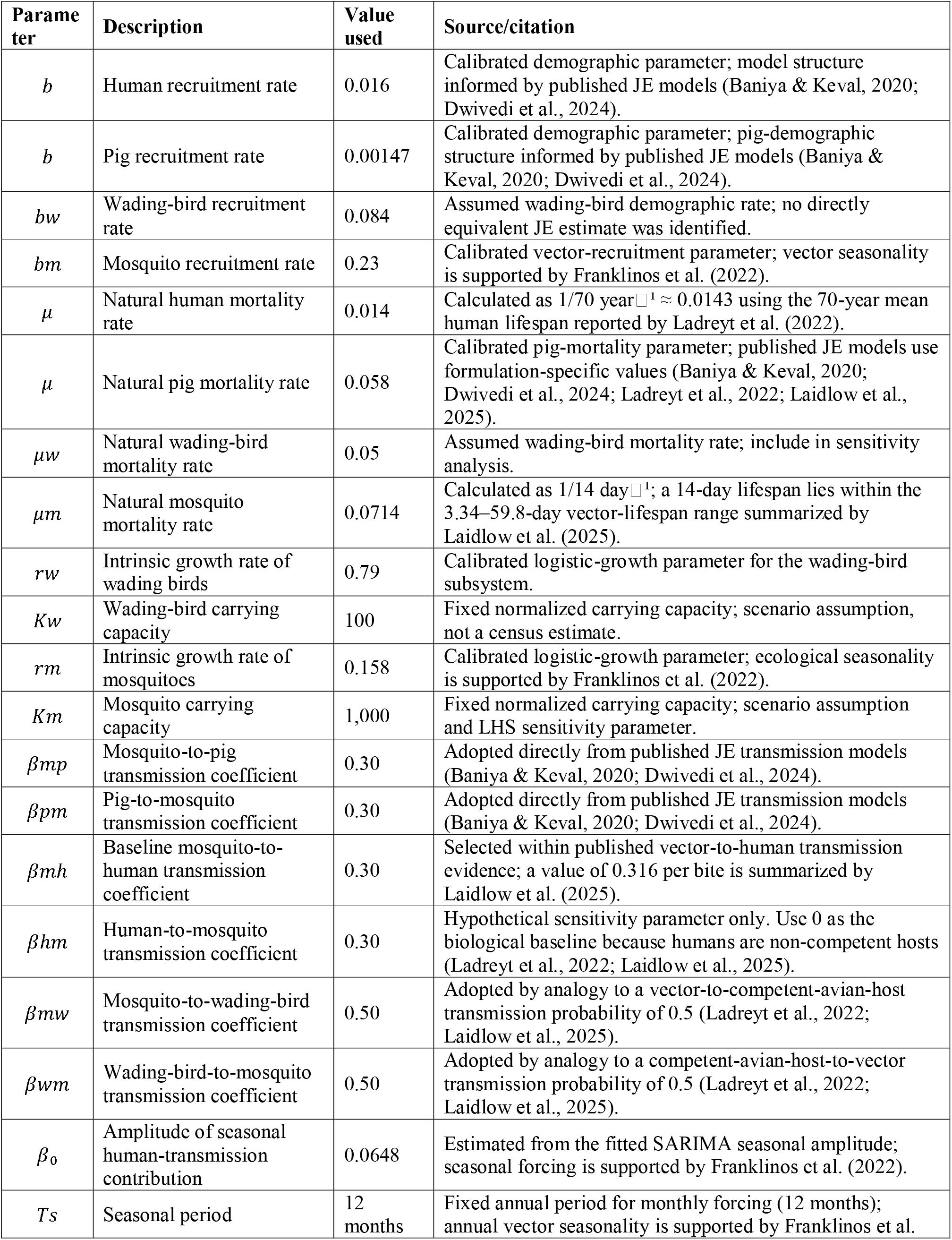
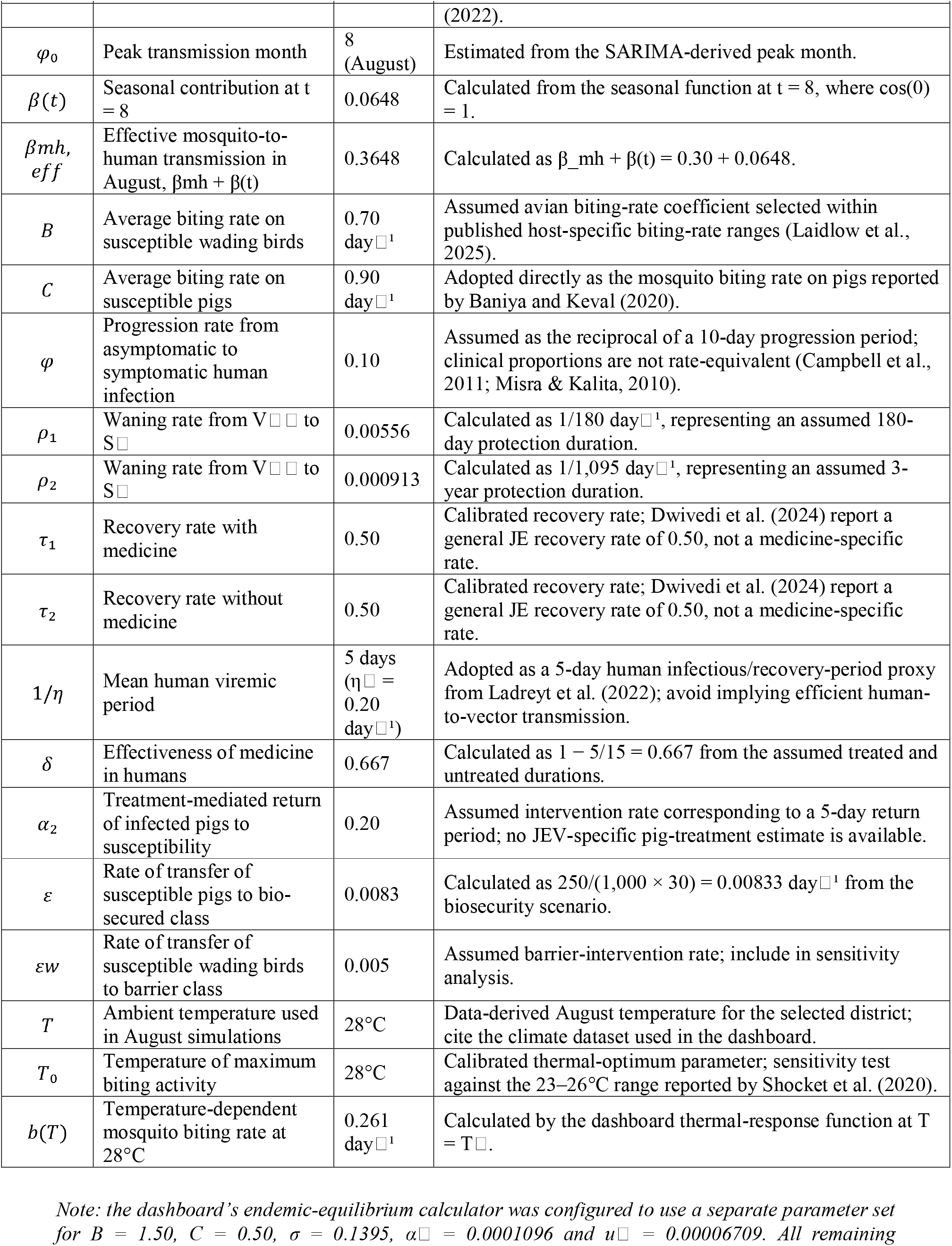
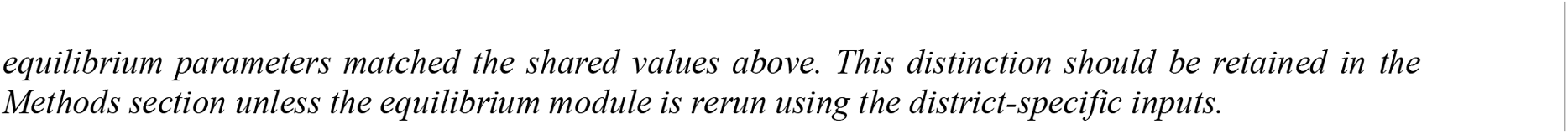
Parameters held constant across the four district simulations.

### 2.5 Initial conditions and model parameterisation

### 2.6 Mathematical analysis

In this section, the fundamental mathematical properties of the proposed Japanese encephalitis transmission model are investigated to establish its theoretical validity and epidemiological consistency. The analysis includes the proof of non-negativity of solutions, boundedness of the feasible region, the existence of biologically meaningful equilibrium points, local and global stability analyses of the equilibrium states, and optimal control analysis. The model admits three biologically meaningful equilibrium points: the vector-free equilibrium (VFE), where the mosquito population is absent; the disease-free equilibrium (DFE), where infection is eliminated from all host and vector populations; and the endemic equilibrium (EE), where the disease persists at a steady state. The stability properties of these equilibrium points provide insights into the threshold conditions under which Japanese encephalitis can either be eliminated or persist in the population. These analytical results demonstrate that the proposed model is mathematically well posed and epidemiologically meaningful. For brevity and to improve the manuscript’s readability, the detailed mathematical proofs and derivations are presented in Appendix A.

#### 2.6.1 Equilibrium points

The mathematical derivations for the positivity (non-negativity) of the solutions, boundedness of the feasible region, and the existence of equilibrium points are presented in Appendix A (Theorems A1– A3).

##### 2.6.1.1 Vector-free equilibrium

The vector-free equilibrium is obtained by setting the mosquito-related compartments (i.e., *S*_*m*_ and *I*_*m*_) to zero, assuming the absence of vectors in the system. The rest of the compartments are solved under the assumption that no mosquito-mediated transmission occurs, thus representing a scenario where JE cannot be sustained due to the lack of vectors.

Thus, the VFE points denoted as (*E*_0_):

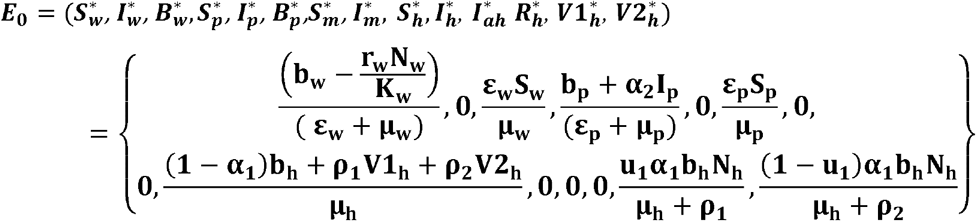

The equilibrium Vector – free equilibrium point (*E*_0_) exist, if *N*_*m*_ =0.

##### 2.6.1.2 Disease-free equilibrium

The disease-free equilibrium is derived by setting all infected compartments to zero, including *I*_*h*_, *I*_*ah*_, *I*_*p*_, *I*_*w*_, and *I*_*m*_, which represent infected humans, pigs, wading birds, and mosquitoes respectively. The system is then solved to find the steady state values of the susceptible and non-infected classes. This equilibrium reflects a state where the disease is absent in all host and vector populations, and transmission does not occur.

Thus, the *DFE* points denoted as 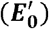:

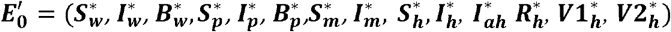

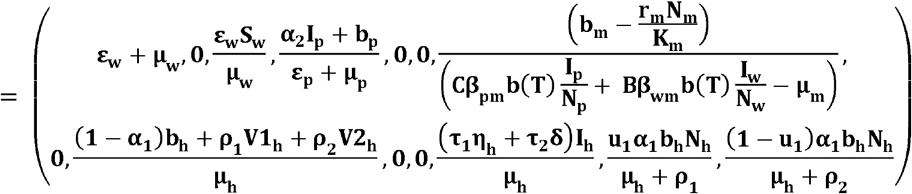

The equilibrium disease–free equilibrium point (*E*_0_) exists, if *N*_*m*_ =*K*_*m*_ and *N*_*w*_ =*K*_*w*_.

##### 2.6.1.3 Endemic equilibrium

The disease endemic equilibrium point is determined by setting the right-hand side of the model equation to zero, solving for the infected and carrier state variables.

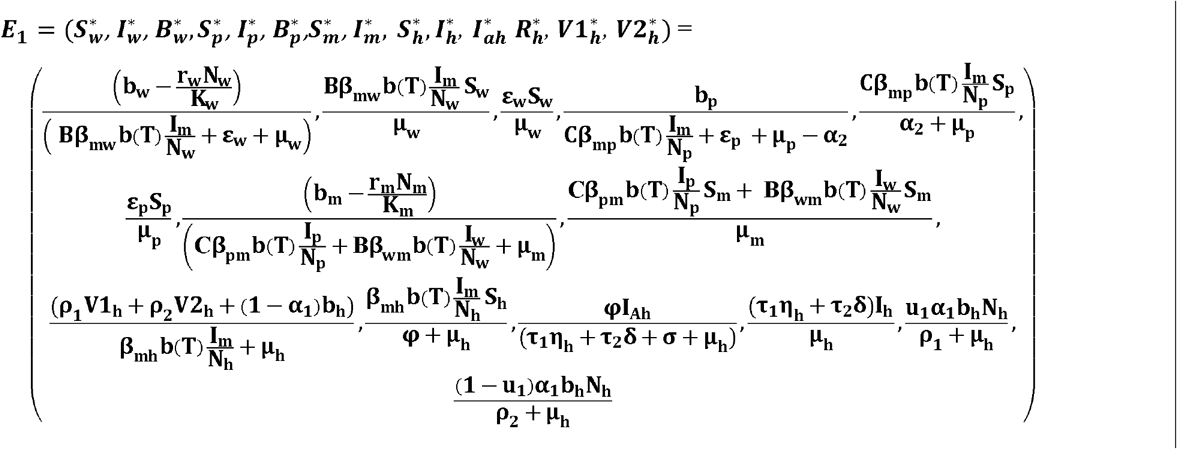

#### 2.6.2 Local stability analysis

Stability analysis is used to assess whether the system will return to its equilibrium state over time. A stable disease-free equilibrium suggests that the infection will eventually die out, while an endemic equilibrium indicates that the disease is likely to persist and spread within the population.

##### Theorem 2

The vector-free equilibrium point *E*^*0*^ is stable when _*R*_0_ < 1_ and unstable if *R*_0_ > 1.

Proof. The Jacobian matrix ***J***(***E***^0^) of the model at the vector-free equilibrium point *E*^*0*^ as below:

By applying the characteristic equation to *J*(*E*_0_), we obtain the eigenvalues. If all eigenvalues are real and negative, the vector-free equilibrium is stable, i.e. *R*_0_ < 1otherwise, the endemic equilibrium is stable, i.e. *R*_0_ > 1

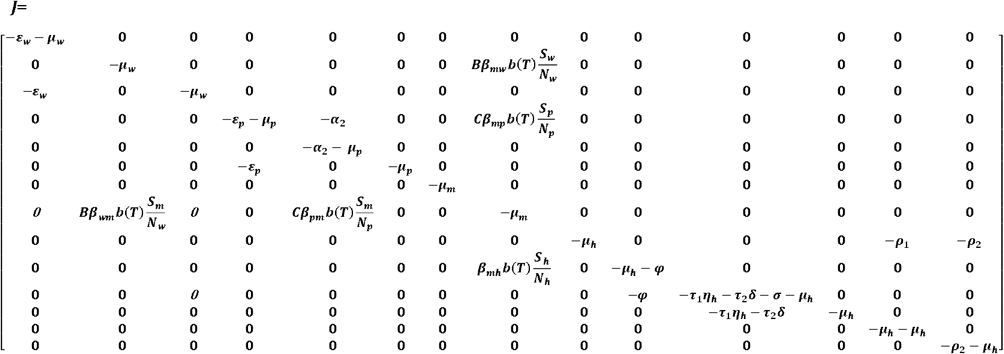

From above, we see that all the characteristic polynomials of the Jacobian matrix are positive; from this, we will get the negative eigenvalues. If *R*_0_ < 1 then the equilibrium point (***E***_0_) is locally asymptotically stable and unstable if *R*_0_ > 1

The detailed proof based on the Jacobian matrix and eigenvalue analysis is provided in Appendix A.

#### 2.6.3 Basic reproduction number

In this section, we estimate the basic reproduction number ***R***_0_, representing the average number of secondary infections caused by a single infected individual in a fully susceptible population. The estimation is carried out using the **next-generation matrix method defined in** (Diekmann et al., 2010). By applying this method to the model equation, we obtained the following results.

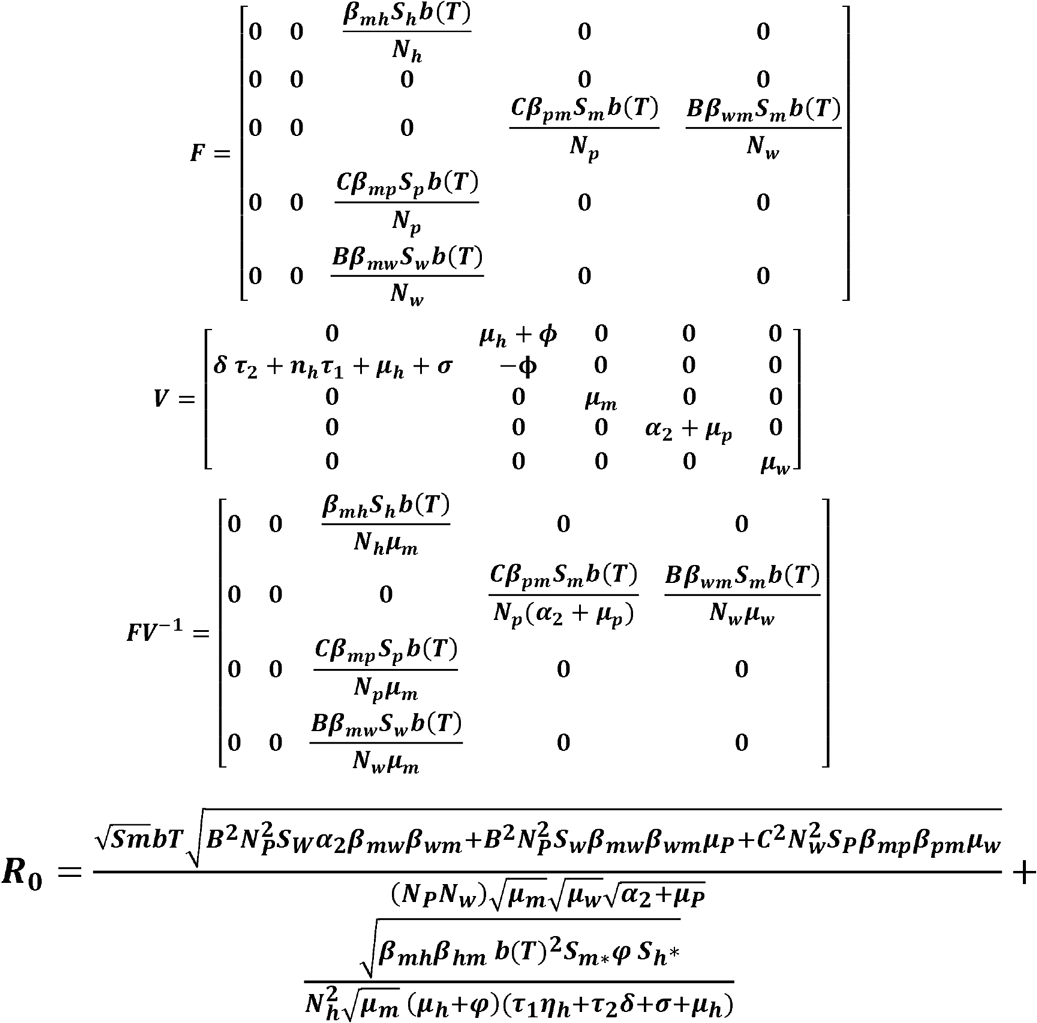

#### 2.6.4 Local sensitivity analysis

***R***_0_ We conducted a sensitivity analysis of the basic reproduction number (R□) to evaluate the impact of key model parameters and identify those that most strongly influence disease transmission. Following the normalized forward-sensitivity approach described by Kouidere et al. (2021), we used the sensitivity index given by:

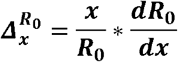

Here, denotes any parameter affecting. This method quantifies how changes in each parameter influence, indicating whether their relationship is positive or negative. For instance, the sensitivity index for the mosquito mortality rate () was calculated, suggesting a negative correlation with, while the transmission parameter implying a positive association. Similar computations were performed for other parameters and are summarized in Table 2.

**Table 2.** Sensitivity index of the parameters.

| Parameter symbol | $\mu_p$ | $\mu_m$ | $\mu_w$ | $\alpha_2$ | $\beta_{pm}$ | $\beta_{mp}$ | $\beta_{wm}$ | $\beta_{mw}$ |
| --- | --- | --- | --- | --- | --- | --- | --- | --- |
| Value | -0.058 | -0.0714 | -0.05 | -0.37 | 0.3 | 0.3 | 0.009 | 0.009 |

### 2.7 Numerical simulation and analytical procedures

#### 2.7.1 District simulation setup

District-level simulations were performed for Bellary and Udupi in Karnataka and Kolkata and Purba Bardhaman in West Bengal over 51 days (days 0–50) during August, the assumed seasonal transmission peak. At 28°C, the temperature-dependent biting rate was b(T) = 0.261 bites per mosquito per day. The seasonal human-transmission contribution was β(t) = 0.0648, giving an effective mosquito-to-human transmission parameter of β_mh = 0.3648. Vaccination-adjusted reproduction-number calculations used protective efficacies of VE□ = 0.60 for dose series I and VE□ = 0.90 for dose series II. The district reproduction-number values and all simulation outputs were retained exactly as generated by the implemented model and dashboard.

#### 2.7.2 Global sensitivity analysis using LHS–PRCC

Global sensitivity analysis was performed for the general reference model rather than separately for each district. Thirty uncertain biological, transmission, demographic, treatment, and ecological parameters were varied simultaneously using Latin hypercube sampling (LHS), following the LHS– PRCC framework described by Marino et al. (2008). Each parameter was sampled independently from a uniform distribution bounded by the lower and upper values reported in Table 3.

**Table 3.** Parameter ranges and LHS–PRCC results for peak total human infections in the general Japanese encephalitis model (500 samples; 500 successful simulations).

| Rank | Parameter | Baseline | LHS range | PRCC | p-value | Significant |
| --- | --- | --- | --- | --- | --- | --- |
| 1 | $bm$ | 0.23 | 0.1–0.4 | +0.833 | 2.16e-122 | Yes |
| 2 | $bT$ | 0.261 | 0.1–0.5 | +0.826 | 5.65e-119 | Yes |
| 3 | $Km$ | 1,000 | 500–5,000 | +0.758 | 6.33e-89 | Yes |
| 4 | $\mu m$ | 0.0714 | 0.03–0.15 | -0.681 | 2.39e-65 | Yes |
| 5 | $rm$ | 0.158 | 0.05–0.3 | -0.673 | 1.69e-63 | Yes |
| 6 | $\beta mh$ | 0.3648 | 0.15–0.7 | +0.584 | 2.49e-44 | Yes |
| 7 | $\varphi$ | 0.1 | 0.03–0.3 | -0.543 | 1.61e-37 | Yes |
| 8 | $\beta_{wm}$ | 0.5 | 0.2–0.8 | +0.301 | 2.52e-11 | Yes |
| 9 | $Brate$ | 0.7 | 0.3–1.2 | +0.290 | 1.34e-10 | Yes |
| 10 | $\beta mp$ | 0.3 | 0.1–0.6 | +0.112 | 0.0148 | Yes |
| 11 | $\delta$ | 0.667 | 0.3–0.95 | -0.105 | 0.0222 | Yes |
| 12 | $\tau 2$ | 0.5 | 0.2–0.8 | -0.085 | 0.0640 | No |
| 13 | $\beta pm$ | 0.3 | 0.1–0.6 | +0.082 | 0.0771 | No |
| 14 | $\epsilon w$ | 0.005 | 0.001–0.03 | -0.077 | 0.0945 | No |
| 15 | $\eta h$ | 0.2 | 0.05–0.5 | -0.066 | 0.1501 | No |
| 16 | $bp$ | 0.00147 | 0.0005–0.005 | -0.063 | 0.1753 | No |
| 17 | $Crate$ | 0.9 | 0.4–1.5 | +0.060 | 0.1918 | No |
| 18 | $bh$ | 0.016 | 0.008–0.03 | -0.056 | 0.2225 | No |
| 19 | $\rho 1$ | 0.00556 | 0.001–0.02 | -0.050 | 0.2792 | No |
| 20 | $\rho 2$ | 0.000913 | 0.0001–0.005 | +0.046 | 0.3226 | No |
| 21 | $Kw$ | 100 | 50–500 | +0.044 | 0.3418 | No |
| 22 | $\alpha 2$ | 0.2 | 0.05–0.5 | -0.036 | 0.4418 | No |
| 23 | $\mu w$ | 0.05 | 0.02–0.1 | +0.033 | 0.4734 | No |
| 24 | $\mu p$ | 0.058 | 0.02–0.1 | +0.031 | 0.4997 | No |
| 25 | $\mu h$ | 0.014 | 0.005–0.025 | -0.028 | 0.5421 | No |
| 26 | $\mu mw$ | 0.5 | 0.2–0.8 | -0.018 | 0.6963 | No |
| 27 | $\epsilon p$ | 0.0083 | 0.001–0.05 | -0.017 | 0.7205 | No |
| 28 | $bw$ | 0.084 | 0.04–0.15 | -0.014 | 0.7624 | No |
| 29 | $\tau 1$ | 0.5 | 0.2–0.8 | -0.013 | 0.7713 | No |
| 30 | $rw$ | 0.79 | 0.3–1.5 | -0.013 | 0.7824 | No |

A total of 500 LHS parameter sets were generated, and the complete 14-compartment ordinary differential equation system was simulated for 51 days for every parameter set. The sole response variable was peak total human infection, defined as max□[I□(t) + I_A□(t)], thereby combining symptomatic and asymptomatic infectious humans at the maximum point of the simulated trajectory. All 500 sampled simulations completed successfully and were retained for analysis.

The sampled parameter values and the corresponding peak total human infection outputs were rank-transformed. A partial rank correlation coefficient (PRCC) was then calculated for each parameter while controlling for the remaining 29 sampled parameters. Two-sided p-values were used to assess statistical significance at p < 0.05. Positive PRCC values indicate that increasing the parameter increases peak total human infection, whereas negative values indicate an inverse association. Parameters were ranked by absolute PRCC magnitude; the sampling ranges and final PRCC estimates are reported in Table 3.

#### 2.7.3 Estimation of the time-varying reproduction number

To characterize the temporal variation in Japanese Encephalitis (JE) transmission during the simulation period, the time-varying effective reproduction number (***R***_***t***_) was estimated using daily incidence data generated by the mathematical model. Unlike the basic reproduction number (***R***_0_), which assumes a completely susceptible population, ***R***_***t***_ quantifies the average number of secondary infections produced by an infectious individual at a given time under prevailing epidemiological conditions. The estimation was performed in **RStudio (version 4.6.1)** using the **EpiEstim** package following the Bayesian framework proposed by Cori et al. (2013). The model incorporated a seasonal transmission modifier, ***β*(*t*) = 0.0648**, to account for the increased transmission intensity during the peak transmission month (August), thereby capturing seasonal fluctuations in vector abundance and mosquito biting activity. The renewal equation approach was employed using simulated daily incidence together with the assumed serial interval distribution of Japanese Encephalitis to estimate ***R***_***t***_ throughout the epidemic. Temporal estimates of ***R***_***t***_ were further smoothed to reduce short-term stochastic fluctuations and facilitate interpretation of transmission dynamics. Values of ***R***_***t***_ > 1 indicate sustained transmission and epidemic growth, whereas ***R***_***t***_ < 1 signifies declining transmission and progression toward outbreak control. The same computational framework can be applied to simulations for any other month by modifying the seasonal transmission parameter and rerunning the R script.

#### 2.7.4 Seasonal forcing

To evaluate the influence of seasonality on JE transmission, a cosine-based seasonal forcing function was externally added to the existing simulation without changing the original model structure or other parameter values:

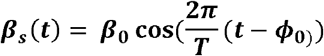

where (***β***_**0**_ =0.0648) is the seasonal amplitude, (T=12) months is the seasonal period, and (***ϕ***_**0**_ = 8) represents the August transmission peak. The seasonal mosquito-to-human transmission rate was calculated as:

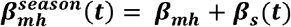

Simulation results with and without seasonal forcing were compared using peak total human infections, peak timing, final infections, and (R_t_) to determine the effect of seasonality on JE transmission.

#### 2.7.5 Intervention scenario analysis

The model incorporates multiple control measures, including human vaccination, fogging to increase mosquito mortality, larviciding to reduce mosquito carrying capacity, pig biosecurity and treatment, physical barriers for wading birds, treatment of infected humans, personal protection to reduce mosquito biting, and enhanced surveillance to improve early detection. Together, these interventions target human susceptibility, mosquito abundance and biting activity, animal–vector contact, and disease recovery, allowing their combined effect on JE transmission to be evaluated.

Baseline trajectories were compared with minimal, moderate, and maximum integrated-control scenarios. Where uncertainty was evaluated, 100-run Monte Carlo simulations were used and the final outcomes were reported as mean ± standard deviation. The intervention settings were applied without altering the underlying district initial conditions or baseline simulation values.

#### 2.7.6 Model validation

Model outputs were compared with annual district surveillance observations for 2008–2020 in Bellary, Udupi, and Kolkata and for 2017–2020 in Purba Bardhaman. Validation was assessed using outbreak-classification accuracy together with absolute and percentage errors in predicted case counts. False-negative district-years were identified explicitly, and the validation was interpreted primarily in relation to outbreak-presence classification and comparative scenario assessment rather than exact case-count forecasting.

#### 2.7.7 Interactive decision-support dashboard

The model was implemented in an interactive simulation dashboard (https://nivedi.res.in/simulation/JE_dashboard.php). The interface allows users to enter farm-level or regional inputs, visualize projected disease trajectories, compare control scenarios, and support district-level interpretation of JE transmission dynamics.

## 3. Results

The district simulations demonstrated marked heterogeneity in transmission thresholds, peak timing, final infection burdens, response to integrated interventions, and validation performance. Results are presented below without modification of the original simulation values.

### 3.1 Bellary District, Karnataka

#### 3.1.1 Compartmental Dynamics and Epidemic Curve

Bellary district was initialized with a human population of N_h = 2,452,595, including 2,250,563 susceptible, 66 symptomatic infectious, 26 asymptomatic infectious, 11 recovered, 107,169 dose-series-I vaccinated, and 94,760 dose-series-II vaccinated individuals. The pig population comprised 15,396 animals (14,935 susceptible, 154 infected, and 307 bio-secured), the wading-bird population comprised 1,005 birds (955 susceptible, 30 infected, and 20 barrier-protected), and the mosquito population was 50,000 (49,700 susceptible and 300 infected) (Figure 4). During the simulation, the combined human infectious burden (I_h + I_Ah) increased from 92 to a maximum of 116 on day 37 and was 114 at day 50 (I_h = 101; I_Ah = 13). Infected pigs declined to 28, infected birds to 25, and infected mosquitoes to 126, producing a final total infected population of 292 (Figure 5). The vaccination-adjusted basic reproduction number was R□ = 0.905, with a herd-immunity threshold of 0.0%, indicating transmission below the self-sustaining threshold. A stable infectious plateau was detected from day 37 (Table 4).

**Table 4.** Summary of Bellary district simulation parameters and epidemic characteristics.

| Parameter | Value |
| --- | --- |
| $R_0$ | 0.905 |
| Herd immunity threshold | 0.0% |
| b(T) biting rate (28°C) | 0.261 |
| Effective $\beta mh$ | 0.3648 |
| Peak human infectious ($I_h + I_{Ah}$) | 116 |
| Peak day | Day 37 |
| $N_h$ (human population) | 2,452,595 |
| $N_p$ (pig population) | 15,396 |
| $N_m$ (mosquito population) | 50,000 |
| Final $I_{total}$ (day 50) | 292 |
| Endemic plateau start | Day 37 |
| JE mortality rate ($\sigma$) | 0 |

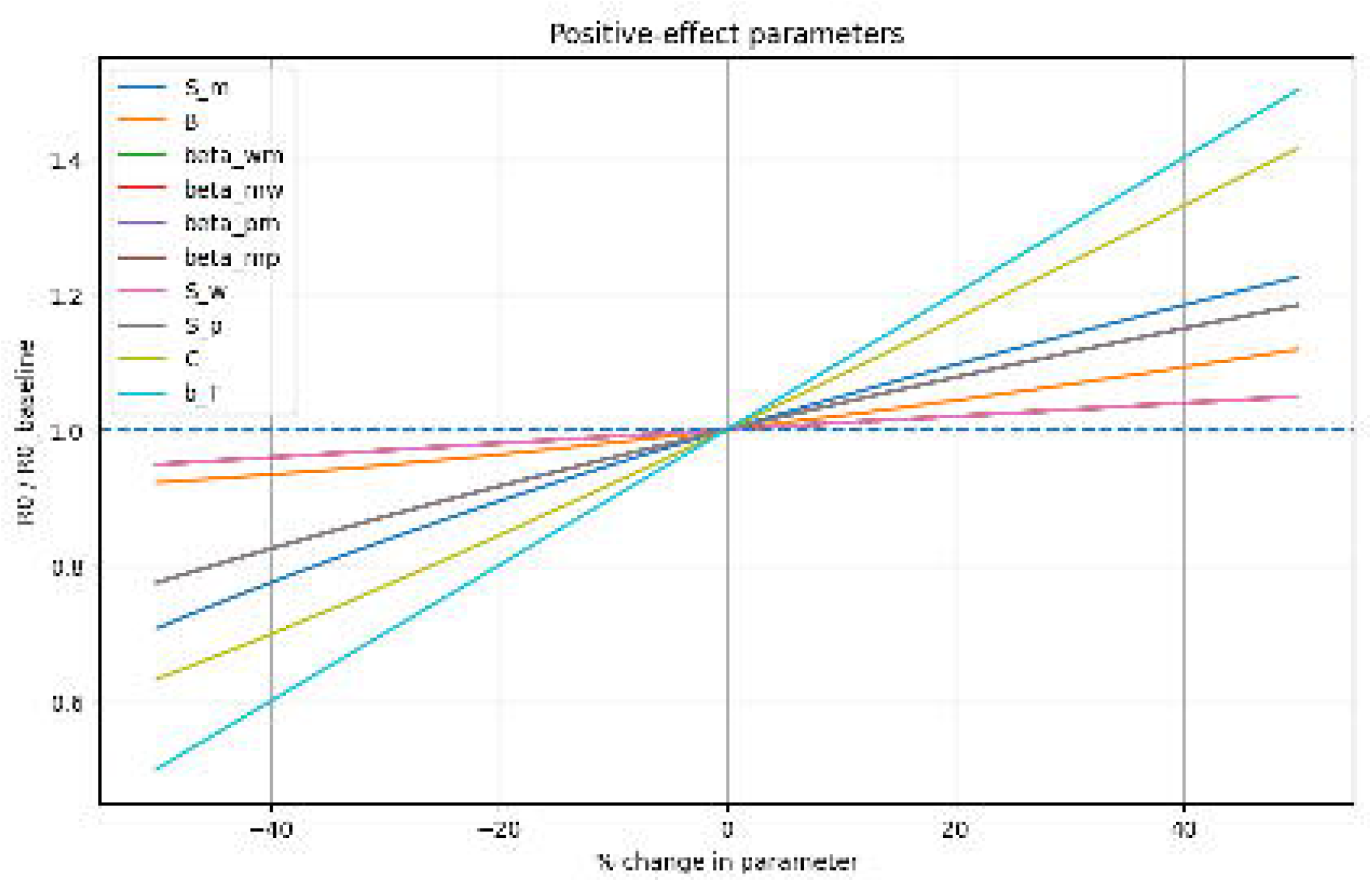

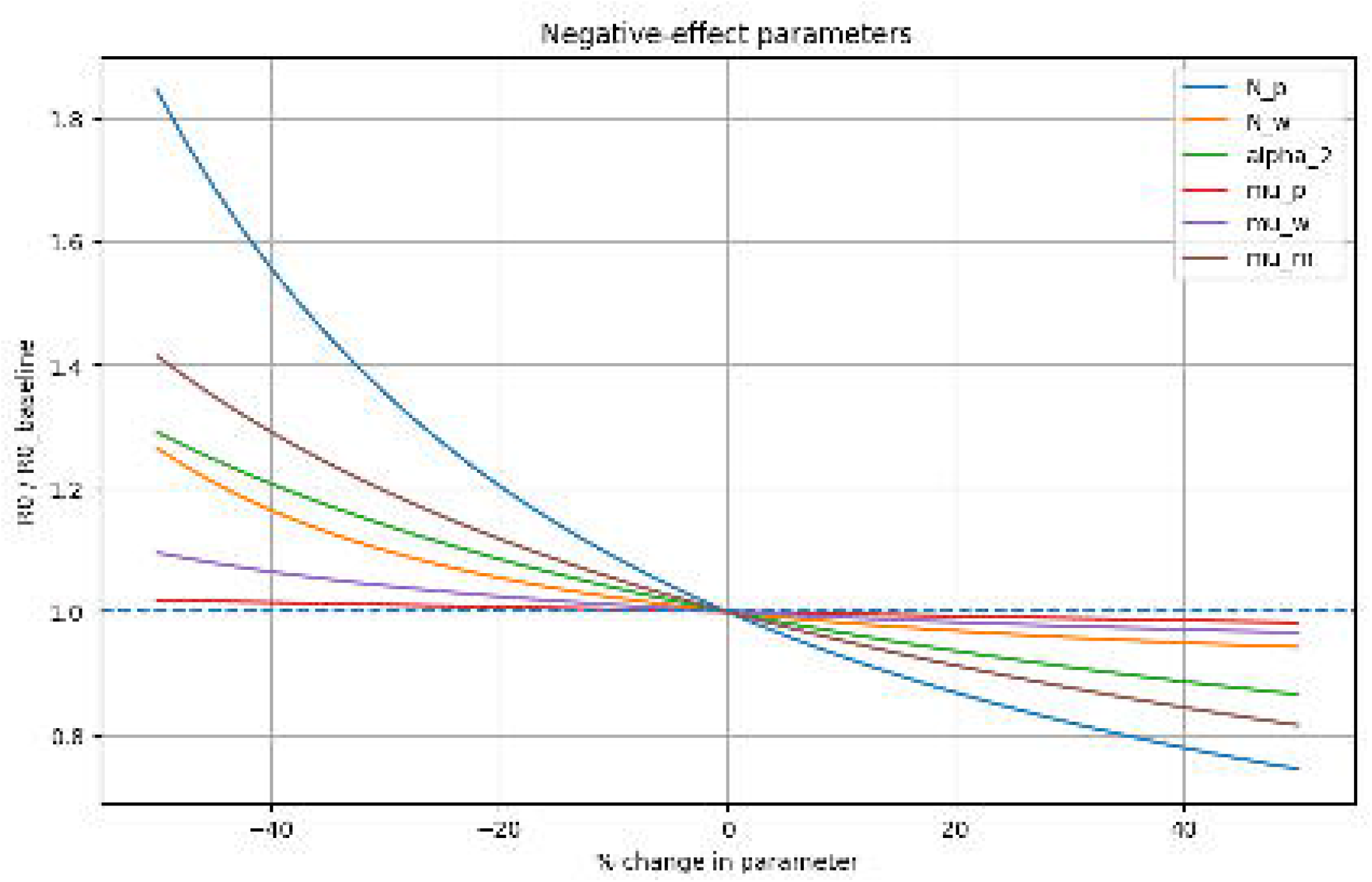

#### 3.1.2 Intervention Impact and Scenario Analysis

Integrated control reduced Bellary’s final total infected population from 292 to 57, corresponding to an 80.5% reduction (Figure 6, Table 5). Final symptomatic human infections decreased from 101 to 47 (53.7%), infected pigs from 28 to 8 (71.7%), infected birds from 25 to 3 (89.9%), and infected mosquitoes from 126 to 0 (100%). The initial total-infection peak of 576 occurred on day 0 and therefore remained unchanged because intervention was introduced later. In the 100-run Monte Carlo analysis, final total infections declined to 159 ± 21 under minimal control (45.5% reduction), 72 ± 5 under moderate control (75.5%), and 57 ± 0 under maximum control (80.6%) (Figure 7). The maximum scenario yielded R_e = 0.4153 and an attack rate of 0.02%. Infection remained above the operational threshold throughout the 51-day simulation in all scenarios.

**Table 5.**
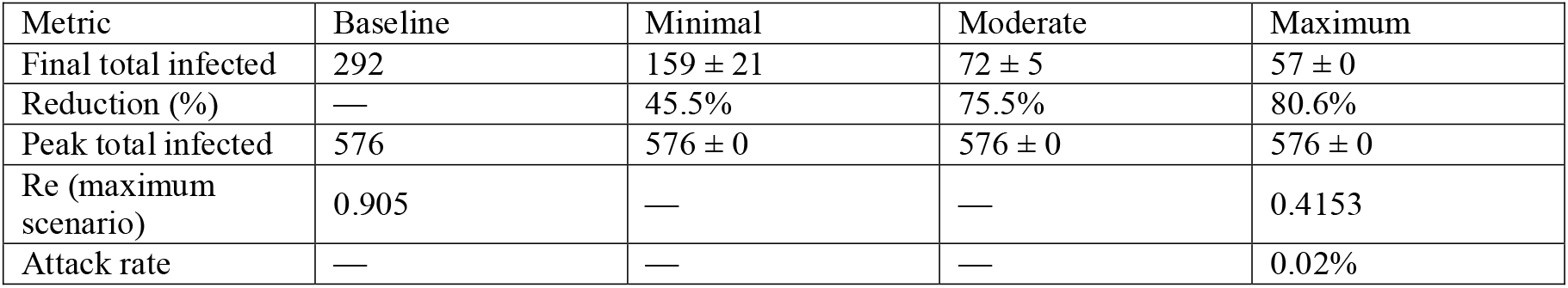
Comparative effectiveness of intervention scenarios in Bellary district.

| Metric | Baseline | Minimal | Moderate | Maximum |
| --- | --- | --- | --- | --- |
| Final total infected | 292 | 159 ± 21 | 72 ± 5 | 57 ± 0 |
| Reduction (%) | — | 45.5% | 75.5% | 80.6% |
| Peak total infected | 576 | 576 ± 0 | 576 ± 0 | 576 ± 0 |
| Re (maximum scenario) | 0.905 | — | — | 0.4153 |
| Attack rate | — | — | — | 0.02% |

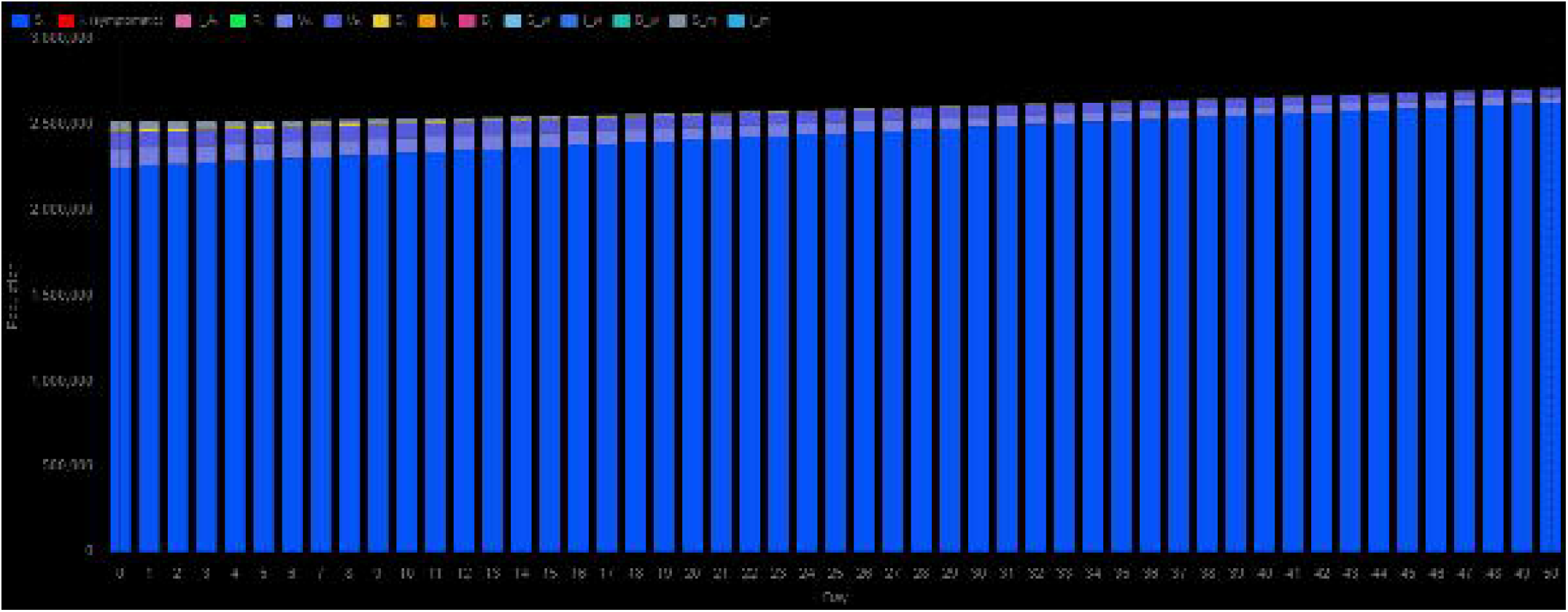

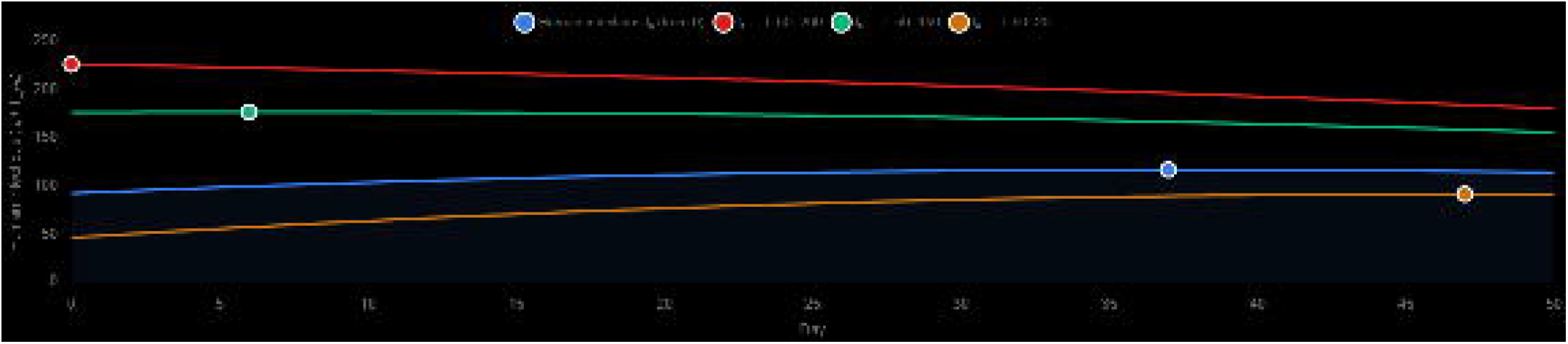

#### 3.1.3 Model Validation

Validation against district surveillance data for 2008–2020 achieved 100% outbreak-classification accuracy (13/13 district-years) (Figure 8, Table 6). All zero-case years were correctly classified. In 2019, 66 cases were observed, and 14 were predicted, giving an absolute error of 52 cases and a percentage error of 78.79%. Thus, the model correctly detected outbreak occurrence but underestimated its magnitude.

**Table 6.** Year-wise validation statistics for Bellary district (2008–2020).

| Year | Observed | Predicted | Classification | % Error |
| --- | --- | --- | --- | --- |
| 2008–2018 (11 years) | 0 each | 0 each | Correct (11/11) | — |
| 2019 | 66 | 14 | Correct | 78.79% |
| 2020 | 0 | 0 | Correct | — |
| Overall accuracy |  |  | 13/13 = 100% |  |

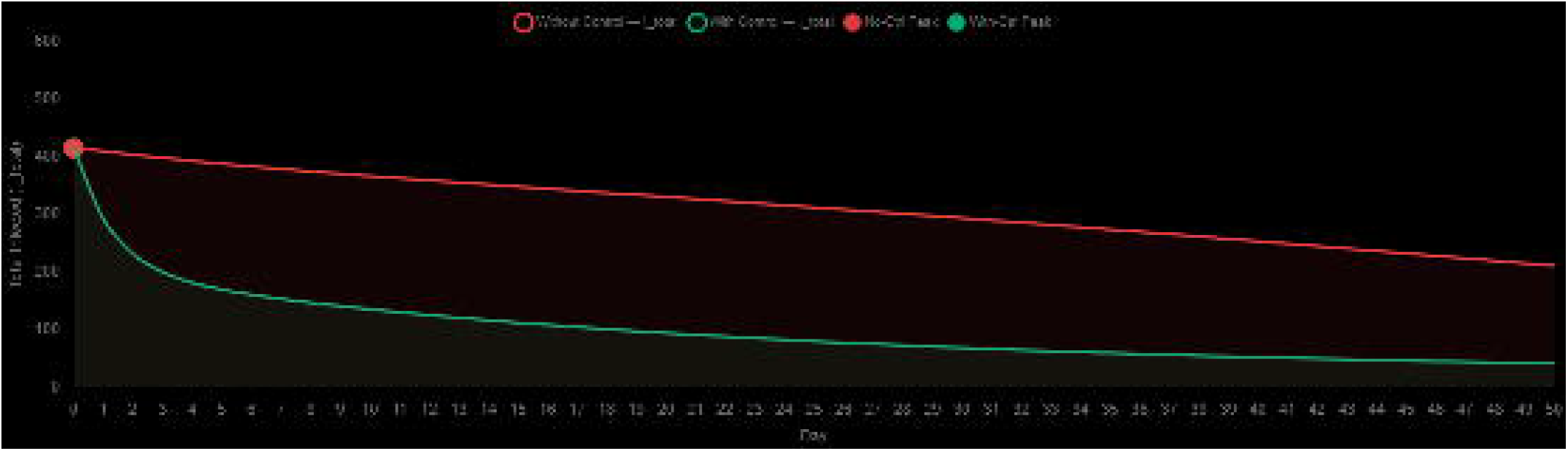

### 3.2 Udupi District, Karnataka

#### 3.2.1 Compartmental Dynamics and Epidemic Curve

Udupi district was initialized with N_h = 1,177,361, including 1,177,289 susceptible, 8 symptomatic infectious, 3 asymptomatic infectious, 1 recovered, 45 dose-series-I vaccinated, and 15 dose-series-II vaccinated individuals. The pig reservoir comprised 3,289 animals (3,190 susceptible, 33 infected, and 66 bio-secured), together with 1,005 wading birds and 50,000 mosquitoes (Figure 9). The combined human infectious burden increased from 11 to 71 on day 48, the latest human peak among the four districts, and remained 71 at day 50 (I_h = 56; I_Ah = 15). Infected pigs declined to 22 and infected mosquitoes to 143, while the final total infected population was 263 (Figure 10). The vaccination-adjusted R□ was 0.965, marginally below unity, indicating a near-threshold system in which modest ecological or transmission changes could permit sustained spread. No stable endemic plateau was detected within the 51-day simulation (Table 7).

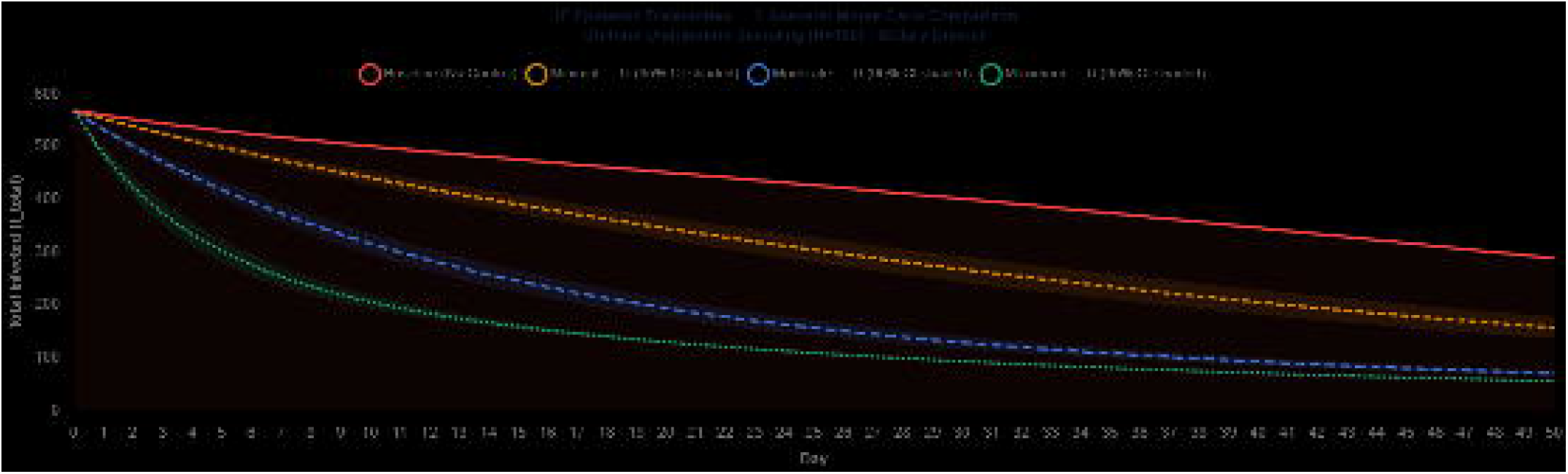

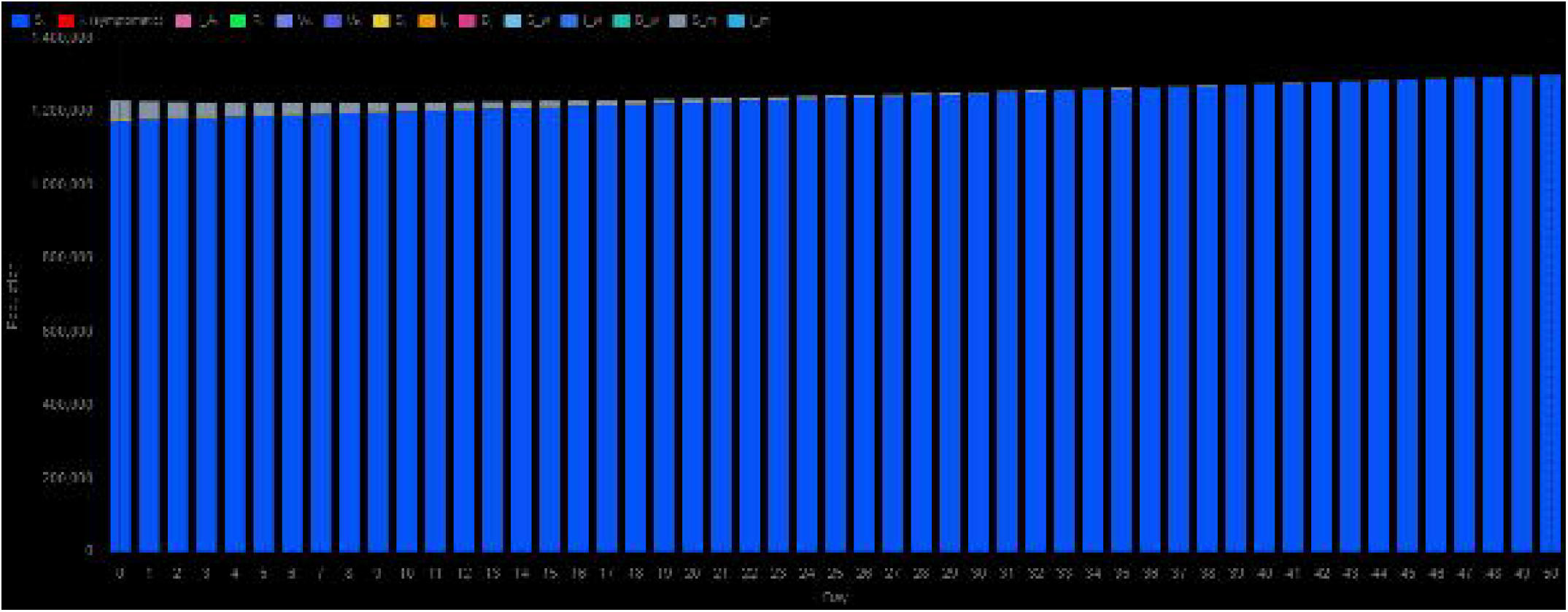

**Table 7.**
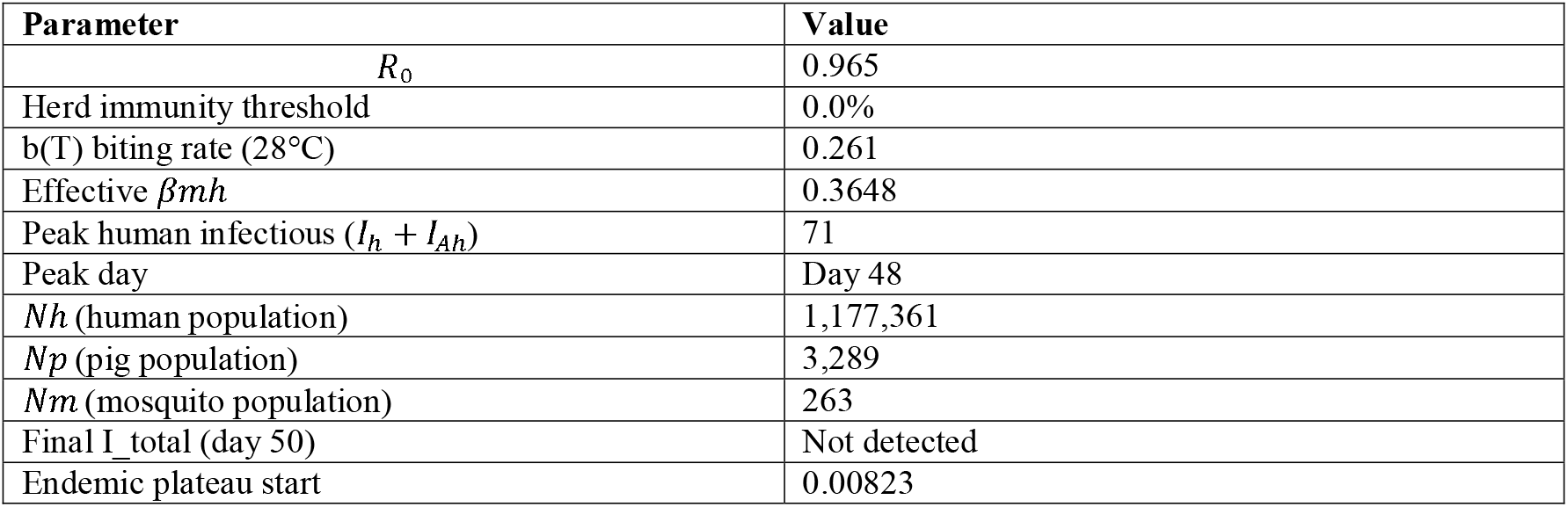
Summary of Udupi district simulation parameters and epidemic characteristics.

#### 3.2.2 Intervention Impact and Scenario Analysis

Udupi showed the greatest response to integrated intervention. The peak total infected population declined slightly from 378 to 374 (1.0%), whereas the final total infected population fell from 263 to 9, a 96.6% reduction (Figure 11, Table 8). Final symptomatic human infections decreased from 56 to 5 (91.9%), infected pigs from 22 to 2 (92.1%), infected birds from 26 to 3 (90.5%), and infected mosquitoes from 143 to 0 (100%). Monte Carlo analysis produced final totals of 108 ± 25 under minimal control (58.9% reduction), 22 ± 5 under moderate control (91.7%), and 9 ± 0 under maximum control (96.8%) (Figure 12). The maximum intervention scenario yielded R_e = 0.4826 and an attack rate of 0.03%. The 37.9-percentage-point difference between minimal and maximum control demonstrates the importance of intervention intensity in this near-threshold district.

**Table 8.** Comparative effectiveness of intervention scenarios in Udupi district.

| Metric | Baseline | Minimal | Moderate | Maximum |
| --- | --- | --- | --- | --- |
| Final total infected | 263 | $108 \pm 25$ | $22 \pm 5$ | $9 \pm 0$ |
| Reduction (%) | — | 58.9% | 91.7% | 96.8% |
| Peak total infected | 378 | $374 \pm 0$ | $374 \pm 0$ | $374 \pm 0$ |
| $R_e$ (maximum scenario) | 0.965 | — | — | 0.4826 |
| Attack rate | — | — | — | 0.03% |

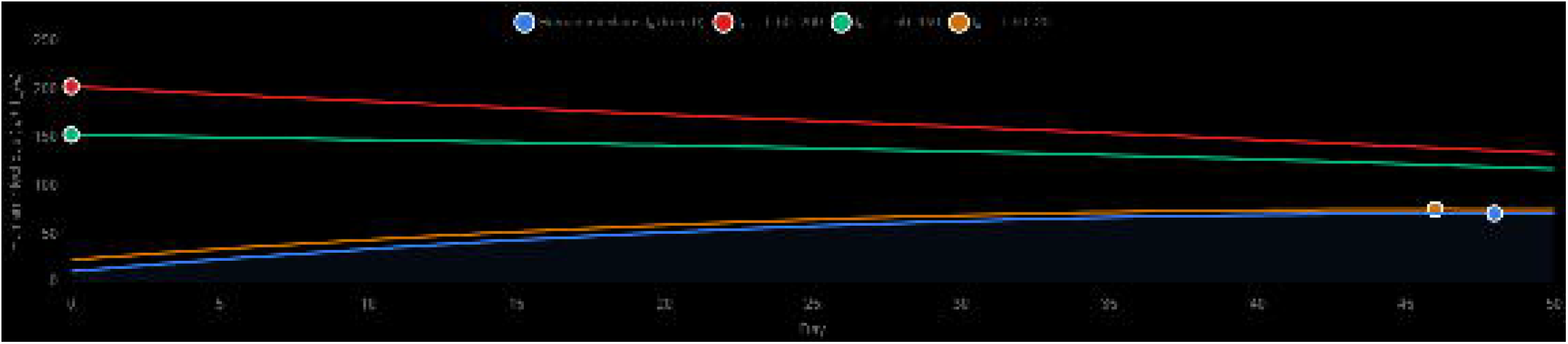

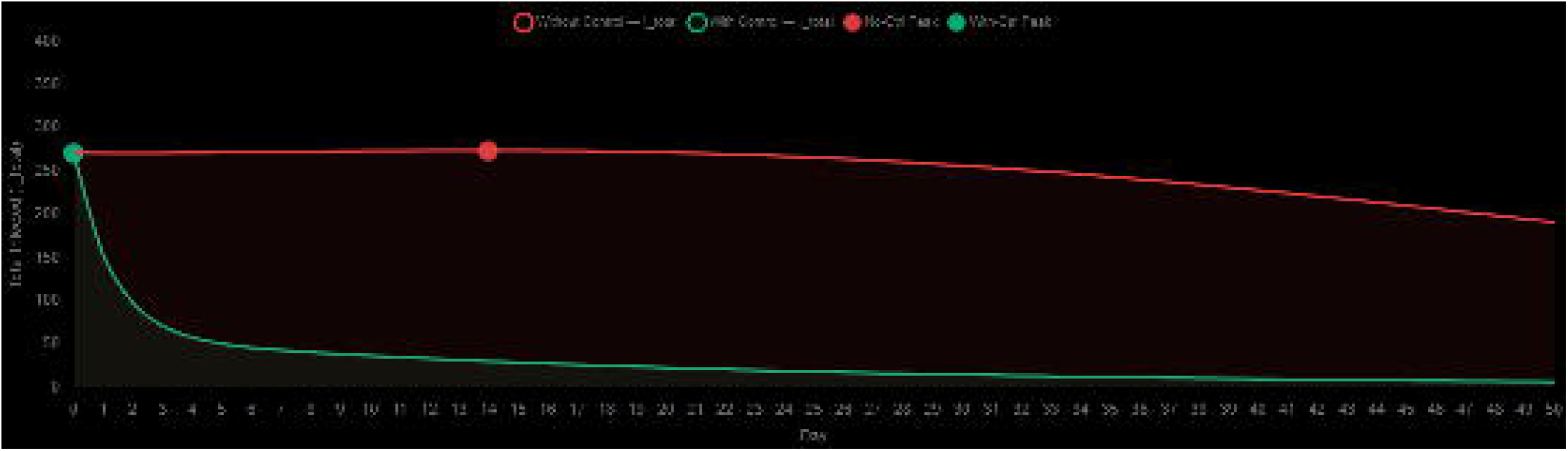

#### 3.2.3 Model Validation

Year-wise validation for 2008–2020 correctly classified 12 of 13 district-years (92.3%) (Figure 13, Table 9). The model correctly classified all nine zero-case years from 2008 to 2016 and the zero-case year 2020. A false-negative classification occurred in 2017, when 2 cases were observed but the model predicted 0. In 2018, 6 cases were observed and 1 was predicted (83.33% error), while in 2019, 3 cases were observed and 1 was predicted (66.67% error). These results indicate good discrimination of quiescent and outbreak years but continued underestimation of low-count outbreaks.

**Table 9.** Year-wise validation statistics for Udupi district (2008–2020).

| Year | Observed | Predicted | Classification | % Error |
| --- | --- | --- | --- | --- |
| 2008–2016 (9 years) | 0 each | 0 each | Correct (9/9) | — |
| 2017 | 2 | 0 | Incorrect | 100.00% |
| 2018 | 6 | 1 | Correct | 83.33% |
| 2019 | 3 | 1 | Correct | 66.67% |
| 2020 | 0 | 0 | Correct | — |
| Overall accuracy |  |  | 12/13 = 92.3% |  |

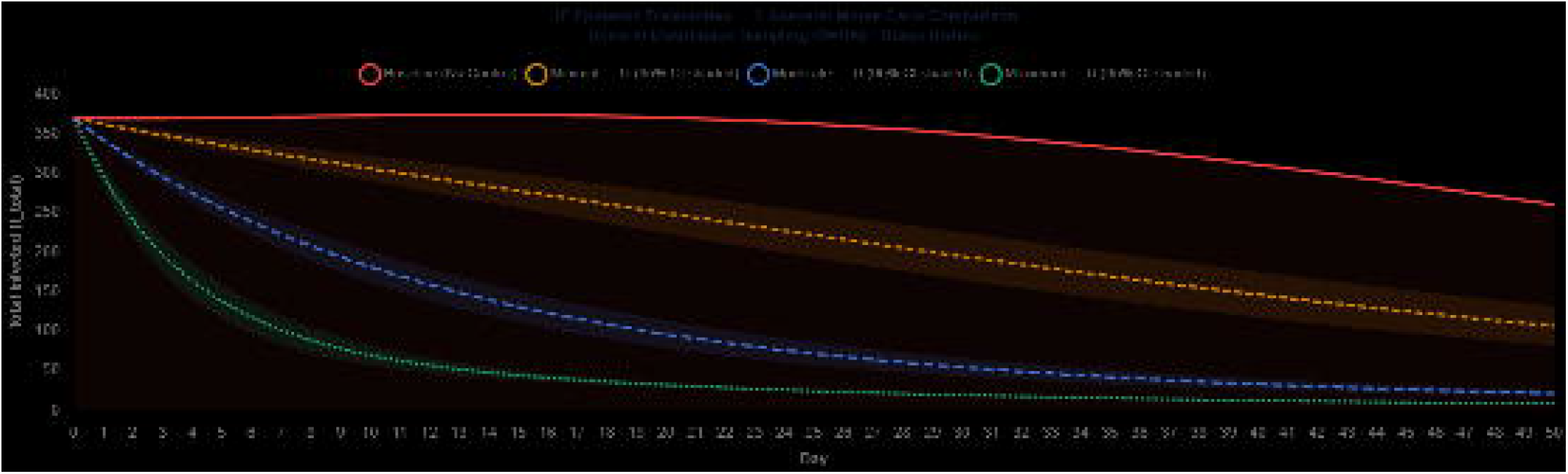

### 3.3 Kolkata District, West Bengal

#### 3.3.1 Compartmental Dynamics and Epidemic Curve

Kolkata was initialized with N_h = 4,496,694, including 4,184,679 susceptible, 207 symptomatic infectious, 83 asymptomatic infectious, 36 recovered, 165,529 dose-series-I vaccinated, and 146,160 dose-series-II vaccinated individuals. The pig reservoir was small (N_p = 335; 3 infected and 7 bio-secured), consistent with a predominantly urban and peri-urban setting (Figure 14). Kolkata was the only district with R□ > 1: the vaccination-adjusted R□ was 1.817, corresponding to a herd-immunity threshold of 45.0%. The combined human infectious burden was already maximal at day 0 (290 individuals) and declined to 231 by day 50 (I_h = 201; I_Ah = 30). In contrast, infected mosquitoes increased from 300 to 313 and infected birds increased from 30 to 38, producing the largest final total infected burden (598) among the four districts (Figure 15). A stable endemic plateau was detected from day 20 (Table 10).

**Table 10.** Summary of Kolkata district simulation parameters and epidemic characteristics.

| Parameter | Value |
| --- | --- |
| $R_0$ | 1.817 |
| Herd immunity threshold | 45.0% |
| b(T) biting rate (28°C) | 0.261 |
| Effective $\beta m h$ | 0.3648 |
| Peak human infectious ($I_h + I_{Ah}$) | 290 |
| Peak day | Day 0 |
| $Nh$ (human population) | 4,496,694 |
| $Np$ (pig population) | 335 |
| $Nm$ (mosquito population) | 598 |
| Final I_total (day 50) | Day 20 |
| Endemic plateau start | 0.00478 |

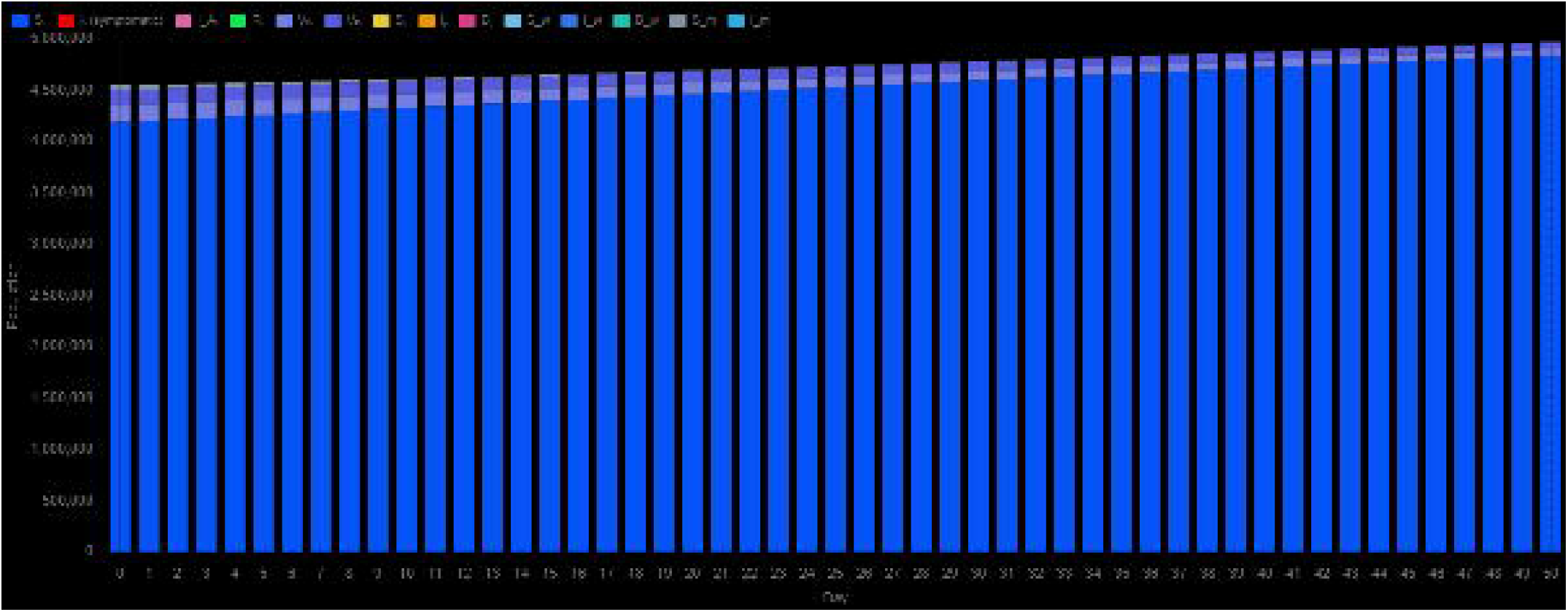

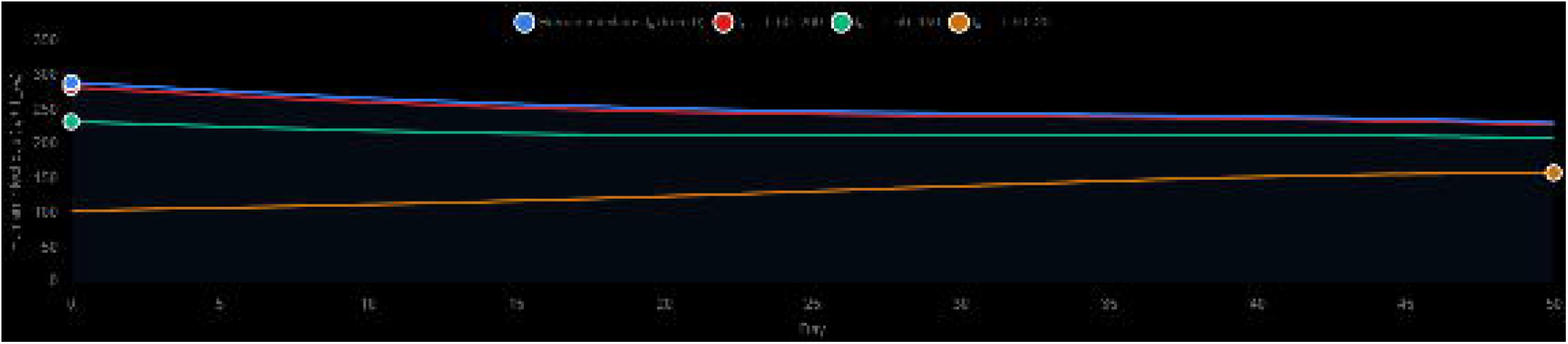

#### 3.2.2 Intervention Impact and Scenario Analysis

Kolkata showed the greatest suppression of the epidemic peak. Integrated control reduced the peak total infected population from 758 to 623 (17.8%) and the final total infected population from 598 to 117 (80.4%) (Figure 16, Table 11). Final symptomatic human infections decreased from 201 to 114 (43.0%), infected pigs from 15 to approximately 0 (98.3%), infected birds from 38 to 3 (93.4%), and infected mosquitoes from 313 to 0 (100%). Monte Carlo analysis yielded final totals of 301 ± 51 under minimal control (49.8% reduction), 134 ± 7 under moderate control (77.6%), and 117 ± 0 under maximum control (80.5%) (Figure 17). The maximum scenario reduced R_e to 0.8453 and produced an attack rate of 0.01%. The 2.9-percentage-point gain from moderate to maximum control indicates diminishing marginal returns at the highest intervention intensity.

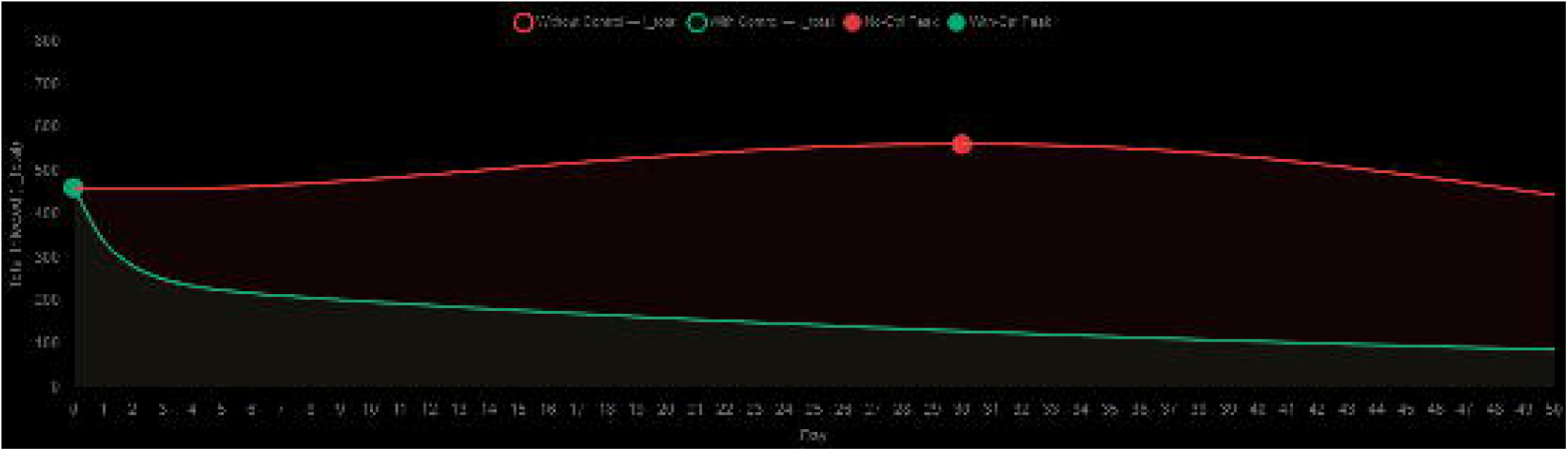

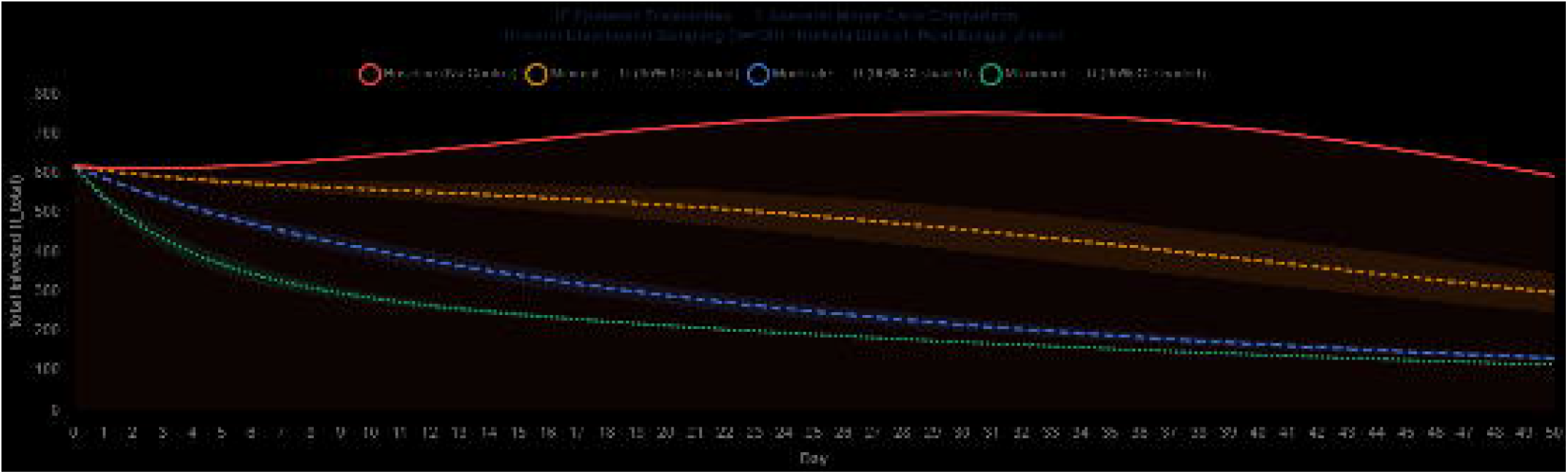

**Table 11.** Comparative effectiveness of intervention scenarios in Kolkata district.

| Metric | Baseline | Minimal | Moderate | Maximum |
| --- | --- | --- | --- | --- |
| Peak total infected | 758 | $623 \pm 0$ | $623 \pm 0$ | $623 \pm 0$ |
| Peak reduction | — | 17.8% | 17.8% | 17.8% |
| Final total infected | 598 | $301 \pm 51$ | $134 \pm 7$ | $117 \pm 0$ |
| Final reduction | — | 49.8% | 77.6% | 80.5% |
| $R_e$ (maximum scenario) | 1.817 | — | — | 0.8453 |

#### 3.3.3 Model Validation

Validation for 2008–2020 achieved 100% classification accuracy (13/13 district-years) (Figure 18, Table 12). The model correctly identified all four outbreak years but underestimated case magnitude: 8 observed versus 2 predicted in 2017 (75.00% error), 84 versus 20 in 2018 (76.19%), 105 versus 25 in 2019 (76.19%), and 10 versus 2 in 2020 (80.00%). JE-associated deaths were recorded in 2017 (6), 2018 (1), and 2019 (2).

**Table 12.**
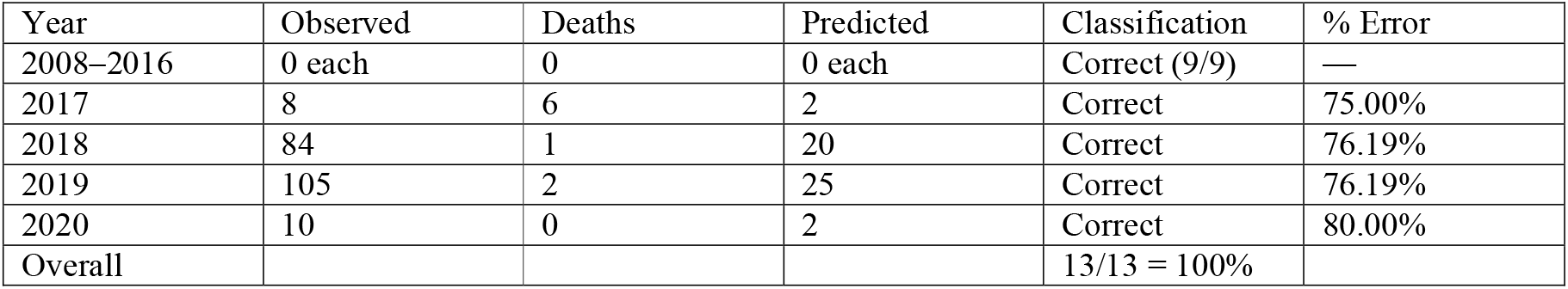
Year-wise validation statistics for Kolkata district (2008–2020).

| Year | Observed | Deaths | Predicted | Classification | % Error |
| --- | --- | --- | --- | --- | --- |
| 2008–2016 | 0 each | 0 | 0 each | Correct (9/9) | — |
| 2017 | 8 | 6 | 2 | Correct | 75.00% |
| 2018 | 84 | 1 | 20 | Correct | 76.19% |
| 2019 | 105 | 2 | 25 | Correct | 76.19% |
| 2020 | 10 | 0 | 2 | Correct | 80.00% |
| Overall |  |  |  | 13/13 = 100% |  |

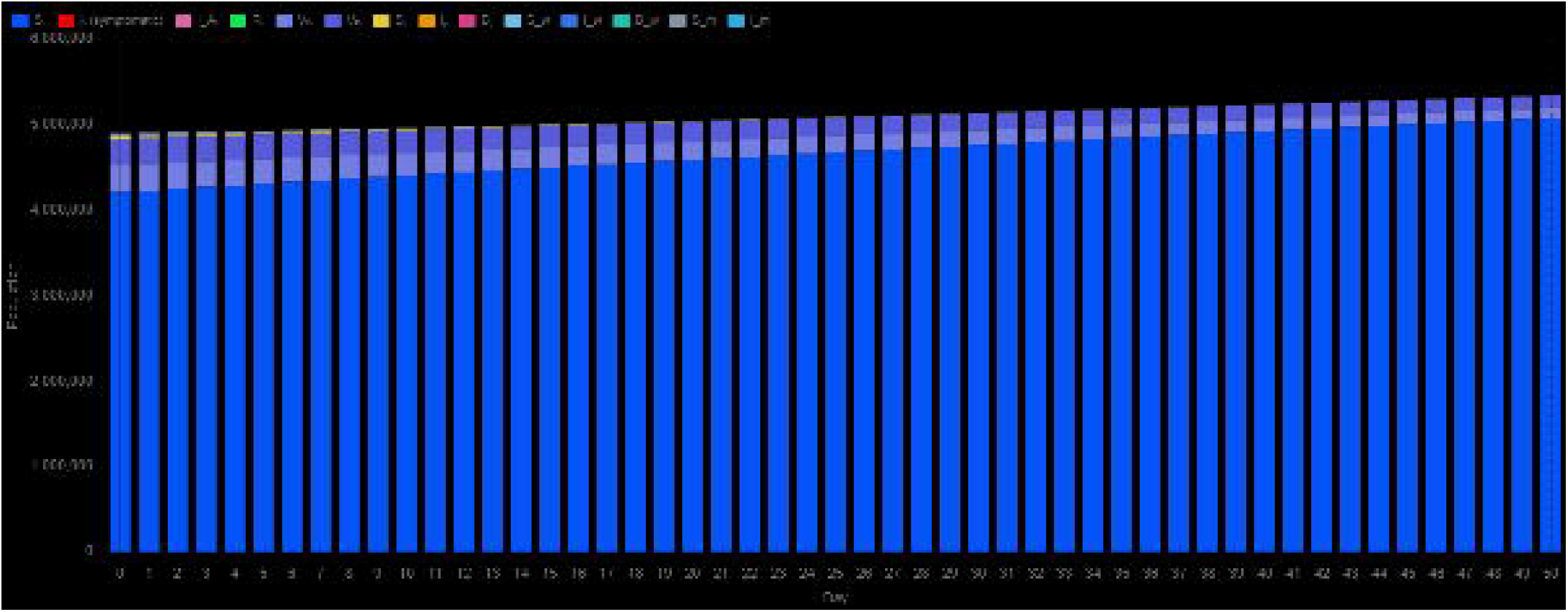

#### 3.3.4 Vaccination Effectiveness

In Kolkata, the baseline (R_0) was 1.817 for a human population of 4,496,694. Vaccinating the entire population with dose series I reduced (R_0) only to 1.767, whereas vaccinating 3.6 million people with dose series II reduced it to 0.9846. A combined strategy of 2.35 million dose-series-I and 2.0 million dose-series-II vaccinations reduced (R_0) to 0.9956, confirming the greater effectiveness of dose series II.

### 3.4 Purba Bardhaman District, West Bengal

#### 3.4.1 Compartmental Dynamics and Epidemic Curve

Purba Bardhaman was initialized with the largest human population (N_h = 4,835,532), including 4,210,766 susceptible, 112 symptomatic infectious, 45 asymptomatic infectious, 19 recovered, 319,666 dose-series-I vaccinated, and 304,924 dose-series-II vaccinated individuals. It also had the largest pig reservoir (N_p = 21,327; 213 infected and 427 bio-secured) (Figure 19). The combined human infectious burden increased from 157 to 158 on day 9 and then declined to 140 by day 50 (I_h = 127; I_Ah = 13). Infected pigs declined to 31 and infected mosquitoes to 124, giving a final total infected population of 320 (Figure 20). The vaccination-adjusted R□ was 0.885, with a herd-immunity threshold of 0.0%. A stable infectious plateau was detected from day 9. The contrast between this sub-threshold rural setting and Kolkata’s supra-threshold transmission highlights the importance of district-specific demography, vaccination, and ecological contact structure (Table 13).

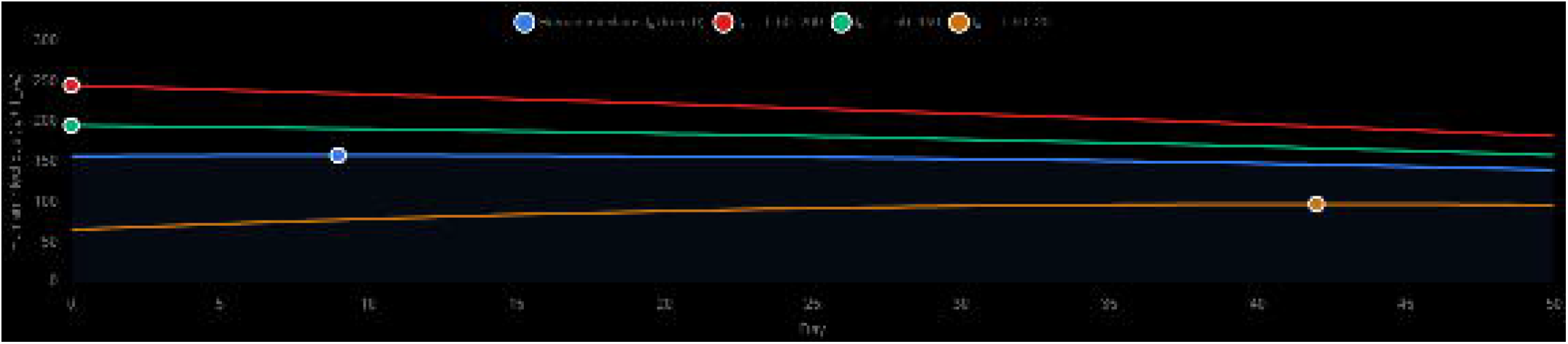

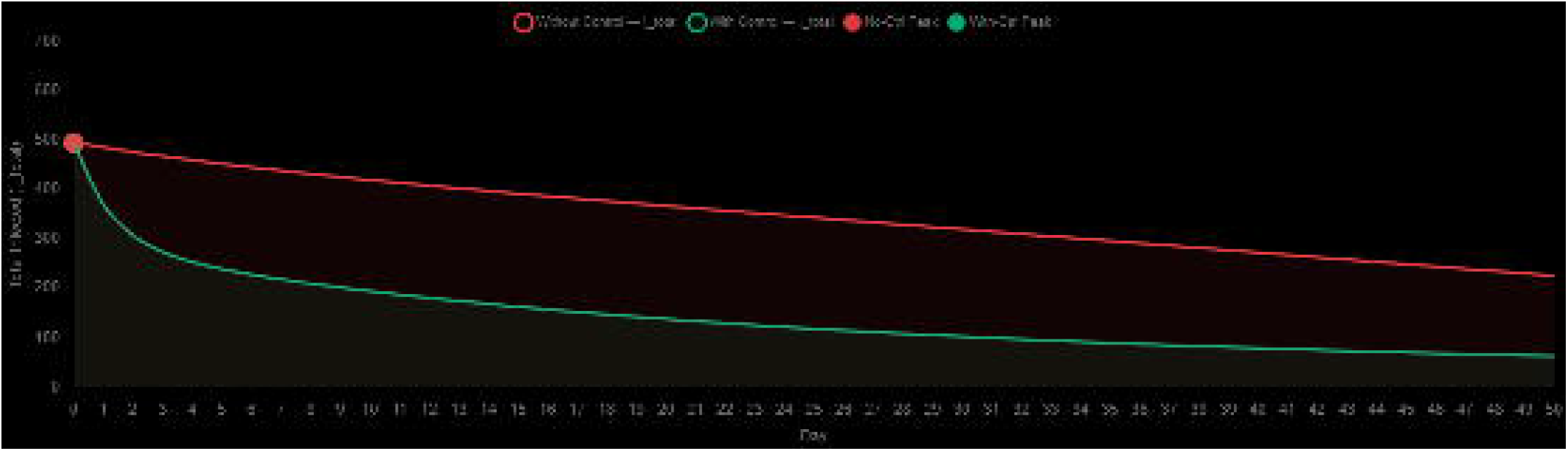

**Table 13.** Summary of Purba Bardhaman district simulation parameters and epidemic characteristics.

| Parameter | Value |
| --- | --- |
| $R_0$ | 0.885 |
| Herd immunity threshold | 0.0% |
| b(T) biting rate (28°C) | 0.261 |
| Effective $\beta mh$ | 0.3648 |
| Peak human infectious ($I_h + I_{Ah}$) | 158 |
| Peak day | Day 9 |
| $N_h$ (human population) | 4,835,532 |
| $N_p$ (pig population) | 21,327 |
| $N_m$ (mosquito population) | 320 |
| Final $I_{total}$ (day 50) | Day 9 |
| Endemic plateau start | 0.000737 |

#### 3.4.2 Intervention Impact and Scenario Analysis

The peak total infected population in Purba Bardhaman remained 700 under both uncontrolled and controlled conditions because it occurred on day 0, before intervention (Figure 21, Table 14). The final total infected population declined from 320 to 89 (72.1%). Final symptomatic human infections decreased from 127 to 76 (40.4%), infected pigs from 31 to 11 (64.7%), infected birds from 25 to 3 (89.9%), and infected mosquitoes from 124 to 0 (100%). Monte Carlo analysis produced final totals of 182 ± 20 under minimal control (43.1% reduction), 103 ± 5 under moderate control (67.9%), and 89 ± 0 under maximum control (72.2%) (Figure 22). The maximum scenario yielded R_e = 0.3852 and an attack rate of 0.01%. The 29.1-percentage-point difference between minimal and maximum control was the smallest among the four districts.

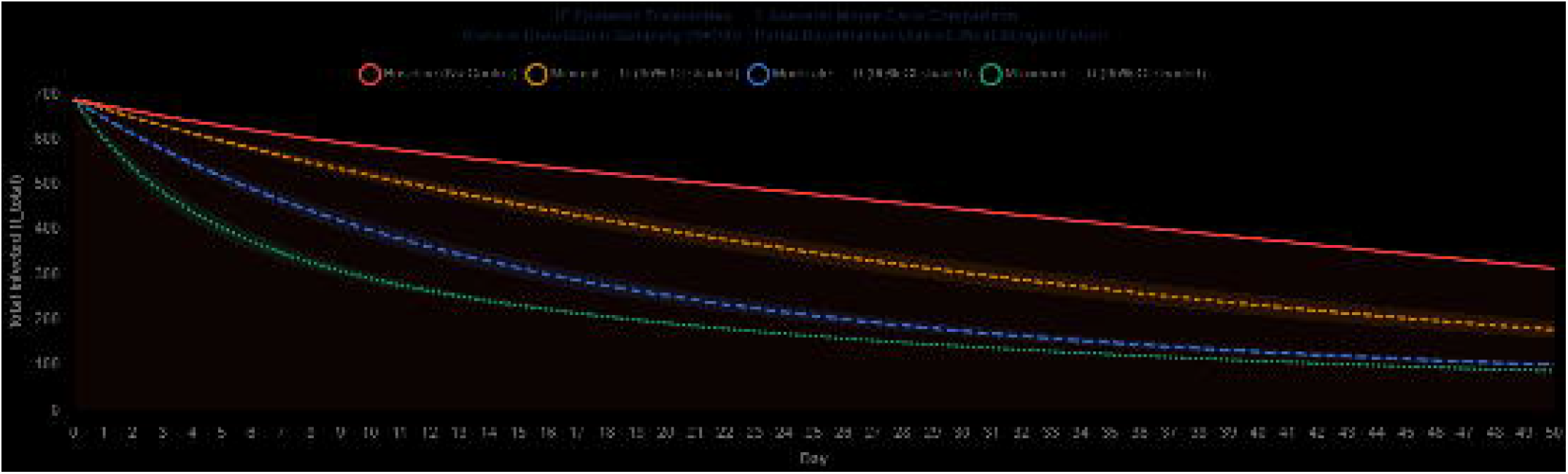

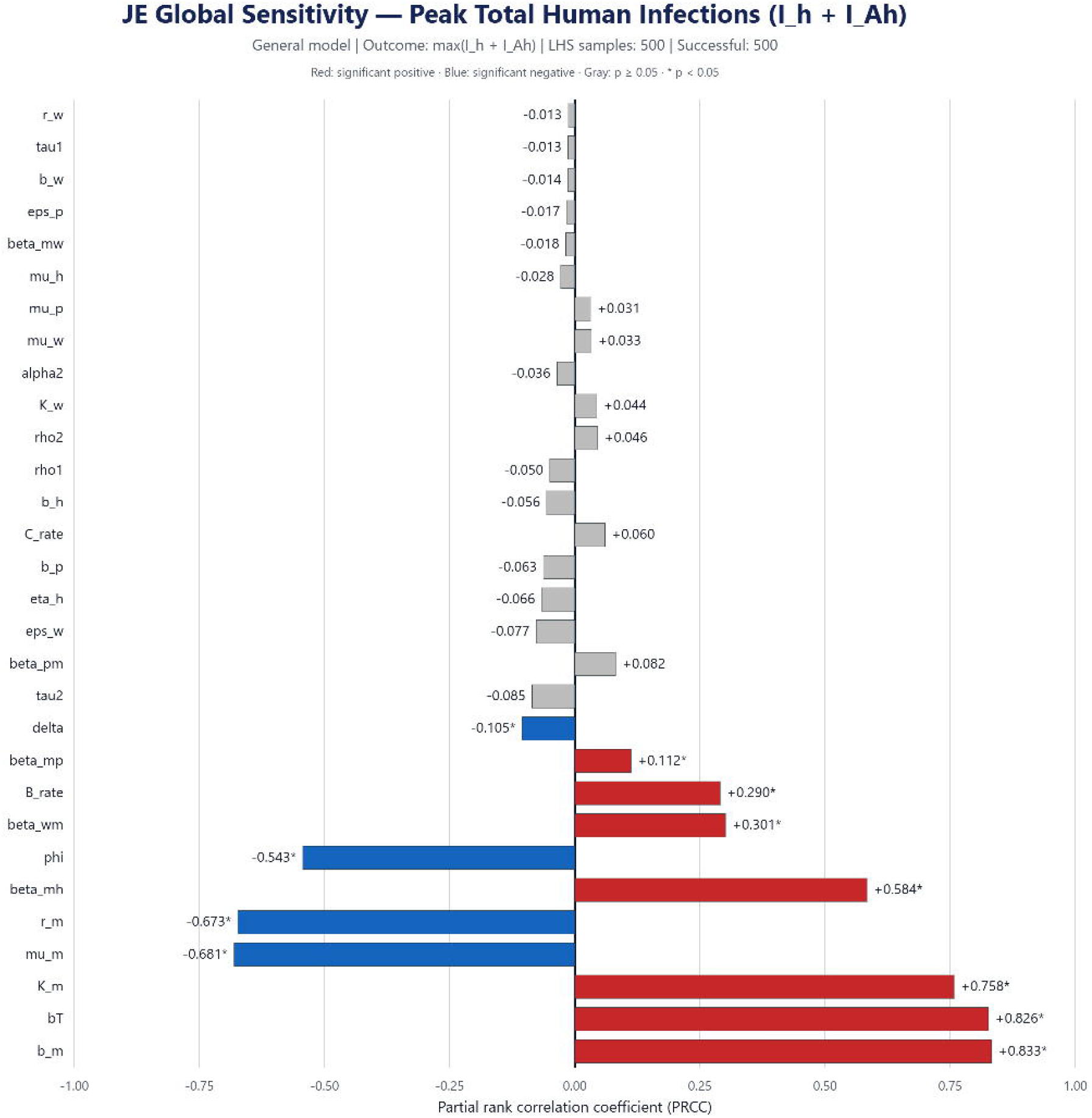

**Table 14.** Comparative effectiveness of intervention scenarios in Purba Bardhaman district.

| Metric | Baseline | Minimal | Moderate | Maximum |
| --- | --- | --- | --- | --- |
| Final total infected | 320 | 182 ± 20 | 103 ± 5 | 89 ± 0 |
| Reduction (%) | — | 43.1% | 67.9% | 72.2% |
| Peak total infected | 700 | 700 ± 0 | 700 ± 0 | 700 ± 0 |
| R<sub>e</sub> (maximum scenario) | 0.885 | — | — | 0.3852 |
| Attack rate | — | — | — | 0.01% |

#### 3.4.3 Model Validation

Validation for 2017–2020 achieved 75% classification accuracy (3/4 district-years) (Figure 23, Table 15). The model correctly detected outbreaks in 2017 (56 observed; 13 predicted; 76.79% error), 2018 (35 observed; 8 predicted; 77.14%), and 2019 (20 observed; 5 predicted; 75.00%). The single-case year 2020 was misclassified because 1 case was observed and 0 was predicted. No JE-associated deaths were reported during the validation period.

**Table 15.**
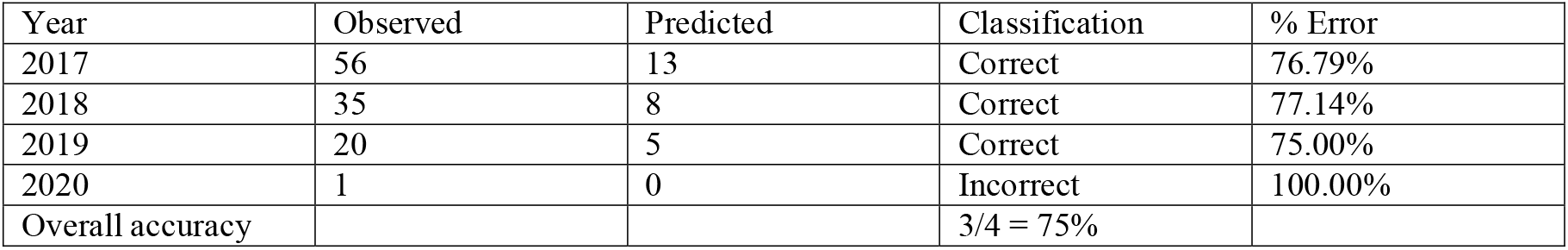
Year-wise validation statistics for Purba Bardhaman district (2017–2020).

### 3.5 Cross-District Comparative Synthesis

Across the four districts, vaccination-adjusted transmission intensity followed the gradient Kolkata (R□ = 1.817) > Udupi (0.965) > Bellary (0.905) > Purba Bardhaman (0.885). Only Kolkata exceeded the epidemic threshold and required an estimated 45.0% immune proportion to interrupt transmission. Human infectious peak timing ranged from day 0 in Kolkata to day 48 in Udupi. Under maximum intervention, final total infections were reduced by 80.6% in Bellary, 96.8% in Udupi, 80.5% in Kolkata, and 72.2% in Purba Bardhaman. Infected mosquitoes were reduced to zero in every district. Udupi was the most intervention-responsive setting, whereas Purba Bardhaman retained the largest proportional residual burden despite a sub-threshold R□.

The model achieved 95.3% accuracy in classifying outbreak occurrence. The two false-negative classifications occurred in Udupi in 2017 (2 observed cases; 0 predicted) and Purba Bardhaman in 2020 (1 observed case; 0 predicted). During outbreak years, percentage error ranged from 66.67% to 100%, indicating systematic underestimation of case magnitude, particularly for low-count events. These findings support the use of the framework for outbreak-presence classification, temporal risk stratification, and comparative intervention assessment rather than for predicting absolute case numbers.

### 3.6 Global Sensitivity Analysis

The general-model LHS–PRCC analysis included 500 parameter sets, and all 500 simulations were completed successfully. Eleven of the 30 evaluated parameters were significantly associated with peak total human infection, defined as max□ [I□(t) + I_A□(t)] (Table 3; Figure 24).

The strongest positive associations were observed for mosquito recruitment rate (b□; PRCC = 0.833, p = 2.16 × 10□^122^), temperature-dependent biting rate [b(T); PRCC = 0.826, p = 5.65 × 10□^11^□], mosquito carrying capacity (K□; PRCC = 0.758, p = 6.33 × 10□□□), and effective mosquito-to-human transmission (β□□; PRCC = 0.584, p = 2.49 × 10□□□). Significant positive effects were also detected for bird-to-mosquito transmission (β_wm; PRCC = 0.301), the average biting rate on birds (B; PRCC = 0.290), and mosquito-to-pig transmission (β_mp; PRCC = 0.112).

The strongest inverse associations were mosquito mortality (μ□; PRCC = −0.681, p = 2.39 × 10□□□), the mosquito density-regulation parameter r□ (PRCC = −0.673, p = 1.69 × 10□□^3^), and progression from asymptomatic to symptomatic human infection (φ; PRCC = −0.543, p = 1.61 × 10□^3^□). Medicine effectiveness also had a smaller but significant negative association (δ; PRCC = −0.105, p = 0.022). The remaining 19 parameters were not statistically significant at p < 0.05. Overall, peak human infection was governed predominantly by mosquito recruitment, biting intensity, environmental carrying capacity, mortality, and mosquito-to-human transmission. The negative association of r□ should be interpreted within the implemented logistic formulation, where increasing r□ strengthens density-dependent suppression of the mosquito population.

## 4. Discussion

### 4.1 Principal findings and concordance with published estimates

Our 14-compartment deterministic multi-host ODE model integrates SVIIR dynamics in humans, SI dynamics in Culex mosquitoes, SISB dynamics in pigs and SIB dynamics in wading birds across four epidemiologically distinct Indian districts. This extends the JEV modelling literature beyond the single-site, two- or three-species frameworks that have predominated (Baniya and Keval, 2020; Solomon, 2000). The model combines advances in livestock-management modelling (Ladreyt et al., 2022), aquatic-environment parameterization (Ndaïrou et al., 2020) and spatiotemporal vector modelling (Franklinos et al., 2022) within one analytically tractable structure, while contributing multi-district empirical validation of a four-population JEV framework in India.

The mathematical analysis confirms that the proposed model satisfies the essential theoretical properties required for epidemiological compartmental systems. The positivity and boundedness analyses ensure biologically feasible solutions, whereas the local and global stability analyses characterize the long-term behaviour of the disease-free and endemic equilibria. Local and global sensitivity analyses further identify the dominant parameters governing disease transmission. The detailed mathematical proofs supporting these results are presented in Appendix A.

The model revealed a vaccination-adjusted transmission gradient with R□ ranging from 0.885 in Purba Bardhaman to 1.817 in Kolkata. Only Kolkata exceeded the epidemic threshold (HIT = 45.0%), while Bellary (0.905), Udupi (0.965) and Purba Bardhaman (0.885) remained below it. The Kolkata estimate is comparable with the R□ range reported by Baniya and Keval (2020) and exceeds the lower range reported for a Cambodian rural multi-host system by Ladreyt et al. (2022) (Table 16). The approximately two-fold district variation emphasizes the need for local parameterization rather than national averaging. Udupi’s near-unity R□ is particularly important because relatively small changes in vector abundance, seasonal contact or host exposure could shift the district above the self-sustaining threshold.

**Table 16.** Comparison of R□ estimates from published JE transmission models and the present district simulations.

| Study | Location | Host System | $R_0$ |
| --- | --- | --- | --- |
| Baniya and Keval (2020) | Nepal | Human–Pig–Mosquito | 1.8–2.4 |
| Ladreyt et al. (2022) | Cambodia | Human–Pig–Mosquito | 1.07–1.38 |
| Present – Kolkata | India | 4-species | 1.817 |
| Present – Udupi | India | 4-species | 0.965 |
| Present – Bellary | India | 4-species | 0.905 |
| Present – Bardhaman | India | 4-species | 0.885 |

### 4.2 Multi-host transmission structure and ecological interpretation

The explicit inclusion of wading birds as a fourth population—beyond the standard human–pig– mosquito framework (Baniya and Keval, 2020; Dwivedi et al., 2024)—reveals transmission-network redundancy: the bird–mosquito cycle can sustain vector infectiousness when pig-targeted interventions reduce the pig–mosquito pathway. This result is consistent with ecological evidence linking JEV risk to wetlands, ardeid birds and domestic pigs (Walsh et al., 2022; Le Flohic et al., 2013), and with findings that pig management alone may be insufficient in multi-host landscapes (Ladreyt et al., 2022).

District variation despite common vector-transmission assumptions highlights the influence of local demography, vaccination, and ecological contact. Kolkata’s supra-threshold R□ occurred despite the smallest pig reservoir (N_p = 335), suggesting an important contribution from bird–mosquito and urban vector-contact pathways. In contrast, Purba Bardhaman remained below threshold (R□ = 0.885) despite the largest pig reservoir, consistent with the moderating effect of its high vaccine coverage and the higher assumed protection of dose series II. The temperature-dependent biting rate was calibrated at T□ = 28°C for the August monsoon setting.

### 4.3 Intervention and vaccination implications

The three-scenario Monte Carlo analysis demonstrated a consistent intervention dose–response with substantial district variation. Udupi was the most responsive district, achieving a 96.8% reduction in final total infections under maximum control, whereas Purba Bardhaman showed the lowest proportional reduction (72.2%). Infected mosquitoes were eliminated in all maximum-control simulations, but residual symptomatic human infections ranged from 5 in Udupi to 114 in Kolkata, indicating that integrated control of human susceptibility, animal amplification, and vector exposure is required. Kolkata showed diminishing returns between moderate and maximum intervention (77.6% versus 80.5%), suggesting that additional locally targeted measures may be needed in high-R□ settings.

### 4.4 Validation performance and intended application

Aggregate validation accuracy for outbreak classification was 95.3%. The two false-negative classifications occurred in low-count years: Udupi in 2017 (2 cases) and Purba Bardhaman in 2020 (1 case). Outbreak-year magnitude errors ranged from 66.67% to 100%, reflecting the limitations of deterministic monthly averaging and uniform annual βmh parameterization in reproducing small case counts. These findings support the model’s application for outbreak detection and comparative risk assessment, whereas prediction of absolute case numbers would require finer temporal calibration and district-specific transmission parameterization.

### 4.5 Strengths and limitations

The principal strengths of the study are the explicit representation of four interacting host–vector populations, district-specific parameterisation, and incorporation of temperature and seasonal forcing. The framework enables simultaneous evaluation of interventions targeting human, animal, vector, and environmental components, together with validation against district-level surveillance data. These features support location-specific assessment of JE transmission and control strategies within a One Health framework.

Several limitations warrant consideration. The deterministic ODE framework cannot represent stochastic extinction or individual-level heterogeneity, particularly for single-case or low-count outbreaks. Spatial homogeneity is assumed within districts, although JE transmission clusters around rice paddies, piggeries, wetlands, and mosquito breeding habitats. The uniform annual β_mh values and monthly averaging contributed to systematic underestimation of outbreak magnitude. Validation was limited to 2008–2020 in Karnataka and 2017–2020 in Purba Bardhaman. Nevertheless, the 95.3% classification accuracy across four ecologically distinct districts supports the model’s utility for outbreak detection and scenario comparison, while not establishing precise case-count forecasting performance.

### 4.6 Policy implications and future directions

Three actionable policy implications emerge. First, integrated multi-domain interventions—combining vaccination, pig biosecurity, bird barriers, and vector reduction—are necessary, as no single-domain strategy achieved sufficient reduction across any district, operationalizing the One Health paradigm for JEV control. Second, district-level R□ heterogeneity demands locally tailored strategies rather than uniform national protocols; the NADRES v2 dashboard provides the operational platform for district-specific projections. Third, Udupi’s near-threshold R□ identifies it as a priority district where moderate-intensity intervention could yield disproportionate impact—a targeting insight invisible to national-level averaging. Future work should pursue sub-monthly temporal resolution, Bayesian parameter estimation, multi-district spatial coupling, cost-effectiveness analysis, and stochastic extensions to characterize elimination thresholds under real-world conditions.

## 5. Conclusion

This study developed a deterministic 14-compartment, multi-host model to characterize Japanese encephalitis transmission across interconnected human, pig, wading bird, and mosquito populations. The mathematical analysis established the well-posedness and dynamic properties of the model through non-negativity, boundedness, equilibrium, stability, optimal control, and sensitivity analyses, as detailed in Appendix A. District-specific simulations demonstrated substantial heterogeneity in transmission potential, with vaccination-adjusted reproduction numbers of 0.905 in Bellary, 0.965 in Udupi, 1.817 in Kolkata, and 0.885 in Purba Bardhaman; among the four study settings, only Kolkata exceeded the epidemic threshold. Maximum integrated control reduced final total infections by 80.6%, 96.8%, 80.5%, and 72.2%, respectively, while infected mosquito populations declined to zero across all four settings. Global sensitivity analysis further identified mosquito recruitment, temperature-dependent biting, mosquito carrying capacity, mosquito mortality, and mosquito-to-human transmission as key determinants of peak human infection. Model validation demonstrated consistency between the simulated transmission patterns and the epidemiological observations used for evaluation, supporting the framework’s application for comparative assessment of JE transmission and intervention strategies. Overall, the findings emphasize the importance of integrated, context-specific control approaches targeting multiple components of the JE transmission cycle. Together with the interactive dashboard, the proposed model provides a One Health framework for evaluating transmission risk and exploring locally appropriate strategies for JE prevention and control.

## Supporting information

Appendix A: Mathematical Proofs and Derivations

## Data Availability

All data produced in the present work are contained in the manuscript. The interactive Japanese encephalitis simulation dashboard is also available online.

https://nivedi.res.in/simulation/je_simulation.php

## Author contributions

Suresh K. P. provided the overall conceptual framework for the mathematical modeling approach and guided its theoretical formulation. Mangala C. D. conceptualized the simulation experiments, performed data extraction, conducted the simulations, and led the manuscript writing. Sudarshan P. V. contributed to the development and structuring of the mathematical model. Devika T. R. developed the interactive simulation dashboard to support visualization and application of the model. Jagadish Hiremath and Sengupta P. P. were involved in reviewing, refining, and editing the manuscript for clarity and scientific accuracy. All authors reviewed and approved the final version of the manuscript.

## Funding

Funding for this study was provided by the Gates Foundation (INV-044445).

## Acknowledgements

The authors acknowledge the National Disease Modelling Consortium, Directorate of Health & Family Welfare Services, Arogya Soudha, Bengaluru, Karnataka, India. The authors also acknowledge the Indian Council of Agricultural Research (ICAR), the Spatial Epidemiology Laboratory, and the Director, ICAR–NIVEDI, for providing the facilities and support required to conduct this work.

## Disclaimer

The findings and opinions expressed in this article are based on the authors’ research and do not necessarily represent the views or official position of the Government.

## Competing interests

All authors declare that they have no competing interests.

## Dashboard availability

The interactive JE simulation dashboard is available at https://nivedi.res.in/simulation/JE_scenario.php

## Notes

### Competing Interest Statement

The authors have declared no competing interest.

