## Appendix A: Mathematical Proofs and Derivations for "Mathematical Modeling of Japanese Encephalitis: Multi-Host Transmission Dynamics and Intervention Strategies"

### Appendix A. Mathematical Theorems and Proofs

**Theorem A1 (Positivity / Non-negativity)**
Apply the positivity (non-negativity) theorem from dynamical systems theory to prove non-negativity.
**Goal:** We demonstrate that if all initial conditions are non-negative, then all state variables remain non-negative for all $t \geq 0$.

**Assumptions:** All parameters are non-negative constants, and all initial conditions are non-negative:
$S_w(t), I_w(t), B_w(t), S_p(t), I_p(t), B_p(t), S_m(t), I_m(t), S_h(t), I_{Ah}(t), I_h(t), R_h(t), V1_h(t), V2_h(t) \geq 0$.

**Proof:** We'll prove this for each differential equation using the standard approach:
**WADING BIRDS**

$$\frac{dS_w}{dt} = \left(b_w - \frac{r_w N_w}{K_w}\right) - B\beta_{mw} b(T) \frac{I_m}{N_w} S_w - \varepsilon_w S_w - \mu_w S_w$$

$$\frac{dS_w}{dt}\Big]_{\substack{t=0 \\ S_w=0}} = \left(b_w - \frac{r_w N_w}{K_w}\right) \geq 0$$

All terms are non-negative, except the last, which is subtractive. Let's assume that a positive value would be zero. The depletion term is proportional to $S_w$, so, $\frac{dS_w}{dt} \geq 0$when $S_w = 0 \rightarrow$ no negative flow out of zero.

$$\frac{dS_w}{dt}\Big]_{\substack{t=0 \\ S_w=0}} = \left(b_w - \frac{r_w N_w}{K_w}\right)$$

$\frac{dI_w}{dt} = B\beta_{mw} b(T) \frac{I_m}{N_w} S_w - \mu_w I_w$. If $I_w$ is 0, the loss term vanishes, production from $S_w \geq 0$, so $\frac{dI_w}{dt} \geq 0$.

$\frac{dB_w}{dt} = \varepsilon_w S_w - \mu_w B_w$ Again, if $B_w$is 0, its depletion stops; $I_w \geq 0$ produces it. So, $B_w(t) \geq 0$

**PIGS**

$$\frac{dS_p}{dt} = b_p - C\beta_{mp} b(T) \frac{I_m}{N_p} S_p - \varepsilon_p S_p - \mu_p S_p + \alpha_2 I_p$$

$$\frac{dS_p}{dt}\Big]_{\substack{t=0 \\ S_p=0}} = b_p + \alpha_2 I_p$$

All terms are non-negative, except the last, which is subtractive. Let's assume that a positive value would be zero. The depletion term is proportional to $S_P$, so, $\frac{dS_P}{dt} \geq 0$when $S_P = 0 \rightarrow$ no negative flow out of zero.

$$\frac{dS_p}{dt}\Big]_{\substack{t=0 \\ S_p=0}} = b_p + \alpha_2 I_p \geq 0$$

$\frac{dI_p}{dt} = C\beta_{mp}b(T)\frac{I_m}{N_p}S_p - \alpha_2 I_p - \mu_p I_p$. If $I_P$ is 0, the loss term vanishes, production from $S_P \geq 0$, so $\frac{dI_P}{dt} \geq 0$.

$\frac{dB_p}{dt} = \varepsilon_p S_p - \mu_p B_p$ Again, if $B_P$ is 0, its depletion stops; $I_P \geq 0$ produces it. So, $B_P(t) \geq 0$

**MOSQUITOES**

$$\frac{dS_m}{dt} = \left(b_m - \frac{r_m N_m}{K_m}\right) - \left(C\beta_{pm}b(T)\frac{I_p}{N_p}S_m + B\beta_{wm}b(T)\frac{I_w}{N_w}S_m\right) - \mu_m S_m$$

$$\frac{dS_m}{dt}\Big]_{\substack{t=0\\S_m=0}} = \left(b_m - \frac{r_m N_m}{K_m}\right) \geq 0$$

All terms are non-negative, except the last, which is subtractive. Let's assume that a positive value would be zero. The depletion term is proportional to $S_m$, so, $\frac{dS_m}{dt} \geq 0$ when $S_m = 0 \rightarrow$ no negative flow out of zero.

$$\frac{dS_m}{dt}\Big]_{\substack{t=0\\S_m=0}} = \left(b_m - \frac{r_m N_m}{K_m}\right)$$

$\frac{dI_m}{dt} = \left(C\beta_{pm}b(T)\frac{I_p}{N_p}S_m + B\beta_{wm}b(T)\frac{I_w}{N_w}S_m\right) - \mu_m I_m$,If $I_m$ is 0, the loss term vanishes, production from $S_m \geq 0$, so $\frac{dI_m}{dt} \geq 0$.

**HUMANS**

$$\frac{dS_h}{dt} = (1-\alpha_1)b_h - \beta_{mh}b(T)\frac{I_m}{N_h}S_h + \rho_1 V1_h + \rho_2 V2_h - \mu_h S_h$$

$$\frac{dS_h}{dt}\Big]_{\substack{t=0\\S_h=0}} = (1-\alpha_1)b_h + \rho_1 V1_h + \rho_2 V2_h \geq 0$$

All terms are non-negative, except the last, which is subtractive. Let's assume that a positive value would be zero. The depletion term is proportional to $S_h$, so, $\frac{dS_h}{dt} \geq 0$when $S_h = 0 \rightarrow$ no negative flow out of zero.

$$\frac{dS_h}{dt}\Big]_{\substack{t=0\\S_h=0}} = (1-\alpha_1)b_h + \rho_1 V1_h + \rho_2 V2_h$$

$\frac{dI_{Ah}}{dt} = \beta_{mh}b(T)\frac{I_m}{N_h}S_h - \mu_h I_{Ah} - \varphi I_{Ah}$ ,If $I_{Ah}$ is 0, the loss term vanishes, production from $S_h \geq 0$, so $\frac{dI_{Ah}}{dt} \geq 0$.

$\frac{dI_h}{dt} = \varphi I_{Ah} - (\tau_1\eta_h + \tau_2\delta)I_h - \sigma I_h - \mu_h I_h$ , If $I_h$ is 0, the loss term vanishes, production from $I_{Ah} \geq 0$, so $\frac{dI_h}{dt} \geq 0$.

$\frac{dR_h}{dt} = (\tau_1\eta_h + \tau_2\delta)I_h - \mu_h R_h$ ,If $R_h$ is 0, the loss term vanishes, production from $I_h \geq 0$, so $\frac{dR_h}{dt} \geq 0$.

$\frac{dV1_h}{dt} = u_1\alpha_1 b_h N_h - \rho_1 V1_h - \mu_h V1_h$ ,If $V1_h$ is 0, the loss term vanishes, production from $R_h \geq 0$, so $\frac{dV1_h}{dt} \geq 0$.

$\frac{dV2_h}{dt} = (1-u_1)\alpha_1 b_h N_h - \rho_2 V2_h - \mu_h V2_h$ ,If $V2_h$ is 0, the loss term vanishes, production from $V1_h \geq 0$, so $\frac{dV2_h}{dt} \geq 0$.

Thus, the system is mathematically and epidemiologically well-posed, with all state variables remaining non-negative.

**Theorem A2 (Boundedness)**

**Boundedness of the Wading birds Population**:
The boundedness proof for the model is based on the provided system of differential equations.
Total **Wading birds** Population $N_w = S_w + I_w + B_w$

$$\frac{dN_w}{dt} = \left(b_w - \frac{r_w N_w}{K_w}\right) - B\beta_{mw} b(T)\frac{I_m}{N_w}S_w - \varepsilon_w S_w - \mu_w S_w + B\beta_{mw} b(T)\frac{I_m}{N_w}S_w - \mu_w I_w + \varepsilon_w S_w - \mu_w B_w$$

$$= \left(b_w - \frac{r_w N_w}{K_w}\right) - \mu_w(S_w + I_w + B_w) = b_w - \frac{r_w N_w}{K_w} - \mu_w N_w$$

Internal transfer terms cancel because population flows between compartments conserve the total population size. The only net sources are the birth or entry terms: $b_w$ and $\mu_w$. Every compartment has a net sink due to natural death at a rate $\mu_w$.

Therefore: $\frac{dN_w}{dt} + \left(\frac{r_w}{K_w} + \mu_w\right)N_w = b_w$, compare this to a standard linear differential inequality to the form $dy/_{dt} + p(t)y = Q(t) \Rightarrow e^{\int p(t)\,dt}$

Multiply the integral factor $e^{\int\left(\frac{r_w}{K_w}+\mu_w\right)dt}$ on both sides

$$e^{\left(\frac{r_w}{K_w}+\mu_w\right)t}\left(\frac{dN_w}{dt} + \left(\frac{r_w}{K_w} + \mu_w\right)N_w\right) = b_w\, e^{-\frac{r_w}{K_w}+\mu_w}$$

$$\frac{dN_w\, e^{\left(\frac{r_w}{K_w}+\mu_w\right)t}}{dt} = b_w\, e^{\left(\frac{r_w}{K_w}+\mu_w\right)t}$$

Integrating on both sides $\int \frac{d\left(N_w\, e^{\left(\frac{r_w}{K_w}+\mu_w\right)t}\right)}{dt} = \int b_w\, e^{\left(\frac{r_w}{K_w}+\mu_w\right)t} \Rightarrow N_w\, e^{\left(\frac{r_w}{K_w}+\mu_w\right)t} = \frac{b_w\, e^{\left(\frac{r_w}{K_w}+\mu_w\right)t}}{\left(\frac{r_w}{K_w}+\mu_w\right)} + C$ ...1

At $(t = 0) \Rightarrow N_w(0) - \frac{b_w}{\left(\frac{r_w}{K_w}+\mu_w\right)} = C$ .........2

Substituting eqn (2) in (1):$N_w\, e^{\left(\frac{r_w}{K_w}+\mu_w\right)t} = \frac{b_w\, e^{\left(\frac{r_w}{K_w}+\mu_w\right)t}}{\left(\frac{r_w}{K_w}+\mu_w\right)} + N_w(0) - \frac{b_w}{\left(\frac{r_w}{K_w}+\mu_w\right)}$ …….. 1

Divide $e^{\left(\frac{r_w}{K_w}+\mu_w\right)t}$ term by both side$N_w = \frac{b_w}{\left(\frac{r_w}{K_w}+\mu_w\right)} + \left(N_w(0) - \frac{b_w}{\left(\frac{r_w}{K_w}+\mu_w\right)}\right) e^{-\left(\frac{r_w}{K_w}+\mu_w\right)t}$

At $(t = \infty) \Rightarrow N_w = \frac{b_w}{\left(\frac{r_w}{K_w}+\mu_w\right)}$

The total wading bird population $N_W(t)$ remains bounded above by a constant $N_W(t) \le \max\left(N_W(0), \frac{b_w}{\left(\frac{r_w}{K_w}+\mu_w\right)}\right)$. This proves that the set $\left(S_w(t), I_w(t), B_w(t)\right)$ is bounded above by $\frac{b_w}{\left(\frac{r_w}{K_w}+\mu_w\right)}$ and bounded below by 0 .i.e. $0 \le N_w(t) \le \frac{b_w}{\left(\frac{r_w}{K_w}+\mu_w\right)}$.

**Boundedness of the Pigs**

The boundedness proof for the model is based on the provided system of differential equations.

Total **Pigs** Population $N_P = S_P + I_P + B_P$

$\frac{dN_P}{dt} = b_p - C\beta_{mp} b(T) \frac{I_m}{N_p} S_p - \varepsilon_p S_p - \mu_p S_p + \alpha_2 I_p + C\beta_{mp} b(T) \frac{I_m}{N_p} S_p - \alpha_2 I_p - \mu_p I_p + \varepsilon_p S_p - \mu_p B_p = b_p - \mu_p(S_p + B_p + I_p) = b_p - \mu_p N_P$

Many internal transfer terms cancel because what leaves one compartment enters another. The only net sources are the birth or entry terms: $b_p$ and $\mu_p$. Every compartment has a net sink due to natural death at a rate $\mu_P$.

Therefore: $\frac{dN_P}{dt} + \mu_p N_P = b_p$ , compare this to a standard linear differential inequality to the form $dy/_{dt} + p(t)y = Q(t) \Rightarrow e^{\int p(t)\,dt}$

Multiply both side by the integral factor $e^{\int \mu_p dt} \Rightarrow e^{\mu_p t}\left(\frac{dN_P}{dt} + \mu_p N_P\right) = e^{\mu_p t} b_p \Rightarrow \frac{d(N_p\, e^{\mu_p t})}{dt} = b_p\, e^{\mu_p t}$

Integrating on both side$\int d(N_p\, e^{\mu_p t}) = \int b_p\, e^{\mu_p t}\, dt \Rightarrow N_p\, e^{\mu_p t} = \frac{b_p\, e^{\mu_p t}}{\mu_p} + C$ …….(1)

At $(t = 0) \Rightarrow N_p(0) - \frac{b_p}{\mu_p} = C$ …………………(2)

Substituting eqn (2) in (1):$N_p\, e^{\mu_p t} = \frac{b_p\, e^{\mu_p t}}{\mu_p} + N_p(0) - \frac{b_p}{\mu_p}$

Dividing $e^{\mu_p t}$ term by both side:$N_p = \frac{b_p}{\mu_p} + e^{-\mu_p t}\left(N_p(0) - \frac{b_p}{\mu_p}\right)$

At $(t = \infty) \Rightarrow N_p = \frac{b_p}{\mu_p}$

The total Pigs population $N_P(t)$ remains bound above by a constant $N_P(t) \le \max\left(N_P(0), \frac{b_p}{\mu_p}\right)$.

This proves that the set $\left(S_p(t), I_p(t), B_p(t)\right)$is bounded above by $\frac{b_p}{\mu_p}$ and bounded below by 0.

i.e. $0 \le N_p(t) \le \frac{b_p}{\mu_p}$.

**Boundedness of the Mosquitoes**

The proof of boundedness for the model is based on the provided system of differential equations.
Total **Mosquitoes** Population $N_m = S_m + I_m$

$$\begin{aligned}\frac{dN_m}{dt} &= b_m - \frac{r_m N_m}{K_m} - \left(C\beta_{pm}b(T)\frac{I_p}{N_p}S_m + B\beta_{wm}b(T)\frac{I_w}{N_w}S_m\right) - \mu_m S_m \\ &\quad + \left(C\beta_{pm}b(T)\frac{I_p}{N_p}S_m + B\beta_{wm}b(T)\frac{I_w}{N_w}S_m\right) - \mu_m I_m \\ &= b_m - \frac{r_m N_m}{K_m} - \left(C\beta_{pm}b(T)\frac{I_p}{N_p}S_m + B\beta_{wm}b(T)\frac{I_w}{N_w}S_m\right) - \mu_m S_m \\ &\quad + \left(C\beta_{pm}b(T)\frac{I_p}{N_p}S_m + B\beta_{wm}b(T)\frac{I_w}{N_w}S_m\right) - \mu_m I_m \\ &= b_m - \frac{r_m N_m}{K_m} - \mu_m S_m - \mu_m I_m = b_m - \frac{r_m N_m}{K_m} - \mu_m(S_m + I_m) \\ &= b_m - \left(\frac{r_m}{K_m} + \mu_m\right)N_m\end{aligned}$$

Many internal transfer terms cancel because what leaves one compartment enters another. The only net sources are the birth or entry terms: $b_m$ and $\mu_m$. Every compartment has a net sink due to natural death at a rate $\mu_m$.
Therefore: $\frac{dN_m}{dt} + \left(\frac{r_m}{K_m} + \mu_m\right)N_m = b_m$ compare this to a standard linear differential inequality to the form $dy/_{dt} + p(t)y = Q(t) \Rightarrow e^{\int p(t)\,dt}$

Multiply both side by the integral factor $e^{\int(\frac{r_m}{K_m}+\mu_m)dt} \Rightarrow e^{\left(\frac{r_m}{K_m}+\mu_m\right)t}\left(\frac{dN_m}{dt} + \left(\frac{r_m}{K_m} + \mu_m\right)N_m\right) = b_m\, e^{-\frac{r_m}{K_m}+\mu_m}$

$\frac{dN_m\, e^{\left(\frac{r_m}{K_m}+\mu_m\right)t}}{dt} = b_m\, e^{\left(\frac{r_m}{K_m}+\mu_m\right)t}$

Integrating on both sides $\int d(N_m\, e^{(\frac{r_m}{K_m}+\mu_m)t}) = \int b_m\, e^{(\frac{r_m}{K_m}+\mu_m)t}\, dt \Rightarrow N_m\, e^{(\frac{r_m}{K_m}+\mu_m)t} = \frac{b_m\, e^{(\frac{r_m}{K_m}+\mu_m)t}}{(\frac{r_m}{K_m}+\mu_m)} + C$ ........ 1

At (t=0): $N_m(0) - \frac{b_m}{(\frac{r_m}{K_m}+\mu_m)} = C$ .........2

Substituting eqn (2) in (1)$\Rightarrow N_m\, e^{(\frac{r_m}{K_m}+\mu_m)t} = \frac{b_m\, e^{(\frac{r_m}{K_m}+\mu_m)t}}{(\frac{r_m}{K_m}+\mu_m)} + N_m(0) - \frac{b_m}{\left(\frac{r_m}{K_m}+\mu_m\right)}$ ........ 1

Divide $e^{(\frac{r_m}{K_m}+\mu_m)t}$ term by both side $\Rightarrow N_m = \frac{b_m}{\left(\frac{r_m}{K_m}+\mu_m\right)} + \left(N_m(0) - \frac{b_m}{\left(\frac{r_m}{K_m}+\mu_m\right)}\right) e^{-\left(\frac{r_m}{K_m}+\mu_m\right)t}$

At $(t=\infty)$: $N_m = \frac{b_m}{\left(\frac{r_m}{K_m}+\mu_m\right)}$

The total Mosquitoes population $N_m(t)$ remains bounded above by a constant $N_m(t) \le \max\left(N_m(0), \frac{b_m}{\left(\frac{r_m}{K_m}+\mu_m\right)}\right)$ This proves that the set $(S_m(t), I_m(t))$ is bounded above by $\frac{b_m}{\left(\frac{r_m}{K_m}+\mu_m\right)}$ and bounded below by 0.i.e. $0 \le N_m(t) \le \frac{b_m}{\left(\frac{r_m}{K_m}+\mu_m\right)}$.

**Boundedness of the Humans**

The proof of boundedness for the model is based on the provided system of differential equations.

Total Human Population $N_h = S_h + I_{Ah} + I_h + R_h + V1_h + V2_h$

Now, compute: $\frac{dN_h}{dt} = \sum$ all RHS terms

$\frac{dN_h}{dt} = b_h - b_h\alpha_1 - \beta_{mh}b(T)\frac{I_m}{N_h}S_h + \rho_1 V1_h + \rho_2 V2_h - \mu_h S_h + \beta_{mh}b(T)\frac{I_m}{N_h}S_h - \mu_h E_h - \varphi I_{Ah} + \varphi I_{Ah} - (\tau_1\eta_h + \tau_2\delta)I_h - \sigma I_h - \mu_h I_h + (\tau_1\eta_h + \tau_2\delta)I_h - \mu_h R_h + u_1\alpha_1 b_h N_h - \rho_1 V1_h - \mu_h V1_h + \alpha_1 b_h - u_1\alpha_1 b_h N_h - \rho_2 V2_h - \mu_h V2_h = b_h - \mu_h S_h - \mu_h E_h - \sigma I_h - \mu_h I_h - \mu_h R_h - \mu_h V1_h - \mu_h V2_h = b_h - \mu_h(S_h + E_h + I_h + R_h + V1_h + V2_h) - \sigma I_h = b_h - \mu_h N_h - \sigma I_h$

Many internal transfer terms cancel because what leaves one compartment enters another. The only net sources are the birth or entry terms: $b_h$ and $\mu_h$. Every compartment has a net sink due to natural death at a rate $\mu_h$. Additionally, disease-induced death occurs only in the $I_h$ class through $\sigma I_h$.

Since disease-induced mortality $\sigma I_h \ge 0$, it follows that $\frac{dN_h}{dt} \le b_h - \mu_h N_h$.

Therefore: $\frac{dN_h}{dt} + \mu_h N_h = b_h$ compare this to a standard linear differential inequality to the form $dy/_{dt} + p(t)y = Q(t) \Rightarrow e^{\int p(t)\,dt}$

Multiply both side by integral factor term $e^{\int \mu_h dt} \Rightarrow e^{\mu_h t}\left(\frac{dN_h}{dt} + \mu_h N_h\right) = e^{\mu_h t} b_h$

$\frac{d(N_h e^{\mu_h t})}{dt} = b_h e^{\mu_h t}$

Integrating on both side $\int d(N_h e^{\mu_h t}) = \int b_h e^{\mu_h t} dt \Rightarrow N_h e^{\mu_h t} = \frac{b_h e^{\mu_h t}}{\mu_h} + C$.............(1)

At $(t=0)$: $N_h(0) - \frac{b_h}{\mu_h} = C$ ......................(2)

Substituting in eqn (2) in (1) $\Rightarrow N_h e^{\mu_h t} = \frac{b_h e^{\mu_h t}}{\mu_h} + N_h(0) - \frac{b_h}{\mu_h}$

Dividing $e^{\mu_h t}$ term by both side $\Rightarrow N_h = \frac{b_h}{\mu_h} + e^{-\mu_h t}(N_h - b_h\,\mu_h)$

At $(t=\infty)$: $N_h = \frac{b_h}{\mu_h}$

The total human's population $N_h(t)$ remains bounded above by a constant $N_h(t) \leq \max(N_h(0), \frac{b_h}{\mu_h}$.This proves that the set $(S_h(t), I_{Ah}(t), I_h(t), R_h(t), V1_h(t), V2_h(t))$ is bounded above by $\frac{b_h}{\mu_h}$and bounded below by 0 i.e. $0 \leq N_h(t) \leq \frac{b_h}{\mu_h}$.

**Conclusion:** Therefore, all state variables of the JE transmission model remain bounded within the biologically feasible region for all. Together with the non-negativity established in Theorem A1, this shows that the model solutions remain biologically meaningful and do not grow without bound.

#### Equilibrium point

##### The vector-free equilibrium point

The vector-free equilibrium (VFE) represents a steady state in which the mosquito population is eliminated.

**for wading birds**

$$I_m = 0, N_m = 0, \frac{dS_w}{dt} = 0, \frac{dI_w}{dt} = 0, \frac{dB_w}{dt} = 0$$

$$S_w^* = \frac{\left(b_w - \frac{r_w N_w}{K_w}\right)}{(\varepsilon_w + \mu_w)}, B_w^* = \frac{\varepsilon_w S_w}{\mu_w}$$

**for pigs**

$$I_m = 0, \frac{dS_p}{dt} = 0, \frac{dI_p}{dt} = 0, \frac{dB_p}{dt} = 0$$

$$S_p^* = \frac{b_p + \alpha_2 I_p}{(\varepsilon_p + \mu_p)}, I_P^* = 0, B_P^* = \frac{\varepsilon_p S_p}{\mu_p}$$

**For Mosquitoes**

$$S_m = 0, I_m = 0, N_m = 0, \frac{dS_m}{dt} = 0, \frac{dI_m}{dt} = 0, \frac{dN_m}{dt} = 0$$

**For Humans**

$$I_m = 0, I_h = 0, I_{Ah} = 0, \frac{dS_h}{dt} = 0, \frac{dI_{Ah}}{dt} = 0, \frac{dI_h}{dt} = 0, \frac{dR_h}{dt} = 0, \frac{dV1_h}{dt} = 0, \frac{dV2_h}{dt} = 0$$

$$S_h^* = \frac{(1-\alpha_1)b_h + \rho_1 V1_h + \rho_2 V2_h}{\mu_h}, R_h^* = 0, V1_h^* = \frac{u_1 \alpha_1 b_h N_h}{\mu_h + \rho_1}, V2_h^* = \frac{(1-u_1)\alpha_1 b_h N_h}{\mu_h + \rho_2}$$

$$\mathbf{E_{VEF}} = \{S_w, I_w, B_w, S_P, I_P, B_P, S_m, I_m, S_h, I_{Ah}, I_h, R_h, V1_h, V2_h\}$$

$$= \left\{ \begin{array}{c} \frac{\left(b_w - \frac{r_w N_w}{K_w}\right)}{(\varepsilon_w + \mu_w)}, 0, \frac{\varepsilon_w S_w}{\mu_w}, \frac{b_p + \alpha_2 I_p}{(\varepsilon_p + \mu_p)}, 0, \frac{\varepsilon_p S_p}{\mu_p}, 0, \\ 0, \frac{(1-\alpha_1)b_h + \rho_1 V1_h + \rho_2 V2_h}{\mu_h}, 0, 0, 0, \frac{u_1 \alpha_1 b_h N_h}{\mu_h + \rho_1}, \frac{(1-u_1)\alpha_1 b_h N_h}{\mu_h + \rho_2} \end{array} \right\}$$

**Conclusion:** Thus, the vector-free equilibrium represents the steady state of the system in the absence of the mosquito population, with (S_m^=I_m^=0), thereby eliminating mosquito-mediated JE transmission.

**Endemic equilibrium**

An endemic equilibrium (EE) is a steady state in which the disease persists in the population (i.e., all infected compartments are non-zero).

A point where the disease is prevalent, i.e., $E \neq I_w(t) \neq I_p(t) \neq I_m(t) \neq I_{Ah}(t) \neq I_h(t) \neq S_w(t) \neq B_w(t) \neq S_p(t) \neq B_p(t) \neq S_m(t) \neq S_h(t) \neq R_h(t) \neq V1_h(t) \neq V2_h(t)$

Let's denote $E_1^* = (S_w^*, I_w^*, B_w^*, S_p^*, I_p^*, B_p^*, S_m^*, I_m^*, S_h^*, I_{Ah}^*, I_h^*, R_h^*, V1_h^*, V2_h^*)$

**for wading birds**

$$\frac{dS_w}{dt} = 0, \frac{dI_w}{dt} = 0, \frac{dB_w}{dt} = 0, \frac{dN_w}{dt} = 0$$

$$S_w^* = \frac{\left(b_w - \frac{r_w N_w}{K_w}\right)}{\left(B\beta_{mw} b(T)\frac{I_m}{N_w} + \varepsilon_w + \mu_w\right)}, I_w^* = \frac{B\beta_{mw} b(T)\frac{I_m}{N_w} S_w}{\mu_w}, B_w^* = \frac{\varepsilon_w S_w}{\mu_w}, N_w^* = \frac{b_w}{\left(\frac{r_w}{K_w} + \mu_w\right)}$$

**for pigs**

$$\frac{dS_p}{dt} = 0, \frac{dI_p}{dt} = 0, \frac{dB_p}{dt} = 0$$

$$S_p^* = \frac{b_p}{C\beta_{mp} b(T)\frac{I_m}{N_p} + \varepsilon_p + \mu_p - \alpha_2}, I_p^* = \frac{C\beta_{mp} b(T)\frac{I_m}{N_p} S_p}{\alpha_2 + \mu_p}, B_p^* = \frac{\varepsilon_p S_p}{\mu_p}$$

**For Mosquitoes**

$$\frac{dS_m}{dt} = 0, \frac{dI_m}{dt} = 0, \frac{dB_m}{dt} = 0, \frac{dN_m}{dt} = 0$$

$$S_m^* = \frac{\left(b_m - \frac{r_m N_m}{K_m}\right)}{\left(C\beta_{pm} b(T)\frac{I_p}{N_p} + B\beta_{wm} b(T)\frac{I_w}{N_w} + \mu_m\right)}, I_m^* = \frac{C\beta_{pm} b(T)\frac{I_p}{N_p} S_m + B\beta_{wm} b(T)\frac{I_w}{N_w} S_m}{\mu_m}, N_m^* = \frac{b_m}{\left(\frac{r_m}{K_m} + \mu_m\right)}$$

**For Humans**

$$\frac{dS_h}{dt} = 0, \frac{dI_{Ah}}{dt} = 0, \frac{dI_h}{dt} = 0, \frac{dR_h}{dt} = 0, \frac{dV1_h}{dt} = 0, \frac{dV2_h}{dt} = 0$$

$$S_h^* = \frac{(\rho_1 V1_h + \rho_2 V2_h + (1-\alpha_1) b_h)}{\beta_{mh} b(T)\frac{I_m}{N_h} + \mu_h}, I_{Ah}^* = \frac{\beta_{mh} b(T)\frac{I_m}{N_h} S_h}{\varphi + \mu_h}, I_h^* = \frac{\varphi I_{Ah}}{(\tau_1 \eta_h + \tau_2 \delta + \sigma + \mu_h)}, R_h^* = \frac{(\tau_1 \eta_h + \tau_2 \delta) I_h}{\mu_h},$$

$$V1_h^* = \frac{u_1 \alpha_1 b_h N_h}{\rho_1 + \mu_h}, V2_h^* = \frac{(1-u_1)\alpha_1 b_h N_h}{\rho_2 + \mu_h}$$

Thus, the endemic equilibrium $E_I^*$ represents a persistent-infection steady state in which the infected compartments remain non-zero and JE transmission is maintained within the host–vector system

$$\mathbf{E_{EFE}} = \{S_w, I_w, B_w, S_p, I_p, B_p, S_m, I_m, S_h, I_{Ah}, I_h, R_h, V1_h, V2_h\}$$

$$= \left\{ \begin{array}{c} \dfrac{\left(b_w - \dfrac{r_w N_w}{K_w}\right)}{\left(B\beta_{mw} b(T)\dfrac{I_m}{N_w} + \varepsilon_w + \mu_w\right)}, \dfrac{B\beta_{mw} b(T)\dfrac{I_m}{N_w} S_w}{\mu_w}, \dfrac{\varepsilon_w S_w}{\mu_w}, \dfrac{b_p}{C\beta_{mp} b(T)\dfrac{I_m}{N_p} + \varepsilon_p + \mu_p - \alpha_2}, \dfrac{C\beta_{mp} b(T)\dfrac{I_m}{N_p} S_p}{\alpha_2 + \mu_p}, \\ \dfrac{\varepsilon_p S_p}{\mu_p}, \dfrac{\left(b_m - \dfrac{r_m N_m}{K_m}\right)}{\left(C\beta_{pm} b(T)\dfrac{I_p}{N_p} + B\beta_{wm} b(T)\dfrac{I_w}{N_w} + \mu_m\right)}, \dfrac{C\beta_{pm} b(T)\dfrac{I_p}{N_p} S_m + B\beta_{wm} b(T)\dfrac{I_w}{N_w} S_m}{\mu_m}, \\ \dfrac{(\rho_1 V1_h + \rho_2 V2_h + (1-\alpha_1) b_h)}{\beta_{mh} b(T)\dfrac{I_m}{N_h} + \mu_h}, \dfrac{\beta_{mh} b(T)\dfrac{I_m}{N_h} S_h}{\varphi + \mu_h}, \dfrac{\varphi I_{Ah}}{(\tau_1\eta_h + \tau_2\delta + \sigma + \mu_h)}, \dfrac{(\tau_1\eta_h + \tau_2\delta) I_h}{\mu_h}, \dfrac{u_1\alpha_1 b_h N_h}{\rho_1 + \mu_h}, \\ \dfrac{(1-u_1)\alpha_1 b_h N_h}{\rho_2 + \mu_h} \end{array} \right\}$$

#### Disease free equilibrium

The Disease-Free Equilibrium (DFE) occurs when the infection is entirely eradicated from the populations, leaving only susceptible compartments.

$$I_m = 0, I_P = 0, I_h = 0, I_w = 0, I_{Ah} = 0$$

At equilibrium, the rates of change are equal to zero. $\dfrac{dS_w}{dt} = 0, \dfrac{dI_w}{dt} = 0, \dfrac{dB_w}{dt} = 0, \dfrac{dN_w}{dt} = 0$

Let $E_0$ be the disease-free equilibrium.

**Wading birds**

$$S_w{}^* = \varepsilon_w + \mu_w, B_w{}^* = \frac{\varepsilon_w S_w}{\mu_w}, N_w{}^* = \frac{b_w}{\frac{r_w}{K_w} + \mu_w},$$

**Pigs**

$$S_P{}^* = \frac{\alpha_2 I_p + b_p}{\varepsilon_p + \mu_p}, B_P{}^* = \frac{\varepsilon_p S_p}{\mu_p}$$

**Mosquitoes**

$$S_m{}^* = \frac{\left(b_m - \dfrac{r_m N_m}{K_m}\right)}{\left(C\beta_{pm} b(T)\dfrac{I_p}{N_p} + B\beta_{wm} b(T)\dfrac{I_w}{N_w} - \mu_m\right)}, N_m{}^* = \frac{b_m}{\left(\dfrac{r_m}{K_m} + \mu_m\right)}$$

**Humans**

$$S_h{}^* = \frac{(1-\alpha_1) b_h + \rho_1 V1_h + \rho_2 V2_h}{\mu_h}, R_h{}^* = \frac{(\tau_1\eta_h + \tau_2\delta) I_h}{\mu_h}, V1_h{}^* = \frac{u_1\alpha_1 b_h N_h}{\mu_h + \rho_1},$$

$$V2_h{}^* = \frac{(1-u_1)\alpha_1 b_h N_h}{\mu_h + \rho_2}$$

Thus, $E_0$ represents the infection-free steady state of the model, characterized by the absence of infection in wading birds, pigs, mosquitoes, and humans, i.e., $I_w^* = I_p^* = I_m^* = I_{Ah}^* = I_h^* = 0$.

Therefore, the disease-free equilibrium, E0 is given as

$E_0 =$
$(S_w(t), I_w(t), B_w(t), S_p(t), I_p(t), B_p(t), S_m(t), I_m(t), S_h(t), I_{Ah}(t), I_h(t), R_h(t), V1_h(t), V2_h(t))$

That is

$$E_0 =$$

$$\left(\begin{array}{c} \varepsilon_w + \mu_w, 0, \frac{\varepsilon_w S_w}{\mu_w}, \frac{\alpha_2 I_p + b_p}{\varepsilon_p + \mu_p}, 0, 0, \frac{\left(b_m - \frac{r_m N_m}{K_m}\right)}{\left(C\beta_{pm} b(T)\frac{I_p}{N_p} + B\beta_{wm} b(T)\frac{I_w}{N_w} - \mu_m\right)}, \\ 0, \frac{(1-\alpha_1)b_h + \rho_1 V1_h + \rho_2 V2_h}{\mu_h}, 0, 0, \frac{(\tau_1\eta_h + \tau_2\delta)I_h}{\mu_h}, \frac{u_1\alpha_1 b_h N_h}{\mu_h + \rho_1}, \frac{(1-u_1)\alpha_1 b_h N_h}{\mu_h + \rho_2} \end{array}\right)$$

**THEOREM A3 (Local Stability)**

To simplify the stability analysis, the eigenvalue structure of the matrix is examined carefully. In particular, isolated negative eigenvalues that do not interact with other matrix components can be treated separately. This allows the matrix to be reduced without affecting the remaining stability conditions. Based on this observation, the following theorem is stated.

1. $\frac{dS_w}{dt} = \left(b_w - \frac{r_w N_w}{K_w}\right) - \varepsilon_w S_w - \mu_w S_w$
2. $\frac{dI_w}{dt} = B\beta_{mw} b(T)\frac{I_m}{N_w} S_w - \mu_w I_w$
3. $\frac{dB_w}{dt} = \varepsilon_w S_w - \mu_w B_w$
4. $\frac{dS_p}{dt} = b_p - C\beta_{mp} b(T)\frac{I_m}{N_p} S_p - \varepsilon_p S_p - \mu_p S_p + \alpha_2 I_p$
5. $\frac{dI_p}{dt} = C\beta_{mp} b(T)\frac{I_m}{N_p} S_p - \alpha_2 I_p - \mu_p I_p$
6. $\frac{dB_p}{dt} = \varepsilon_p S_p - \mu_p B_p$
7. $\frac{dS_m}{dt} = (b_m - \frac{r_m N_m}{K_m}) - \mu_m S_m$
8. $\frac{dI_m}{dt} = \left(C\beta_{pm} b(T)\frac{I_p}{N_p} S_m + B\beta_{wm} b(T)\frac{I_w}{N_w} S_m\right) - \mu_m I_m$
9. $\frac{dS_h}{dt} = (1-\alpha_1)b_h + \rho_1 V1_h + \rho_2 V2_h - \mu_h S_h$
10. $\frac{dI_{Ah}}{dt} = \beta_{mh} b(T)\frac{I_m}{N_h} S_h - \mu_h I_{Ah} \text{-} \varphi I_{Ah}$
11. $\frac{dI_h}{dt} = \varphi I_{Ah} - (\tau_1\eta_h + \tau_2\delta)I_h - \sigma I_h - \mu_h I_h$

12. $\frac{dR_h}{dt} = (\tau_1\eta_h + \tau_2\delta)I_h - \mu_h R_h$

13. $\frac{dV1_h}{dt} = u_1\alpha_1 b_h - \mu_h V1_h - \mu_h V1_h$

14. $\frac{dV2_h}{dt} = (1 - u_1)\alpha_1 b_h - \rho_2 V2_h - \mu_h V2_h$

# F=

$$\begin{bmatrix}
0 & 0 & 0 & 0 & 0 & 0 & 0 & 0 & 0 & 0 & 0 & 0 & 0 & 0 \\
B\beta_{mw}b(T)\frac{I_m}{N_w} & 0 & 0 & 0 & 0 & 0 & 0 & B\beta_{mw}b(T)\frac{S_w}{N_w} & 0 & 0 & 0 & 0 & 0 & 0 \\
0 & 0 & 0 & 0 & 0 & 0 & 0 & 0 & 0 & 0 & 0 & 0 & 0 & 0 \\
0 & 0 & 0 & 0 & 0 & 0 & 0 & 0 & 0 & 0 & 0 & 0 & 0 & 0 \\
0 & 0 & 0 & C\beta_{mp}b(T)\frac{I_m}{N_p} & 0 & 0 & 0 & C\beta_{mp}b(T)\frac{S_p}{N_p} & 0 & 0 & 0 & 0 & 0 & 0 \\
0 & 0 & 0 & 0 & 0 & 0 & 0 & 0 & 0 & 0 & 0 & 0 & 0 & 0 \\
0 & 0 & 0 & 0 & 0 & 0 & 0 & 0 & 0 & 0 & 0 & 0 & 0 & 0 \\
0 & B\beta_{wm}b(T)\frac{S_m}{N_w} & 0 & 0 & C\beta_{pm}b(T)\frac{S_m}{N_p} & 0 & C\beta_{pm}b(T)\frac{I_p}{N_p} + B\beta_{wm}b(T)\frac{I_w}{N_w} & 0 & 0 & 0 & 0 & 0 & 0 & 0 \\
0 & 0 & 0 & 0 & 0 & 0 & 0 & 0 & 0 & 0 & 0 & 0 & 0 & 0 \\
0 & 0 & 0 & 0 & 0 & 0 & 0 & \beta_{mh}b(T)\frac{S_h}{N_h} & \beta_{mh}b(T)\frac{I_m}{N_h} & 0 & 0 & 0 & 0 & 0 \\
0 & 0 & 0 & 0 & 0 & 0 & 0 & 0 & 0 & 0 & 0 & 0 & 0 & 0 \\
0 & 0 & 0 & 0 & 0 & 0 & 0 & 0 & 0 & 0 & 0 & 0 & 0 & 0 \\
0 & 0 & 0 & 0 & 0 & 0 & 0 & 0 & 0 & 0 & 0 & 0 & 0 & 0 \\
0 & 0 & 0 & 0 & 0 & 0 & 0 & 0 & 0 & 0 & 0 & 0 & 0 & 0
\end{bmatrix}$$

# V=

$$\begin{bmatrix}
\varepsilon_w + \mu_w & 0 & 0 & 0 & 0 & 0 & 0 & 0 & 0 & 0 & 0 & 0 & 0 & 0 \\
0 & \mu_w & 0 & 0 & 0 & 0 & 0 & 0 & 0 & 0 & 0 & 0 & 0 & 0 \\
-\varepsilon_w & 0 & \mu_w & 0 & 0 & 0 & 0 & 0 & 0 & 0 & 0 & 0 & 0 & 0 \\
0 & 0 & 0 & \varepsilon_p + \mu_p & -\alpha_2 & 0 & 0 & 0 & 0 & 0 & 0 & 0 & 0 & 0 \\
0 & 0 & 0 & 0 & \alpha_2 + \mu_p & 0 & 0 & 0 & 0 & 0 & 0 & 0 & 0 & 0 \\
0 & 0 & 0 & -\varepsilon_p & 0 & \mu_p & 0 & 0 & 0 & 0 & 0 & 0 & 0 & 0 \\
0 & 0 & 0 & 0 & 0 & 0 & \mu_m & 0 & 0 & 0 & 0 & 0 & 0 & 0 \\
0 & 0 & 0 & 0 & 0 & 0 & 0 & \mu_m & 0 & 0 & 0 & 0 & 0 & 0 \\
0 & 0 & 0 & 0 & 0 & 0 & 0 & 0 & \mu_h & 0 & 0 & 0 & -\rho_1 & -\rho_2 \\
0 & 0 & 0 & 0 & 0 & 0 & 0 & 0 & 0 & \mu_h + \varphi & 0 & 0 & 0 & 0 \\
0 & 0 & 0 & 0 & 0 & 0 & 0 & 0 & 0 & -\varphi & (\tau_1\eta_h + \tau_2\delta) + \sigma + \mu_h & 0 & 0 & 0 \\
0 & 0 & 0 & 0 & 0 & 0 & 0 & 0 & 0 & 0 & -(\tau_1\eta_h + \tau_2\delta) & \mu_h & 0 & 0 \\
0 & 0 & 0 & 0 & 0 & 0 & 0 & 0 & 0 & 0 & 0 & 0 & \mu_h + \mu_h & 0 \\
0 & 0 & 0 & 0 & 0 & 0 & 0 & 0 & 0 & 0 & 0 & 0 & 0 & \rho_2 + \mu_h
\end{bmatrix}$$

# F-V=

$$\begin{bmatrix}
-\varepsilon_w - \mu_w & 0 & 0 & 0 & 0 & 0 & 0 & 0 & 0 & 0 & 0 & 0 & 0 & 0 \\
0 & -\mu_w & 0 & 0 & 0 & 0 & 0 & B\beta_{mw}b(T)\frac{S_w}{N_w} & 0 & 0 & 0 & 0 & 0 & 0 \\
-\varepsilon_w & 0 & -\mu_w & 0 & 0 & 0 & 0 & 0 & 0 & 0 & 0 & 0 & 0 & 0 \\
0 & 0 & 0 & -\varepsilon_p - \mu_p & -\alpha_2 & 0 & 0 & C\beta_{mp}b(T)\frac{S_p}{N_p} & 0 & 0 & 0 & 0 & 0 & 0 \\
0 & 0 & 0 & 0 & -\alpha_2 - \mu_p & 0 & 0 & 0 & 0 & 0 & 0 & 0 & 0 & 0 \\
0 & 0 & 0 & -\varepsilon_p & 0 & -\mu_p & 0 & 0 & 0 & 0 & 0 & 0 & 0 & 0 \\
0 & 0 & 0 & 0 & 0 & 0 & -\mu_m & 0 & 0 & 0 & 0 & 0 & 0 & 0 \\
0 & B\beta_{wm}b(T)\frac{S_m}{N_w} & 0 & 0 & C\beta_{pm}b(T)\frac{S_m}{N_p} & 0 & 0 & -\mu_m & 0 & 0 & 0 & 0 & 0 & 0 \\
0 & 0 & 0 & 0 & 0 & 0 & 0 & 0 & -\mu_h & 0 & 0 & 0 & -\rho_1 & -\rho_2 \\
0 & 0 & 0 & 0 & 0 & 0 & 0 & \beta_{mh}b(T)\frac{S_h}{N_h} & 0 & -\mu_h - \varphi & 0 & 0 & 0 & 0 \\
0 & 0 & 0 & 0 & 0 & 0 & 0 & 0 & 0 & -\varphi & -\tau_1\eta_h - \tau_2\delta - \sigma - \mu_h & 0 & 0 & 0 \\
0 & 0 & 0 & 0 & 0 & 0 & 0 & 0 & 0 & 0 & -\tau_1\eta_h - \tau_2\delta & -\mu_h & 0 & 0 \\
0 & 0 & 0 & 0 & 0 & 0 & 0 & 0 & 0 & 0 & 0 & 0 & -\mu_h - \mu_h & 0 \\
0 & 0 & 0 & 0 & 0 & 0 & 0 & 0 & 0 & 0 & 0 & 0 & 0 & -\rho_2 - \mu_h
\end{bmatrix}$$

For stability analysis, the negative eigenvalue is removed from the matrix when it is the only non $-$ zero value present in its corresponding row or column $- \mu_w$

$$(-\varepsilon_w - \mu_w)(-\mu_w)$$

$$\begin{bmatrix}
-\varepsilon_p - \mu_p & -\alpha_2 & 0 & 0 & C\beta_{mp} b(T)\frac{S_p}{N_p} & 0 & 0 & 0 & 0 & 0 & 0 \\
0 & -\alpha_2 - \mu_p & 0 & 0 & 0 & 0 & 0 & 0 & 0 & 0 & 0 \\
-\varepsilon_p & 0 & -\mu_p & 0 & 0 & 0 & 0 & 0 & 0 & 0 & 0 \\
0 & 0 & 0 & -\mu_m & 0 & 0 & 0 & 0 & 0 & 0 & 0 \\
0 & C\beta_{pm} b(T)\frac{S_m}{N_p} & 0 & 0 & -\mu_m & 0 & 0 & 0 & 0 & 0 & 0 \\
0 & 0 & 0 & 0 & 0 & -\mu_h & 0 & 0 & 0 & -\rho_1 & -\rho_2 \\
0 & 0 & 0 & 0 & \beta_{mh} b(T)\frac{S_h}{N_h} & 0 & -\mu_h - \varphi & 0 & 0 & 0 & 0 \\
0 & 0 & 0 & 0 & 0 & 0 & -\varphi & -\tau_1\eta_h - \tau_2\delta - \sigma - \mu_h & 0 & 0 & 0 \\
0 & 0 & 0 & 0 & 0 & 0 & 0 & -\tau_1\eta_h - \tau_2\delta & -\mu_h & 0 & 0 \\
0 & 0 & 0 & 0 & 0 & 0 & 0 & 0 & 0 & -\mu_h - \mu_h & 0 \\
0 & 0 & 0 & 0 & 0 & 0 & 0 & 0 & 0 & 0 & -\rho_2 - \mu_h
\end{bmatrix}$$

$$-\mu_p$$

$$(-\varepsilon_p - \mu_p)(-\alpha_2 - \mu_p)$$

$$\begin{bmatrix}
-\mu_m & 0 & 0 & 0 & 0 & 0 & 0 & 0 \\
0 & -\mu_m & 0 & 0 & 0 & 0 & 0 & 0 \\
0 & 0 & -\mu_h & 0 & 0 & 0 & -\rho_1 & -\rho_2 \\
0 & \beta_{mh} b(T)\frac{S_h}{N_h} & 0 & -\mu_h - \varphi & 0 & 0 & 0 & 0 \\
0 & 0 & 0 & -\varphi & -\tau_1\eta_h - \tau_2\delta - \sigma - \mu_h & 0 & 0 & 0 \\
0 & 0 & 0 & 0 & -\tau_1\eta_h - \tau_2\delta & -\mu_h & 0 & 0 \\
0 & 0 & 0 & 0 & 0 & 0 & -\mu_h - \mu_h & 0 \\
0 & 0 & 0 & 0 & 0 & 0 & 0 & -\rho_2 - \mu_h
\end{bmatrix}$$

$$-\mu_m$$

$$\begin{bmatrix}
-\mu_m & 0 & 0 & 0 & 0 & 0 & 0 \\
0 & -\mu_h & 0 & 0 & 0 & -\rho_1 & -\rho_2 \\
\beta_{mh} b(T)\frac{S_h}{N_h} & 0 & -\mu_h - \varphi & 0 & 0 & 0 & 0 \\
0 & 0 & -\varphi & -\tau_1\eta_h - \tau_2\delta - \sigma - \mu_h & 0 & 0 & 0 \\
0 & 0 & 0 & -\tau_1\eta_h - \tau_2\delta & -\mu_h & 0 & 0 \\
0 & 0 & 0 & 0 & 0 & -\mu_h - \mu_h & 0 \\
0 & 0 & 0 & 0 & 0 & 0 & -\rho_2 - \mu_h
\end{bmatrix}$$

$$-\mu_h$$

$$-\mu_m$$

$$\begin{bmatrix}
-\mu_h - \varphi & 0 & 0 & 0 & 0 \\
-\varphi & -\tau_1\eta_h - \tau_2\delta - \sigma - \mu_h & 0 & 0 & 0 \\
0 & -\tau_1\eta_h - \tau_2\delta & -\mu_h & 0 & 0 \\
0 & 0 & 0 & -\mu_h - \mu_h & 0 \\
0 & 0 & 0 & 0 & -\rho_2 - \mu_h
\end{bmatrix}$$

$$-\mu_h$$

$$(-\mu_h - \varphi)(-\tau_1\eta_h - \tau_2\delta - \sigma - \mu_h)$$

$$(-\mu_h - \mu_h)(-\rho_2 - \mu_h)$$

**Conclusion:** Since all the eigenvalues of the Jacobian matrix evaluated at the disease-free equilibrium have negative real parts under the stated parameter conditions, the disease-free equilibrium (E_0) is locally asymptotically stable. Hence, sufficiently small perturbations from the disease-free state decay with time, and trajectories starting sufficiently close to (E_0) return to the disease-free equilibrium.

**Theorem A4 (Global Stability for Endemic Equilibrium Points)**

The global stability of the endemic equilibrium $E^*$ensures that, regardless of initial conditions, all trajectories of the system converge to $E^*$whenever $R_0 > 1$, confirming that the disease persists endemically in the population.

**Proof**

Considering the **Goh-Volterra** type function, the Lyapunov function Lis constructed as a sum of Volterra-type terms of the form $x - x^* - x^*\ln(x/x^*)$for each state variable, which is always non-negative and equals zero if and only if $x = x^*$. This ensures L is a valid Lyapunov candidate

$$
\begin{aligned}
L = {} & a_1\left(S_w - S_w^* - S_w^*\ln\left(\frac{S_w}{S_w^*}\right)\right) + a_2\left(I_w - I_w^* - I_w^*\ln\left(\frac{I_w}{I_w^*}\right)\right) + a_3\left(B_w - B_w^* - B_w^*\ln\left(\frac{B_w}{B_w^*}\right)\right) \\
& + a_4\left(S_p - S_p^* - S_p^*\ln\left(\frac{S_p}{S_p^*}\right)\right) + a_5\left(I_p - I_p^* - I_p^*\ln\left(\frac{I_p}{I_p^*}\right)\right) + a_6\left(B_p - B_p^* - B_p^*\ln\left(\frac{B_p}{B_p^*}\right)\right) \\
& + a_7\left(S_m - S_m^* - S_m^*\ln\left(\frac{S_m}{S_m^*}\right)\right) + a_8\left(I_m - I_m^* - I_m^*\ln\left(\frac{I_m}{I_m^*}\right)\right) + a_9\left(S_h - S_h^* - S_h^*\ln\left(\frac{S_h}{S_h^*}\right)\right) \\
& + a_{10}\left(I_{Ah} - I_{Ah}^* - I_{Ah}^*\ln\left(\frac{I_{Ah}}{I_{Ah}^*}\right)\right) + a_{11}\left(I_h - I_h^* - I_h^*\ln\left(\frac{I_h}{I_h^*}\right)\right) + a_{12}\left(R_h - R_h^* - R_h^*\ln\left(\frac{R_h}{R_h^*}\right)\right) \\
& + a_{13}\left(V1_h - V1_h^* - V1_h^*\ln\left(\frac{V1_h}{V1_h^*}\right)\right) + a_{14}\left(V2_h - V2_h^* - V2_h^*\ln\left(\frac{V2_h}{V2_h^*}\right)\right)
\end{aligned}
$$

Differentiating L along the trajectories of the system and substituting the equilibrium conditions $E^*$, we obtain

$$
\begin{aligned}
\frac{dL}{dt} = {} & a_1\left(1 - \frac{S_w^*}{S_w}\right)S_w' + a_2\left(1 - \frac{I_w^*}{I_w}\right)I_w' + a_3\left(1 - \frac{B_w^*}{B_w}\right)B_w' + a_4\left(1 - \frac{S_p^*}{S_p}\right)S_p' + a_5\left(1 - \frac{I_p^*}{I_p}\right)I_p' \\
& + a_6\left(1 - \frac{B_p^*}{B_p}\right)B_p' + a_7\left(1 - \frac{S_m^*}{S_m}\right)S_m' + a_8\left(1 - \frac{I_m^*}{I_m}\right)I_m' + a_9\left(1 - \frac{S_h^*}{S_h}\right)S_h' + a_{10}\left(1 - \frac{I_{Ah}^*}{I_{Ah}}\right)I_{Ah}' \\
& + a_{11}\left(1 - \frac{I_h^*}{I_h}\right)I_h' + a_{12}\left(1 - \frac{R_h^*}{R_h}\right)R_h' + a_{13}\left(1 - \frac{V1_h^*}{V1_h}\right)V1_h' + a_{14}\left(1 - \frac{V2_h^*}{V2_h}\right)V2_h'
\end{aligned}
$$

$$
\begin{aligned}
\frac{dL}{dt} = {} & a_1\left(1-\frac{S_w^*}{S_w}\right)\left(\left(b_w-\frac{r_wN_w}{K_w}\right) - B\beta_{mw}b(T)\frac{I_m}{N_w}S_w-\varepsilon_wS_w-\mu_wS_w\right) \\
& + a_2\left(1-\frac{I_w^*}{I_w}\right)\left(B\beta_{mw}b(T)\frac{I_m}{N_w}S_w-\mu_wI_w\right)+a_3\left(1-\frac{B_w^*}{B_w}\right)(\varepsilon_wS_w-\mu_wB_w) \\
& + a_4\left(1-\frac{S_p^*}{S_p}\right)\left(b_p-C\beta_{mp}b(T)\frac{I_m}{N_p}S_p-\varepsilon_pS_p-\mu_pS_p+\alpha_2I_p\right) \\
& + a_5\left(1-\frac{I_p^*}{I_p}\right)\left(C\beta_{mp}b(T)\frac{I_m}{N_p}S_p-\alpha_2I_p-\mu_pI_p\right)+a_6\left(1-\frac{B_p^*}{B_p}\right)(\varepsilon_pS_p-\mu_pB_p) \\
& + a_7\left(1-\frac{S_m^*}{S_m}\right)\left((b_m-\frac{r_mN_m}{K_m}) - (C\beta_{pm}b(T)\frac{I_p}{N_p}S_m+B\beta_{wm}b(T)\frac{I_w}{N_w}S_m)-\mu_mS_m\right) \\
& + a_8\left(1-\frac{I_m^*}{I_m}\right)\left(\left(C\beta_{pm}b(T)\frac{I_p}{N_p}S_m+B\beta_{wm}b(T)\frac{I_w}{N_w}S_m\right)-\mu_mI_m\right) \\
& + a_9\left(1-\frac{S_h^*}{S_h}\right)\left((1-\alpha_1)b_h-\beta_{mh}b(T)\frac{I_m}{N_h}S_h-\mu_hS_h\right) \\
& + a_{10}\left(1-\frac{I_{Ah}^*}{I_{Ah}}\right)\left(\beta_{mh}b(T)\frac{I_m}{N_h}S_h-\mu_h-\varphi I_{Ah}\right) \\
& + a_{11}\left(1-\frac{I_h^*}{I_h}\right)(\varphi I_{Ah}-(\tau_1\eta_h+\tau_2\delta)I_h-\sigma I_h-\mu_hI_h) \\
& + a_{12}\left(1-\frac{R_h^*}{R_h}\right)((\tau_1\eta_h+\tau_2\delta)I_h-\mu_hR_h)+a_{13}\left(1-\frac{V1_h^*}{V1_h}\right)(u_1\alpha_1b_h-\rho_1V1_h-\mu_hV1_h) \\
& + a_{14}\left(1-\frac{V2_h^*}{V2_h}\right)((1-u_1)\alpha_1b_h-\rho_2V2_h-\mu_hV2_h)
\end{aligned}
$$

$$
\begin{aligned}
\frac{dL}{dt} = {} & a_1\left(1-\frac{S_w^*}{S_w}\right)\left(\frac{r_wN_w}{K_w}+ B\beta_{mw}b(T)\frac{I_m^*}{N_w}S_w^* + \varepsilon_w S_w^* + \mu_w S_w^* - \frac{r_wN_w}{K_w} - B\beta_{mw}b(T)\frac{I_m}{N_w}S_w - \varepsilon_w S_w \right.\\
& \left. - \mu_w S_w\right) + a_2\left(1-\frac{I_w^*}{I_w}\right)\left(B\beta_{mw}b(T)\frac{I_m}{N_w}S_w - B\beta_{mw}b(T)\frac{I_m^*}{N_w}S_w^*\right)\\
& + a_3\left(1-\frac{B_w^*}{B_w}\right)(\mu_w B_w^* - \mu_w B_w)\\
& + a_4\left(1-\frac{S_p^*}{S_p}\right)\left(C\beta_{mp}b(T)\frac{I_m^*}{N_p}S_p^* + \varepsilon_p S_p^* + \mu_p S_p^* - C\beta_{mp}b(T)\frac{I_m}{N_p}S_p - \varepsilon_p S_p - \mu_p S_p\right)\\
& + a_5\left(1-\frac{I_p^*}{I_p}\right)\left(C\beta_{mp}b(T)\frac{I_m}{N_p}S_p - C\beta_{mp}b(T)\frac{I_m^*}{N_p}S_p^*\right) + a_6\left(1-\frac{B_p^*}{B_p}\right)(\mu_p B_p^* - \mu_p B_p)\\
& + a_7\left(1-\frac{S_m^*}{S_m}\right)\left(\frac{r_mN_m}{K_m} + \left(C\beta_{pm}b(T)\frac{I_p^*}{N_p}S_m^* + B\beta_{wm}b(T)\frac{I_w^*}{N_w}S_m^*\right) + \mu_m S_m^* - \frac{r_mN_m}{K_m}\right.\\
& \left. - \left(C\beta_{pm}b(T)\frac{I_p}{N_p}S_m + B\beta_{wm}b(T)\frac{I_w}{N_w}S_m\right) - \mu_m S_m\right)\\
& + a_8\left(1-\frac{I_m^*}{I_m}\right)\left(\left(C\beta_{pm}b(T)\frac{I_p}{N_p}S_m + B\beta_{wm}b(T)\frac{I_w}{N_w}S_m\right)\right.\\
& \left. - \left(C\beta_{pm}b(T)\frac{I_p^*}{N_p}S_m^* + B\beta_{wm}b(T)\frac{I_w^*}{N_w}S_m^*\right)\right)\\
& + a_9\left(1-\frac{S_h^*}{S_h}\right)\left(\beta_{mh}b(T)\frac{I_m^*}{N_h}S_h^* + \mu_h S_h^* - \beta_{mh}b(T)\frac{I_m}{N_h}S_h - \mu_h S_h\right)\\
& + a_{10}\left(1-\frac{I_{Ah}^*}{I_{Ah}}\right)\left(\beta_{mh}b(T)\frac{I_m}{N_h}S_h - \beta_{mh}b(T)\frac{I_m^*}{N_h}S_h^*\right)\\
& + a_{11}\left(1-\frac{I_h^*}{I_h}\right)\left((\tau_1\eta_h + \tau_2\delta)I_h^* + \sigma I_h^* + \mu_h I_h^* - (\tau_1\eta_h + \tau_2\delta)I_h - \sigma I_h - \mu_h I_h\right)\\
& + a_{12}\left(1-\frac{R_h^*}{R_h}\right)(\mu_h R_h^* - \mu_h R_h) + a_{13}\left(1-\frac{V1_h^*}{V1_h}\right)(\rho_1 V1_h^* + \mu_h V1_h^* - \rho_1 V1_h - \mu_h V1_h)\\
& + a_{14}\left(1-\frac{V2_h^*}{V2_h}\right)(\rho_2 V2_h^* + \mu_h V2_h^* - \rho_2 V2_h - \mu_h V2_h)
\end{aligned}
$$

$$
\begin{aligned}
\frac{dL}{dt} = {} & a_1\left(1-\frac{S_w^*}{S_w}\right)\left(\frac{B\beta_{mw}b(T)}{N_w}(I_m^*S_w^*-I_mS_w)+(\varepsilon_w+\mu_w)(S_w^*-S_w)\right) \\
& + a_2\left(1-\frac{I_w^*}{I_w}\right)\left(\frac{B\beta_{mw}b(T)}{N_w}(I_mS_w-I_m^*S_w^*)\right)+a_3\left(1-\frac{B_w^*}{B_w}\right)\mu_w(B_w^*-B_w) \\
& + a_4\left(1-\frac{S_p^*}{S_p}\right)\left(\frac{C\beta_{mp}b(T)}{N_p}(I_m^*S_p^*-I_mS_p)+(\varepsilon_p+\mu_p)(S_p^*-S_p)\right) \\
& + a_5\left(1-\frac{I_p^*}{I_p}\right)\frac{C\beta_{mp}b(T)}{N_p}(I_mS_p-I_m^*S_p^*)+a_6\left(1-\frac{B_p^*}{B_p}\right)\mu_p(B_p^*-B_p) \\
& + a_7\left(1-\frac{S_m^*}{S_m}\right)\left(\left(\frac{C\beta_{pm}b(T)}{N_p}(I_p^*S_m^*-I_pS_m)+\frac{B\beta_{wm}b(T)}{N_w}(I_w^*S_m^*-I_wS_m)\right)+\mu_mS_m^*\right. \\
& \left.-\mu_mS_m\right)+a_8\left(1-\frac{I_m^*}{I_m}\right)\left(\frac{C\beta_{pm}b(T)}{N_p}(I_pS_m-I_p^*S_m^*)+\frac{B\beta_{wm}b(T)}{N_w}(I_wS_m-I_w^*S_m^*)\right) \\
& + a_9\left(1-\frac{S_h^*}{S_h}\right)\left(\frac{\beta_{mh}b(T)}{N_h}(I_m^*S_h^*-I_mS_h)+\mu_h(S_h^*-S_h)\right) \\
& + a_{10}\left(1-\frac{I_{Ah}^*}{I_{Ah}}\right)(I_mS_h-I_m^*S_h^*)\frac{\beta_{mh}b(T)}{N_h}+a_{11}\left(1-\frac{I_h^*}{I_h}\right)(\tau_1\eta_h+\tau_2\delta+\sigma+\mu_h)(I_h^*-I_h) \\
& + a_{12}\left(1-\frac{R_h^*}{R_h}\right)\mu_h(R_h^*-R_h)+a_{13}\left(1-\frac{V1_h^*}{V1_h}\right)(\rho_1+\mu_h)(V1_h^*-V1_h) \\
& + a_{14}\left(1-\frac{V2_h^*}{V2_h}\right)(\rho_2+\mu_h)(V2_h^*-V2_h)
\end{aligned}
$$

$$
\begin{aligned}
\frac{dL}{dt} = {} & \frac{B\beta_{mw}b(T)}{N_w}(I_m^*S_w^*-I_mS_w)\left(1-\frac{S_w^*}{S_w}\right)a_1-a_2\frac{B\beta_{mw}b(T)}{N_w}(I_m^*S_w^*-I_mS_w)\left(1-\frac{I_w^*}{I_w}\right) \\
& + a_1(\varepsilon_w+\mu_w)\left(2S_w^*-S_w-\frac{S_w^*}{S_w}S_w^*\right)+a_3\left(2B_w^*-B_w-\frac{B_w^*}{B_w}B_w^*\right)\mu_w \\
& + a_4\frac{C\beta_{mp}b(T)}{N_p}(I_m^*S_p^*-I_mS_p)\left(1-\frac{S_p^*}{S_p}\right)+a_4(\varepsilon_p+\mu_p)\left(2S_p^*-S_p-\frac{S_p^*}{S_p}S_p^*\right) \\
& + a_5\frac{C\beta_{mp}b(T)}{N_p}(I_mS_p-I_m^*S_p^*)\left(1-\frac{I_p^*}{I_p}\right)+a_6\left(2B_p^*-B_p-\frac{B_p^*}{B_p}B_p^*\right)\mu_p \\
& + a_7\frac{C\beta_{pm}b(T)}{N_p}(I_p^*S_m^*-I_pS_m)\left(1-\frac{S_m^*}{S_m}\right)+a_7\frac{B\beta_{wm}b(T)}{N_w}(I_w^*S_m^*-I_wS_m)\left(1-\frac{S_m^*}{S_m}\right) \\
& + a_7\mu_m\left(2S_m^*-S_m-\frac{S_m^*}{S_m}S_m^*\right)+a_8\frac{C\beta_{pm}b(T)}{N_p}(I_pS_m-I_p^*S_m^*)\left(1-\frac{I_m^*}{I_m}\right) \\
& + a_8\frac{B\beta_{wm}b(T)}{N_w}\left(1-\frac{I_m^*}{I_m}\right)(I_wS_m-I_w^*S_m^*)+a_9\frac{\beta_{mh}b(T)}{N_h}(I_m^*S_h^*-I_mS_h)\left(1-\frac{S_h^*}{S_h}\right) \\
& + a_9\mu_h(S_h^*-S_h)\left(1-\frac{S_h^*}{S_h}\right)+a_{10}\left(1-\frac{I_{Ah}^*}{I_{Ah}}\right)(I_mS_h-I_m^*S_h^*)\frac{\beta_{mh}b(T)}{N_h} \\
& + a_{11}\left(2I_h^*-I_h-\frac{I_h^*}{I_h}I_h^*\right)(\tau_1\eta_h+\tau_2\delta+\sigma+\mu_h)+a_{12}\left(2R_h^*-R_h-\frac{R_h^*}{R_h}R_h^*\right)\mu_h \\
& + a_{13}\left(2V1_h^*-V1_h-\frac{V1_h^*}{V1_h}V1_h^*\right)(\rho_1+\mu_h)+a_{14}\left(2V2_h^*-V2_h-\frac{V2_h^*}{V2_h}V2_h^*\right)(\rho_2+\mu_h)
\end{aligned}
$$

$$
\begin{aligned}
\frac{dL}{dt} = {} & \frac{B\beta_{mw}b(T)}{N_w}\left(I_m^*S_w^* - I_mS_w\right)\left(\left(1-\frac{S_w^*}{S_w}\right)a_1 - a_2\left(1-\frac{I_w^*}{I_w}\right)\right) + a_1(\varepsilon_w+\mu_w)S_w^*\left(2-\frac{S_w}{S_w^*}-\frac{S_w^*}{S_w}\right) \\
& + a_3B_w^*\left(2-\frac{B_w}{B_w^*}-\frac{B_w^*}{B_w}\right)\mu_w + \frac{C\beta_{mp}b(T)}{N_p}\left(I_m^*S_p^* - I_mS_p\right)\left(a_4\left(1-\frac{S_P^*}{S_p}\right) - a_5\left(1-\frac{I_p^*}{I_p}\right)\right) \\
& + a_4(\varepsilon_p+\mu_p)\left(2S_p^* - S_p - \frac{S_P^*}{S_p}S_p^*\right) + a_6B_p^*\left(2-\frac{B_p}{B_p^*}-\frac{B_p^*}{B_p}\right)\mu_p \\
& + \frac{C\beta_{pm}b(T)}{N_p}\left(I_p^*S_m^* - I_pS_m\right)\left(a_7\left(1-\frac{S_m^*}{S_m}\right) - a_8\left(1-\frac{I_m^*}{I_m}\right)\right) \\
& + \frac{B\beta_{wm}b(T)}{N_w}\left(I_w^*S_m^* - I_wS_m\right)\left(a_7\left(1-\frac{S_m^*}{S_m}\right) - a_8\left(1-\frac{I_m^*}{I_m}\right)\right) + a_7\mu_mS_m^*\left(2-\frac{S_m}{S_m^*}-\frac{S_m^*}{S_m}\right) \\
& + \frac{\beta_{mh}b(T)}{N_h}\left(I_m^*S_h^* - I_mS_h\right)\left(a_9\left(1-\frac{S_h^*}{S_h}\right) - a_{10}\left(1-\frac{I_{Ah}^*}{I_{Ah}}\right)\right) + a_9\mu_hS_h^*\left(2-\frac{S_h}{S_h^*}-\frac{S_h^*}{S_h}\right) \\
& + a_{11}I_h^*\left(2-\frac{I_h}{I_h^*}-\frac{I_h^*}{I_h}\right)(\tau_1\eta_h+\tau_2\delta+\sigma+\mu_h) + a_{12}R_h^*\left(2-\frac{R_h}{R_h^*}-\frac{R_h^*}{R_h}\right)\mu_h \\
& + a_{13}V1_h^*\left(2-\frac{V1_h}{V1_h^*}-\frac{V1_h^*}{V1_h}\right)(\rho_1+\mu_h) + a_{14}V2_h^*\left(2-\frac{V2_h}{V2_h^*}-\frac{V2_h^*}{V2_h}\right)(\rho_2+\mu_h)
\end{aligned}
$$

$$a_1 = a_2$$

$$a_4 = a_5$$

$$a_7 = a_8$$

$$a_9 = a_{10}$$

$$
\begin{aligned}
\frac{dL}{dt} = {} & a_1\frac{B\beta_{mw}b(T)}{N_w}\left(I_m^*S_w^* - I_mS_w\right)\left(\frac{I_w^*}{I_w}-\frac{S_w^*}{S_w}\right) + a_1(\varepsilon_w+\mu_w)S_w^*\left(2-\frac{S_w}{S_w^*}-\frac{S_w^*}{S_w}\right) + a_3B_w^*\left(2-\frac{B_w}{B_w^*}-\frac{B_w^*}{B_w}\right)\mu_w \\
& + a_4\frac{C\beta_{mp}b(T)}{N_p}\left(I_m^*S_p^* - I_mS_p\right)\left(\frac{I_p^*}{I_p}-\frac{S_P^*}{S_p}\right) + a_4(\varepsilon_p+\mu_p)S_p^*\left(2-\frac{S_p}{S_p^*}-\frac{S_P^*}{S_p}\right) \\
& + a_6B_p^*\left(2-\frac{B_p}{B_p^*}-\frac{B_p^*}{B_p}\right)\mu_p + a_7\frac{C\beta_{pm}b(T)}{N_p}\left(I_p^*S_m^* - I_pS_m\right)\left(\frac{I_m^*}{I_m}-\frac{S_m^*}{S_m}\right) \\
& + a_7\frac{B\beta_{wm}b(T)}{N_w}\left(I_w^*S_m^* - I_wS_m\right)\left(\frac{I_m^*}{I_m}-\frac{S_m^*}{S_m}\right) + a_7\mu_mS_m^*\left(2-\frac{S_m}{S_m^*}-\frac{S_m^*}{S_m}\right) \\
& + a_9\frac{\beta_{mh}b(T)}{N_h}\left(I_m^*S_h^* - I_mS_h\right)\left(\frac{I_{Ah}^*}{I_{Ah}}-\frac{S_h^*}{S_h}\right) + a_9\mu_hS_h^*\left(2-\frac{S_h}{S_h^*}-\frac{S_h^*}{S_h}\right) \\
& + a_{11}I_h^*\left(2-\frac{I_h}{I_h^*}-\frac{I_h^*}{I_h}\right)(\tau_1\eta_h+\tau_2\delta+\sigma+\mu_h) + a_{12}R_h^*\left(2-\frac{R_h}{R_h^*}-\frac{R_h^*}{R_h}\right)\mu_h \\
& + a_{13}V1_h^*\left(2-\frac{V1_h}{V1_h^*}-\frac{V1_h^*}{V1_h}\right)(\rho_1+\mu_h) + a_{14}V2_h^*\left(2-\frac{V2_h}{V2_h^*}-\frac{V2_h^*}{V2_h}\right)(\rho_2+\mu_h)
\end{aligned}
$$

$$\frac{dL}{dt} = a_1 \frac{B\beta_{mw} b(T)}{N_w}\left(I_m^* S_w^* - I_m S_w\right)\left(\frac{I_w^*}{I_w} - \frac{S_w^*}{S_w}\right) + a_1(\varepsilon_w + \mu_w)S_w^*\left(2 - \frac{S_w}{S_w^*} - \frac{S_w^*}{S_w}\right) + a_3 B_w^*\left(2 - \frac{B_w}{B_w^*} - \frac{B_w^*}{B_w}\right)\mu_w$$
$$+ a_4 \frac{C\beta_{mp} b(T)}{N_p}\left(I_m^* S_p^* - I_m S_p\right)\left(\frac{I_p^*}{I_p} - \frac{S_P^*}{S_p}\right) + a_4(\varepsilon_p + \mu_p)S_p^*\left(2 - \frac{S_p}{S_p^*} - \frac{S_P^*}{S_p}\right)$$
$$+ a_6 B_p^*\left(2 - \frac{B_p}{B_p^*} - \frac{B_p^*}{B_p}\right)\mu_p + a_7 \frac{C\beta_{pm} b(T)}{N_p}\left(I_p^* S_m^* - I_p S_m\right)\left(\frac{I_m^*}{I_m} - \frac{S_m^*}{S_m}\right)$$
$$+ a_7 \frac{B\beta_{wm} b(T)}{N_w}\left(I_w^* S_m^* - I_w S_m\right)\left(\frac{I_m^*}{I_m} - \frac{S_m^*}{S_m}\right) + a_7 \mu_m S_m^*\left(2 - \frac{S_m}{S_m^*} - \frac{S_m^*}{S_m}\right)$$
$$+ a_9 \frac{\beta_{mh} b(T)}{N_h}\left(I_m^* S_h^* - I_m S_h\right)\left(\frac{I_{Ah}^*}{I_{Ah}} - \frac{S_h^*}{S_h}\right) + a_9 \mu_h S_h^*\left(2 - \frac{S_h}{S_h^*} - \frac{S_h^*}{S_h}\right)$$
$$+ a_{11} I_h^*\left(2 - \frac{I_h}{I_h^*} - \frac{I_h^*}{I_h}\right)(\tau_1\eta_h + \tau_2\delta + \sigma + \mu_h) + a_{12} R_h^*\left(2 - \frac{R_h}{R_h^*} - \frac{R_h^*}{R_h}\right)\mu_h$$
$$+ a_{13} V1_h^*\left(2 - \frac{V1_h}{V1_h^*} - \frac{V1_h^*}{V1_h}\right)(\rho_1 + \mu_h) + a_{14} V2_h^*\left(2 - \frac{V2_h}{V2_h^*} - \frac{V2_h^*}{V2_h}\right)(\rho_2 + \mu_h)$$

$$\frac{dL}{dt} = a_1(\varepsilon_w + \mu_w)S_w^*\left(2 - \frac{S_w}{S_w^*} - \frac{S_w^*}{S_w}\right) + a_3 B_w^*\left(2 - \frac{B_w}{B_w^*} - \frac{B_w^*}{B_w}\right)\mu_w + a_4(\varepsilon_p + \mu_p)S_p^*\left(2 - \frac{S_p}{S_p^*} - \frac{S_P^*}{S_p}\right)$$
$$+ a_6 B_p^*\left(2 - \frac{B_p}{B_p^*} - \frac{B_p^*}{B_p}\right)\mu_p + a_7 \mu_m S_m^*\left(2 - \frac{S_m}{S_m^*} - \frac{S_m^*}{S_m}\right) + a_9 \mu_h S_h^*\left(2 - \frac{S_h}{S_h^*} - \frac{S_h^*}{S_h}\right)$$
$$+ a_{11} I_h^*\left(2 - \frac{I_h}{I_h^*} - \frac{I_h^*}{I_h}\right)(\tau_1\eta_h + \tau_2\delta + \sigma + \mu_h) + a_{12} R_h^*\left(2 - \frac{R_h}{R_h^*} - \frac{R_h^*}{R_h}\right)\mu_h$$
$$+ a_{13} V1_h^*\left(2 - \frac{V1_h}{V1_h^*} - \frac{V1_h^*}{V1_h}\right)(\rho_1 + \mu_h) + a_{14} V2_h^*\left(2 - \frac{V2_h}{V2_h^*} - \frac{V2_h^*}{V2_h}\right)(\rho_2 + \mu_h)$$
$$+ a_1 \frac{B\beta_{mw} b(T)}{N_w}\left(I_m^* S_w^* - I_m S_w\right)\left(\frac{I_w^*}{I_w} - \frac{S_w^*}{S_w} + \frac{I_m^*}{I_m} - \frac{S_m^*}{S_m}\right)$$
$$+ a_4 \frac{C\beta_{mp} b(T)}{N_p}\left(\left(I_m^* S_p^* - I_m S_p\right)\left(\frac{I_p^*}{I_p} - \frac{S_P^*}{S_p}\right) + \left(I_p^* S_m^* - I_p S_m\right)\left(\frac{I_m^*}{I_m} - \frac{S_m^*}{S_m}\right)\right)$$
$$+ a_9 \frac{\beta_{mh} b(T)}{N_h}\left(I_m^* S_h^* - I_m S_h\right)\left(\frac{I_{Ah}^*}{I_{Ah}} - \frac{S_h^*}{S_h}\right)$$

$$a_1 = \frac{\beta_{wm} I_w^* S_m^*}{\beta_{mw} I_m^* S_w^*} = a_2 = a_3$$

$$a_4 = \frac{\beta_{pm} I_p^* S_m^*}{\beta_{mp} I_m^* S_p^*} = a_5 = a_6$$

$$a_9 = \frac{\left(\frac{\beta_{mh} b(T) I_m^* S_h^*}{N_h}\right)}{\mu S_h^*\left(\frac{\beta_{mh} b(T) I_m^*}{N_h} + \mu_h\right)} = a_{10} = a_{11} = a_{12} = a_{13} = a_{14}$$

$$a_7 = 1$$

$$
\begin{aligned}
\frac{dL}{dt} = {} & \left(\frac{\beta_{wm} I_w^* S_m^*}{\beta_{mw} I_m^* S_w^*}\right)(\varepsilon_w + \mu_w) S_w^* \left(2 - \frac{S_w}{S_w^*} - \frac{S_w^*}{S_w}\right) + \left(\frac{\beta_{wm} I_w^* S_m^*}{\beta_{mw} I_m^* S_w^*}\right) B_w^* \left(2 - \frac{B_w}{B_w^*} - \frac{B_w^*}{B_w}\right) \mu_w \\
& + \left(\frac{\beta_{pm} I_p^* S_m^*}{\beta_{mp} I_m^* S_p^*}\right)(\varepsilon_p + \mu_p) S_p^* \left(2 - \frac{S_p}{S_p^*} - \frac{S_P^*}{S_p}\right) + \left(\frac{\beta_{pm} I_p^* S_m^*}{\beta_{mp} I_m^* S_p^*}\right) B_p^* \left(2 - \frac{B_p}{B_p^*} - \frac{B_p^*}{B_p}\right) \mu_p \\
& + \mu_m S_m^* \left(2 - \frac{S_m}{S_m^*} - \frac{S_m^*}{S_m}\right) + \left(\frac{\left(\frac{\beta_{mh} b(T) I_m^* S_h^*}{N_h}\right)}{\mu S_h^* \left(\frac{\beta_{mh} b(T) I_m^*}{N_h} + \mu_h\right)}\right) \mu_h S_h^* \left(2 - \frac{S_h}{S_h^*} - \frac{S_h^*}{S_h}\right) \\
& + \left(\frac{\left(\frac{\beta_{mh} b(T) I_m^* S_h^*}{N_h}\right)}{\mu S_h^* \left(\frac{\beta_{mh} b(T) I_m^*}{N_h} + \mu_h\right)}\right) I_h^* \left(2 - \frac{I_h}{I_h^*} - \frac{I_h^*}{I_h}\right)(\tau_1 \eta_h + \tau_2 \delta + \sigma + \mu_h) \\
& + \left(\frac{\left(\frac{\beta_{mh} b(T) I_m^* S_h^*}{N_h}\right)}{\mu S_h^* \left(\frac{\beta_{mh} b(T) I_m^*}{N_h} + \mu_h\right)}\right) R_h^* \left(2 - \frac{R_h}{R_h^*} - \frac{R_h^*}{R_h}\right) \mu_h \\
& + \left(\frac{\left(\frac{\beta_{mh} b(T) I_m^* S_h^*}{N_h}\right)}{\mu S_h^* \left(\frac{\beta_{mh} b(T) I_m^*}{N_h} + \mu_h\right)}\right) V1_h^* \left(2 - \frac{V1_h}{V1_h^*} - \frac{V1_h^*}{V1_h}\right)(\rho_1 + \mu_h) \\
& + \frac{\left(\frac{\beta_{mh} b(T) I_m^* S_h^*}{N_h}\right)}{\mu S_h^* \left(\frac{\beta_{mh} b(T) I_m^*}{N_h} + \mu_h\right)} V2_h^* \left(2 - \frac{V2_h}{V2_h^*} - \frac{V2_h^*}{V2_h}\right)(\rho_2 + \mu_h) \\
& + \left(\frac{\beta_{wm} I_w^* S_m^*}{\beta_{mw} I_m^* S_w^*}\right) \frac{B \beta_{mw} b(T)}{N_w} (I_m^* S_w^* - I_m S_w) \left(\frac{I_w^*}{I_w} - \frac{S_w^*}{S_w} + \frac{I_m^*}{I_m} - \frac{S_m^*}{S_m}\right) \\
& + \left(\frac{\beta_{pm} I_p^* S_m^*}{\beta_{mp} I_m^* S_p^*}\right) \frac{C \beta_{mp} b(T)}{N_p} \left( (I_m^* S_p^* - I_m S_p) \left(\frac{I_p^*}{I_p} - \frac{S_P^*}{S_p}\right) + (I_p^* S_m^* - I_p S_m) \left(\frac{I_m^*}{I_m} - \frac{S_m^*}{S_m}\right) \right) \\
& + \left(\frac{\left(\frac{\beta_{mh} b(T) I_m^* S_h^*}{N_h}\right)}{\mu S_h^* \left(\frac{\beta_{mh} b(T) I_m^*}{N_h} + \mu_h\right)}\right) \frac{\beta_{mh} b(T)}{N_h} (I_m^* S_h^* - I_m S_h) \left(\frac{I_{Ah}^*}{I_{Ah}} - \frac{S_h^*}{S_h}\right)
\end{aligned}
$$

$$
\begin{aligned}
\frac{dL}{dt} = {} & \left(\frac{\beta_{wm} I_w^* S_m^*}{\beta_{mw} I_m^* S_w^*}\right)(\varepsilon_w + \mu_w) S_w^* \left(2 - \frac{S_w}{S_w^*} - \frac{S_w^*}{S_w}\right) + \left(\frac{\beta_{wm} I_w^* S_m^*}{\beta_{mw} I_m^* S_w^*}\right) B_w^* \left(2 - \frac{B_w}{B_w^*} - \frac{B_w^*}{B_w}\right) \mu_w \\
& + \left(\frac{\beta_{pm} I_p^* S_m^*}{\beta_{mp} I_m^* S_p^*}\right)(\varepsilon_p + \mu_p) S_p^* \left(2 - \frac{S_p}{S_p^*} - \frac{S_P^*}{S_p}\right) + \left(\frac{\beta_{pm} I_p^* S_m^*}{\beta_{mp} I_m^* S_p^*}\right) B_p^* \left(2 - \frac{B_p}{B_p^*} - \frac{B_p^*}{B_p}\right) \mu_p \\
& + \mu_m S_m^* \left(2 - \frac{S_m}{S_m^*} - \frac{S_m^*}{S_m}\right) + \left(\frac{\left(\frac{\beta_{mh} b(T) I_m^* S_h^*}{N_h}\right)}{\mu S_h^* \left(\frac{\beta_{mh} b(T) I_m^*}{N_h} + \mu_h\right)}\right) \mu_h S_h^* \left(2 - \frac{S_h}{S_h^*} - \frac{S_h^*}{S_h}\right) \\
& + \left(\frac{\left(\frac{\beta_{mh} b(T) I_m^* S_h^*}{N_h}\right)}{\mu S_h^* \left(\frac{\beta_{mh} b(T) I_m^*}{N_h} + \mu_h\right)}\right) I_h^* \left(2 - \frac{I_h}{I_h^*} - \frac{I_h^*}{I_h}\right)(\tau_1 \eta_h + \tau_2 \delta + \sigma + \mu_h) \\
& + \left(\frac{\left(\frac{\beta_{mh} b(T) I_m^* S_h^*}{N_h}\right)}{\mu S_h^* \left(\frac{\beta_{mh} b(T) I_m^*}{N_h} + \mu_h\right)}\right) R_h^* \left(2 - \frac{R_h}{R_h^*} - \frac{R_h^*}{R_h}\right) \mu_h \\
& + \left(\frac{\left(\frac{\beta_{mh} b(T) I_m^* S_h^*}{N_h}\right)}{\mu S_h^* \left(\frac{\beta_{mh} b(T) I_m^*}{N_h} + \mu_h\right)}\right) V1_h^* \left(2 - \frac{V1_h}{V1_h^*} - \frac{V1_h^*}{V1_h}\right)(\rho_1 + \mu_h) \\
& + \frac{\left(\frac{\beta_{mh} b(T) I_m^* S_h^*}{N_h}\right)}{\mu S_h^* \left(\frac{\beta_{mh} b(T) I_m^*}{N_h} + \mu_h\right)} V2_h^* \left(2 - \frac{V2_h}{V2_h^*} - \frac{V2_h^*}{V2_h}\right)(\rho_2 + \mu_h) \\
& + \left(\frac{\beta_{wm} I_w^* S_m^*}{\beta_{mw} I_m^* S_w^*}\right) \frac{B \beta_{mw} b(T)}{N_w} (I_m^* S_w^* - I_m S_w) \left(\frac{I_w^*}{I_w} - \frac{S_w^*}{S_w} + \frac{I_m^*}{I_m} - \frac{S_m^*}{S_m}\right) \\
& + \left(\frac{\beta_{pm} I_p^* S_m^*}{\beta_{mp} I_m^* S_p^*}\right) \frac{C \beta_{mp} b(T)}{N_p} \left((I_m^* S_p^* - I_m S_p)\left(\frac{I_p^*}{I_p} - \frac{S_P^*}{S_p}\right) + (I_p^* S_m^* - I_p S_m)\left(\frac{I_m^*}{I_m} - \frac{S_m^*}{S_m}\right)\right) \\
& + \left(\frac{\left(\frac{\beta_{mh} b(T) I_m^* S_h^*}{N_h}\right)}{\mu S_h^* \left(\frac{\beta_{mh} b(T) I_m^*}{N_h} + \mu_h\right)}\right) \frac{\beta_{mh} b(T)}{N_h} (I_m^* S_h^* - I_m S_h) \left(\frac{I_{Ah}^*}{I_{Ah}} - \frac{S_h^*}{S_h}\right)
\end{aligned}
$$

**Since $\frac{dL}{dt} \le 0$ holds universally with $\frac{dL}{dt} = 0$ only at $E^*$ , the endemic equilibrium is globally asymptotically stable by LaSalle's Invariance Principle.**

#### Theorem A5 (Optimal Control Theorem)

Theorem: Characterization of the Optimal Control

Consider the 14-compartment Japanese encephalitis transmission model with state vector

$$X(t) = \left(S_w, I_w, B_w, S_p, I_p, B_p, S_m, I_m, S_h, I_A h, I_h, R_h, V_1 h, V_2 h\right).$$

Let $\vartheta_1(t)$, $\vartheta_2(t)$, $\vartheta_3(t)$, $\vartheta_4(t)$ and $\vartheta_5(t)$ denote, respectively, time-dependent controls for wading-bird barrier protection, pig biosecurity, treatment of symptomatic infected humans, dose-series-I human vaccination and dose-series-II human vaccination. The admissible control set is

$$U = \vartheta = (\vartheta_1, \ldots, \vartheta_5): \vartheta_i is\ measurable\ and 0 \le \vartheta_i(t) \le 1, i = 1, \ldots, 5, t \in [0, T_f].$$

$Let f_j(X, \vartheta)$,j=1,...,14, denote the right-hand sides of the corrected controlled state equations obtained from the original JE model by introducing the controls only into their corresponding intervention terms. Define the objective functional

$$J(\vartheta) = \int_0^{T_f} \left[A_w I_w + A_p I_p + A_m I_m + A_A I_A h + A_h I_h + ½(B_1 \vartheta_1{}^2 + B_2 \vartheta_2{}^2 + B_3 \vartheta_3{}^2 + B_4 \vartheta_4{}^2 + B_5 \vartheta_5{}^2)\right] dt.$$

Here $A_w, A_p, A_m, A_A, A_h > 0$ measure the relative epidemiological importance of the infected classes, whereas $B_1,\ldots,B_5>0$ represent the relative costs of the interventions.

Assume that the controlled state system admits a non-negative solution on [0,T_f], that the right-hand sides are continuously differentiable with respect to the state variables and controls on the biologically feasible region, and that the admissible control set is non-empty, closed and convex. Then there exists an optimal control $\vartheta^*=(\vartheta_1^*,\ldots,\vartheta_5^*)\in U$ minimizing J, subject to the controlled state system. Moreover, there exist adjoint variables $\lambda_1,\ldots,\lambda_{14}$ satisfying

$$d\lambda_j/dt = -\partial H/\partial x_j, j = 1, \ldots, 14,$$

$\lambda_j(T_f) = 0, j = 1, \ldots, 14,$

where the Hamiltonian is

$$H = A_w I_w + A_p I_p + A_m I_m + A_A I_A h + A_h I_h + ½\Sigma_{i=1}{}^{5} B_i \vartheta_i{}^2 + \Sigma_{j=1}{}^{14} \lambda_j f_j(X, \vartheta).$$

At almost every t∈[0,T_f], the optimal control minimizes H over the admissible control interval.

#### Proof

For each intervention, the intensity is restricted to the closed interval [0,1]. Hence U is non-empty. The interval [0,1] is closed and convex, and therefore the Cartesian product defining the five-control set is also closed and convex. Measurability ensures that the controls can vary with time while remaining admissible.

For a fixed admissible control $\vartheta\in U$, the controlled JE model is a finite system of ordinary differential equations. On any biologically feasible bounded region for which the population denominators $N_w, N_p$and N_h remain positive, the incidence and transfer terms are continuous and locally Lipschitz in the state variables. Thus the state system has a unique local solution for the prescribed initial conditions. The optimal-control theorem is applied on a finite time interval $[0, T_f]$ under the stated assumption that this solution remains in the non-negative feasible region.

The running cost is linear in the infected state variables and quadratic in the controls. Since B_i>0,

$$\partial^2 L/\partial \vartheta_i{}^2 = B_i > 0, i = 1, \ldots, 5.$$

Therefore, the running cost is strictly convex with respect to each control. The quadratic terms also penalize unnecessarily large intervention intensities and provide the standard coercive structure used in optimal-control formulations.

Introduce one adjoint variable for each of the 14 state variables. With x_j denoting the j-th component of X, the Hamiltonian is the sum of the running objective and the inner product between the adjoint vector and the controlled state vector field:

$$H = L(X, \vartheta) + \Sigma_{j=1}{}^{14} \lambda_j f_j(X, \vartheta).$$

Pontryagin's Maximum Principle gives the necessary adjoint equations

$$\dot{\lambda}_j = -\partial H/\partial x_j, j = 1, \ldots, 14.$$

Because there is no terminal-state cost in J and the terminal states are free, the corresponding transversality conditions are

$$\lambda_j(T_f) = 0, j = 1, \ldots, 14.$$

Consequently, the optimality system is a two-point boundary-value problem: the state equations are solved forward from their initial conditions, while the adjoint equations are solved backward from the terminal conditions.

In the corrected controlled bird subsystem, $\vartheta_1$ transfers susceptible wading birds from $S_w$ to $B_w$at rate $\vartheta_1 \varepsilon_w S_w$. Therefore, the terms of H containing $\vartheta_1$ are

$$H(\vartheta_1) = ½ B_1 \vartheta_1{}^2 - \lambda_S w \vartheta_1 \varepsilon_w S_w + \lambda_B w \vartheta_1 \varepsilon_w S_w + termsindependentof\vartheta_1.$$

For an interior optimum, $\partial H/\partial \vartheta_1=0$, giving

$$B_1\vartheta_1 - \lambda_S w\varepsilon_w S_w + \lambda_B w\varepsilon_w S_w = 0,$$

$$\vartheta_1 = \left((\lambda_S w - \lambda_B w)\varepsilon_w S_w\right)/B_1.$$

Projection onto the admissible interval [0,1] yields

$$\vartheta_1 *= max0, min\left[1, \left((\lambda_S w - \lambda_B w)\varepsilon_w S_w\right)/B_1\right].$$

Similarly, $\vartheta_2$ transfers susceptible pigs from$S_p$ to $B_p$ at rate$\vartheta_2\varepsilon_p S_p$. Hence

$$\partial H/\partial\vartheta_2 = B_2\vartheta_2 - \lambda_S p\varepsilon_p S_p + \lambda_B p\varepsilon_p S_p.$$

Setting this derivative equal to zero and projecting onto [0,1] gives

$$\vartheta_2 *= max0, min\left[1, \left((\lambda_S p - \lambda_B p)\varepsilon_p S_p\right)/B_2\right].$$

The treatment control $\vartheta_3$ increases the treatment-mediated transfer from symptomatic infection $I_h$ to recovery$R_h$through the term $\vartheta_3\tau_2\delta I_h$. The $\vartheta_3$-dependent Hamiltonian terms therefore give

$$\partial H/\partial\vartheta_3 = B_3\vartheta_3 - \lambda_I h\tau_2\delta I_h + \lambda_R h\tau_2\delta I_h.$$

Thus

$$\vartheta_3 *= max0, min\left[1, \left((\lambda_I h - \lambda_R h)\tau_2\delta I_h\right)/B_3\right].$$

The same differentiation procedure applies to $\vartheta_4$ and $\vartheta_5$. However, the supplied manuscript describes vaccination as movement of susceptible humans into dose series I and subsequently into dose series II, while the exact controlled source/sink equations are not fully fixed in the present Appendix formulation. Therefore a unique explicit formula for $\vartheta_4$* and $\vartheta_5$* should be written only after the vaccination transfer terms are specified consistently. In general,

$$\vartheta_4 *= Proj_{[0,1]} - (1/B_4)\partial\left[\Sigma\lambda_j f_j\right]/\partial\vartheta_4,$$

$$\vartheta_5 *= Proj_{[0,1]} - (1/B_5)\partial\left[\Sigma\lambda_j f_j\right]/\partial\vartheta_5.$$

Once the exact S_h→V_1h and V_1h→V_2h transfers are fixed, these derivatives give the corresponding explicit vaccination-control characterizations.

The admissible control set is closed and convex, the running cost is convex in the controls, and the Hamiltonian satisfies the pointwise minimum condition. Together with the assumed existence of non-negative state trajectories and the adjoint equations with transversality conditions, these relations provide the necessary optimality system. Hence an optimal intervention strategy, when it exists under the stated feasibility assumptions, is characterized by the forward state system, backward adjoint system and projected control formulas. This completes the characterization.

#### Results

The theorem produces a mathematical rule for choosing intervention intensity as a function of time rather than assuming that the same intervention level is applied throughout the epidemic. Its principal outputs are:

A coupled optimality system containing 14 forward state equations and 14 backward adjoint equations.

Terminal conditions $\lambda_j(T_f) = 0$ for all 14 adjoint variables.

A bounded optimal wading-bird barrier strategy $\vartheta_1$*(t).

A bounded optimal pig biosecurity strategy $\vartheta_2$*(t).

A bounded optimal human treatment strategy $\vartheta_3$*(t).

General optimality conditions for the two vaccination controls, to be made explicit after the vaccination transfers are fixed.

A formal balance between epidemiological benefit (reduction of infected populations) and intervention cost through the weights$A_i$ and $B_i$.

The projected form of each control has an important interpretation. If the marginal benefit of increasing an intervention is sufficiently large relative to its cost, the optimal control moves toward its upper bound. If the marginal benefit is small relative to the cost, the control moves toward zero. Intermediate values correspond to situations in which epidemiological benefit and marginal control cost are balanced.

#### Conclusion

The optimal control theorem extends the 14-compartment Japanese encephalitis transmission model from a descriptive epidemiological system to a theoretical decision framework for time-dependent intervention. By applying Pontryagin's Maximum Principle, the analysis links the disease states, adjoint variables, and intervention costs to characterize the intensity of wading-bird barrier protection, pig biosecurity, human treatment, and vaccination over time.

The key theoretical result is that the optimal intervention is not necessarily constant throughout the control period. Instead, each control is determined dynamically by its marginal effect on the epidemic and its assigned implementation cost, subject to the feasible bounds $0 \leq \vartheta_i \leq 1$. The resulting state-adjoint-control system therefore provides the mathematical basis for identifying when an intervention should be intensified, maintained, or relaxed.

In summary, the optimal-control analysis establishes a rigorous theoretical framework for balancing JE infection reduction against intervention costs. The derived state-adjoint system and bounded control characterizations provide the necessary conditions for determining time-dependent intervention intensities. Numerical implementation may subsequently be undertaken after calibration of the intervention-cost weights and complete specification of the vaccination transfer parameters.
